# Epidemics of chikungunya, Zika, and COVID-19 reveal bias in case-based mapping

**DOI:** 10.1101/2021.07.23.21261038

**Authors:** Fausto Andres Bustos Carrillo, Brenda Lopez Mercado, Jairo Carey Monterrey, Damaris Collado, Saira Saborio, Tatiana Miranda, Carlos Barilla, Sergio Ojeda, Nery Sanchez, Miguel Plazaola, Harold Suazo Laguna, Douglas Elizondo, Sonia Arguello, Anna M. Gajewski, Hannah E. Maier, Krista Latta, Bradley Carlson, Josefina Coloma, Leah Katzelnick, Hugh Sturrock, Angel Balmaseda, Guillermina Kuan, Aubree Gordon, Eva Harris

## Abstract

Accurate tracing of epidemic spread over space enables effective control measures. We examined three metrics of infection and disease in a pediatric cohort (N ≈ 3,000) over two chikungunya and one Zika epidemic, and in a household cohort (N=1,793) over one COVID-19 epidemic in Managua, Nicaragua. We compared spatial incidence rates (cases/total population), infection risks (infections/total population), and disease risks (cases/infected population). We used generalized additive and mixed-effects models, Kulldorf’s spatial scan statistic, and intracluster correlation coefficients. Across different analyses and all epidemics, incidence rates considerably underestimated infection and disease risks, producing large and spatially non-uniform biases distinct from biases due to incomplete case ascertainment. Infection and disease risks exhibited distinct spatial patterns, and incidence clusters inconsistently identified areas of either risk. While incidence rates are commonly used to infer infection and disease risk in a population, we find that this can induce substantial biases and adversely impact policies to control epidemics.

**Article summary line:** Inferring measures of spatial risk from case-only data can substantially bias estimates, thereby weakening and potentially misdirecting measures needed to control an epidemic.

## INTRODUCTION

Controlling epidemic spread requires accurate data on the movement of pathogens through populations. Standard spatial studies of infectious diseases use passively collected, individual-level data for cases (symptomatic infections) from health facilities after cases present for medical treatment (1–5). Then, by using census data to obtain the total population in an area, these studies estimate incidence rates (attack rates, incidence proportions) as the ratio of cases to the total population. However, because this approach does not capture subclinical (clinically inapparent) infections, this incidence approach may not recapitulate the spatial contour of infections, which may have a distinct pattern and magnitude. These issues may be compounded when the incidence rate, estimated from passively collected and hence incomplete case data, is used to infer infection risk (1–3) or disease risk (4,5) in policy decision-making on epidemic control.

Epidemiological risk is the probability of a susceptible individual experiencing an outcome. For an immunologically naïve population, all persons are at risk for an initial infection. However, only infected individuals are at risk for experiencing illness, as only infected persons are susceptible to disease. Consequently, measuring infection status is necessary to estimate the numerator of infection risk (infections/total population) and the denominator of disease risk (cases/infected population). These metrics are related to the incidence rate through an application of conditional probability, expressed in multiple ways below:

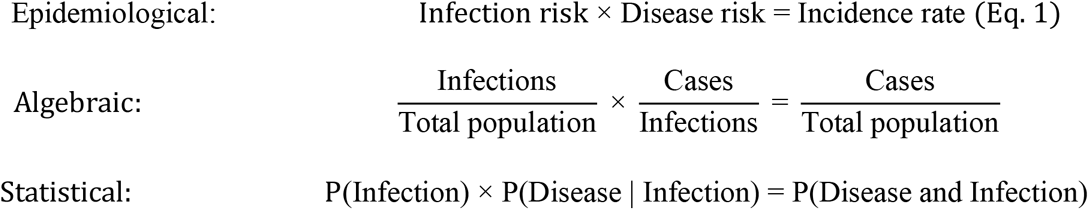

Eq. 1, applicable to infectious disease epidemics in an initially naïve population, demonstrates that incidence is the product of two underlying probabilities of interest. Thus, the incidence rate is explained by, and can be decoupled into, infection and disease risks.

We spatially analyzed four explosive epidemics in two longitudinal Nicaraguan cohorts. Our analysis covers the 2014 and 2015 chikungunya epidemics caused by chikungunya virus (CHIKV) (6,7), the 2016 Zika epidemic caused by Zika virus (ZIKV) (8,9), and the first wave of the COVID-19 epidemic in 2020 caused by severe acute respiratory syndrome coronavirus 2 (SARS-CoV-2) (10). While *Aedes* mosquitoes transmit CHIKV and ZIKV (11), SARS-CoV-2 primarily spreads by respiratory droplets (12). We analyzed the epidemics in parallel to identify commonalities across epidemics of different pathogens and transmission routes. We demonstrated differences in the fine-scale spatial characterization of epidemics by standard incidence-based measures versus a more comprehensive approach that included infection and disease risks. Finally, we quantified and mapped the separate biases induced by using passive versus active surveillance.

## METHODS

### Ethics statement

The Pediatric Dengue Cohort Study (PDCS) was approved by Institutional Review Boards (IRBs) of the University of California, Berkeley; the University of Michigan, Ann Arbor; and the Nicaraguan Ministry of Health. The Household Influenza Cohort Study (HICS) was approved by the University of Michigan, Ann Arbor, and the Nicaraguan Ministry of Health IRBs. Participants’ parents or legal guardians provided written informed consent. Subjects six years and older provided verbal assent.

### Study design and eligibility criteria

The PDCS (13) is an open, population-based, prospective cohort of children initiated in 2004 to study dengue virus and later expanded to include CHIKV and ZIKV. We assessed ∼3,000 PDCS participants 2-14 years old who experienced two chikungunya epidemics and one Zika epidemic (6–9). The HICS is an open, population-based, prospective cohort of households that has studied influenza virus and coronaviruses since 2017. We evaluated 1,793 HICS participants 0-87 years old who experienced the first COVID-19 epidemic (10). The age structure of the HICS is representative of Managua’s general population.

Both cohort studies share the same study site (Fig. 1) in Managua, Nicaragua’s capital. During the studies’ annual sampling (serosurvey) in March/April, participants provide blood samples to ascertain infection status during the prior year. A mid-year sampling was instituted in the HICS in October/November 2020 to measure SARS-CoV-2 infections after the first COVID-19 wave but before the second. Both studies provide participants with primary care; participants agree to visit the study health center at the first indication of any illness.

**Figure 1.**
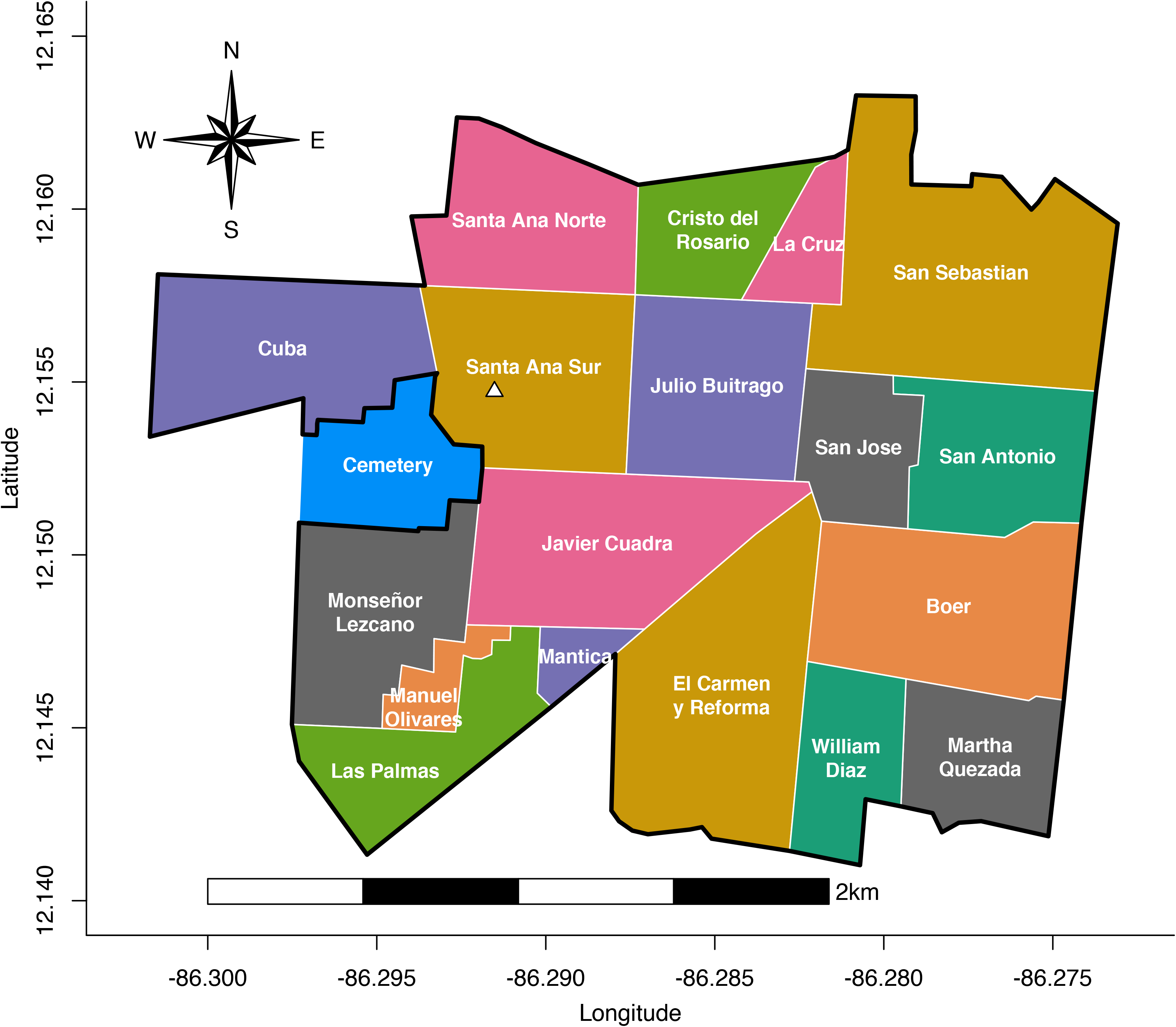
The neighborhoods of the study area in Managua, Nicaragua. The cemetery is shown in blue, and the study health center is indicated by a white triangle.

Analysis of each epidemic was restricted to participants who lived within the health center’s catchment area and were immunologically naïve. By further restricting to participants who were enrolled before each epidemic, we analyzed a closed cohort of initially uninfected participants who subsequently experienced an epidemic. The Appendix (pages 1-3) contains additional study design information.

### Laboratory methods

Upon collection, annual blood samples were immediately transported to the Nicaraguan National Virology Laboratory for processing and storage at -80°C. Paired annual samples (2014-2015 and 2015-2016) demonstrating seroconversion by CHIKV Inhibition ELISA (14) indicated CHIKV infection. ZIKV infection status was confirmed by the 2017 result of the ZIKV NS1 blockade-of-binding assay (15) on paired 2017-2018 annual samples. SARS-CoV-2 infection status was confirmed by the “Mount Sinai ELISA” protocol (16), primarily on 2020 midyear samples. Participants with laboratory-confirmed infections who did not seek medical care were categorized as experiencing subclinical infections. Acute and convalescent samples from participants suspected of chikungunya, Zika, or COVID-19 were tested using molecular, virological, and serological assays (7,8,13). The Appendix (pages 3-4) contains detailed laboratory methods.

### Statistical analyses

We measured the incidence rate, infection risk, and disease risk of each epidemic. Overall values of these metrics were estimated using intercept-only logistic models. The metrics’ values across the study area were estimated with generalized additive models (17) using two-dimensional splines on households’ longitude and latitude, where participants were geolocated. To quantify bias arising from incomplete case ascertainment, Zika case data was disaggregated by whether they were obtainable through active or passive surveillance, the only epidemic where this was possible. The intracluster correlation coefficient was used to measure the intra-household correlation of infection and disease outcomes. We used SaTScan v9.4.4 and Kulldorf’s spatial scan statistic to identify hierarchical and Gini clusters of case incidence, infection risk, and disease risk (18,19). Geostatistical mixed models (20) were used to describe the association of risk factors with infection and disease outcomes. Infection dynamics were estimated by treating cases as a spatiotemporal Poisson point process arising from the total population and then accounting for the spatial distribution of disease risk, assumed to be time-invariant. Initially uninfected participants were considered at risk for infection; infected participants were considered at risk for disease. Analyses used the EPSG:4326 coordinate reference system and were performed in R v3.6.2. The Appendix (pages 5-15) contains detailed statistical methods.

## RESULTS

### Participant characteristics

We refer to the first chikungunya epidemic as *ChikE1*, the second as *ChikE2*, the Zika epidemic as *ZikaE*, and the COVID-19 epidemic as *CovidE*. Our study assessed infection and disease outcomes for 4,884 distinct individuals, including 3,693 unique PDCS participants across ChikE1, ChikE2, and ZikaE. Of the 1,793 HICS participants, 602 children were also enrolled in the PDCS, and 1,192 mostly adult participants were only enrolled in the HICS. Approximately 3,000 PDCS participants were analyzed in ChikE1, ChikE2, and ZikaE (Table 1). These three epidemics occurred in 2014-2016 throughout Managua’s rainy period of June-November (Fig.2), during which an abundance of mosquitoes is observed in the study area. In contrast, CovidE peaked during May-July of 2020.

**Table 1.** Summary and descriptive statistics of infection and disease outcomes across four epidemics in the PDCS and HICS in Managua, Nicaragua^*^

|  | First<br>chikungunya<br>epidemic (ChikE1) | Second chikungunya<br>epidemic (ChikE2) | Zika<br>epidemic<br>(ZikaE) | COVID-19 epidemic<br>(CovidE) |
| --- | --- | --- | --- | --- |
| Cohort | PDCS | PDCS | PDCS | HICS |
| Participant age range | 2-14 | 2-14 | 2-14 | 0-87 |
| Epidemic period | 9/2014 – 2/2015 | 7/2015 – 2/2016 | 1/2016 – 1/2017 | 3/2020 – 10/2020 |
| Primary transmission<br>pathway | *Aedes* mosquitoes | *Aedes* mosquitoes | *Aedes* mosquitoes | Respiratory droplets |
| Number at risk of infection | 3,124 | 2,864 | 3,017 | 1,793 |
| Number of infections (Number<br>at risk of being a case) | 199 | 710 | 1,416 | 1,039 |
| Number of cases | 90 | 416 | 494 | 306 |
| Risk of infection<br>(95% CI)<sup>†</sup> | 6.4%<br>(5.5%, 7.4%) | 24.8%<br>(23.2%, 26.6%) | 47.1%<br>(45.1%, 49.1%) | 57.5%<br>(54.1%, 60.9%) |
| Risk of disease<br>(95% CI)<sup>†</sup> | 45.6%<br>(38.6%, 52.6%) | 58.7%<br>(54.9%, 62.4%) | 35.4%<br>(32.8%, 38.1%) | 28.9%<br>(25.5%, 32.5%) |
| Incidence rate<br>(95% CI)<sup>†</sup> | 2.9%<br>(2.3%, 3.6%) | 14.5%<br>(13.2%, 16.0%) | 16.6%<br>(15.2%, 18.1%) | 17.1%<br>(14.8%, 19.6%) |
| Bias due to treating the<br>incidence rate as the infection<br>risk (%)<sup>‡</sup> | 2.9% – 6.4%<br>= –3.5% | 14.5% – 24.8%<br>= –10.3% | 16.6% – 47.1%<br>= –30.5% | 17.1% – 57.5%<br>= –40.4% |
| Bias due to treating the<br>incidence rate as the disease<br>risk (%)<sup>‡</sup> | 2.9% – 45.6%<br>= –42.7% | 14.5% – 58.7%<br>= –44.2% | 16.6% – 35.4%<br>= –18.8% | 17.1% – 28.9%<br>= –11.8% |
| **ANOVA-based ICC for intra-household correlation of infection risk (95% CI)<sup>§</sup>** | 0.22<br>(0.17, 0.28) | 0.21<br>(0.16, 0.27) | 0.22<br>(0.17, 0.27) | 0.30<br>(0.25, 0.35) |
| **ANOVA-based ICC for intra-household correlation of disease risk (95% CI)<sup>§</sup>** | 0.14<br>(0.00, 0.51) | 0.26<br>(0.09, 0.42) | 0.28<br>(0.18, 0.37) | 0.21<br>(0.15, 0.28) |

\*Abbreviations: ANOVA, analysis of variance; CI, confidence interval; GEE, generalized estimating equations; HICS, Household Influenza Cohort Study; ICC, intraclass correlation coefficient; PDCS, Pediatric Dengue Cohort Study
<sup>†</sup>GEE model estimates are presented.
<sup>‡</sup>Negative values indicate that the incidence rate underestimates the risk of infection and the risk of disease.
<sup>§</sup>Table S5 contains additional information.

**Figure 2.**
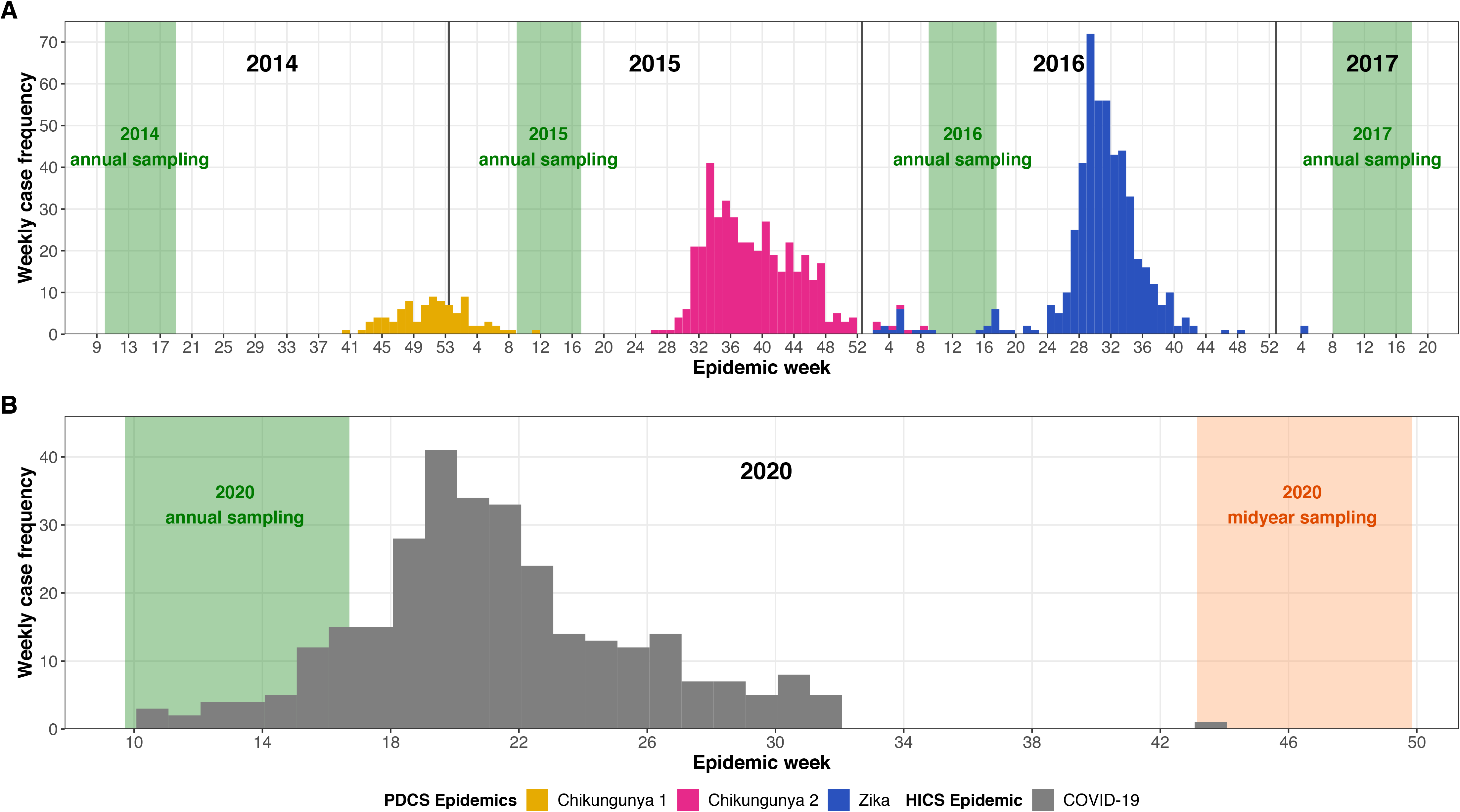
Epidemic curves for four epidemics in Managua, Nicaragua, on a weekly basis. Data for epidemics in the PDCS (A) and HICS (B) are shown. The duration of the annual sampling periods for serosurveillance of infection history is shown in green. The additional 2020 midyear sampling, instituted to capture the first COVID-19 wave, is shown in orange. The epidemic curves for the chikungunya and Zika epidemics reflect case counts that were confirmed by rRT-PCR and a serological algorithm, as detailed in the Appendix. Due to the retrospective collection of illness onset data from some HICS participants, the COVID-19 epidemic curve reflects 1) the date of acute sample collection from rRT-PCR-positive cases, 2) the date of illness onset as reported by ELISA-positive participants, or 3) a randomly selected date from the month in which ELISA-positive participants recalled experiencing illness consistent with COVID-19. The epidemic curves for the PDCS and HICS are purposefully shown in different panels as direct comparisons of case counts between cohorts of different sample sizes can result in misleading inferences.

In the PDCS, the distribution of age and sex was constant across ChikE1, ChikE2, and ZikaE (Table S1, Fig. S1), with approximately 50% of PDCS participants being female. In the HICS, there was an over-enrollment of adult females relative to adult males.

### Summary measures of infection, disease, and case-based incidence

We first examined summary statistics of the four epidemics. ChikE1 exhibited the lowest incidence at 2.9 cases per 100 population (2.9%) but featured higher infection (6.4%) and disease risks (45.6%) (Table 1). ChikE2, ZikaE, and CovidE exhibited similar incidence rates between 14.5-17.1%, but these incidence rates differed substantially from infection and disease risks. ChikE2 had a medium level of infection risk (24.8%) and a high disease risk (58.7%), an inverted pattern from what was observed during CovidE, with high infection risk (57.5%) and medium disease risk (28.9%) (Table 1). In contrast, ZikeE displayed intermediate levels of infection (47.1%) and disease (35.4%) risk. Across epidemics, the case-based incidence rate thus recapitulated neither risk-based metric and often underestimated them considerably (Table 1).

We then assessed summary statistics by sex and age. Sex-based differences for incidence and risk-based measures, even when statistically significant, tended to be small, as when females had an infection risk 6% higher than males during ZikaE (Fig. S2). Similarly, accounting for the over-enrollment of adult females in the HICS had little effect (∼1%) on overall estimates (Fig.S3-S4). In contrast, we observed age-based incidence patterns for all epidemics (Fig S5), which were explained by the underlying and more striking age trends observed for infection and/or disease risks (Fig S6-8). For example, COVID-19 incidence was low across age, particularly during childhood (Fig. S5). However, SARS-CoV-2 infection risk was high across all ages, increasing modestly from ∼48% in infants to ∼62% at age 24 and thereafter plateauing. Despite the relative stability of infection risk by age, disease risk during CovidE increased dramatically from ∼11% in infants to ∼50% at age 70. Thus, the low COVID-19 incidence neither recapitulated age-based risk dynamics nor reflected the greater age-based changes in disease risk as compared to infection risk.

### Mapping infection, disease, and incidence

Next, we mapped the infection risk, disease risk, and case-only incidence rate across our study area. For all epidemics, infection risk varied at small spatial scales (Fig. 3A-D), suggesting that the local environment was an important determinant of infection risk. During ChikE1, ChikE2, and ZikaE, infection risk was elevated in western neighborhoods adjacent to a large cemetery that is heavily infested with *Aedes* mosquitoes during the rainy season (data not shown). Only adjusting Fig. 3A-D for distance to the cemetery appreciably changed the spatial patterns of infection risk, whereas adjusting for age, sex, and household water availability did not (Figs. S9-S14). Conversely, SARS-CoV-2 infection risk was high in eastern neighborhoods that contain large public spaces and commercial attractions (Figs. 3D, S15-S16). Together, these observations imply that infection risk across epidemics was spatially mediated by distinct transmission routes.

**Figure 3.**
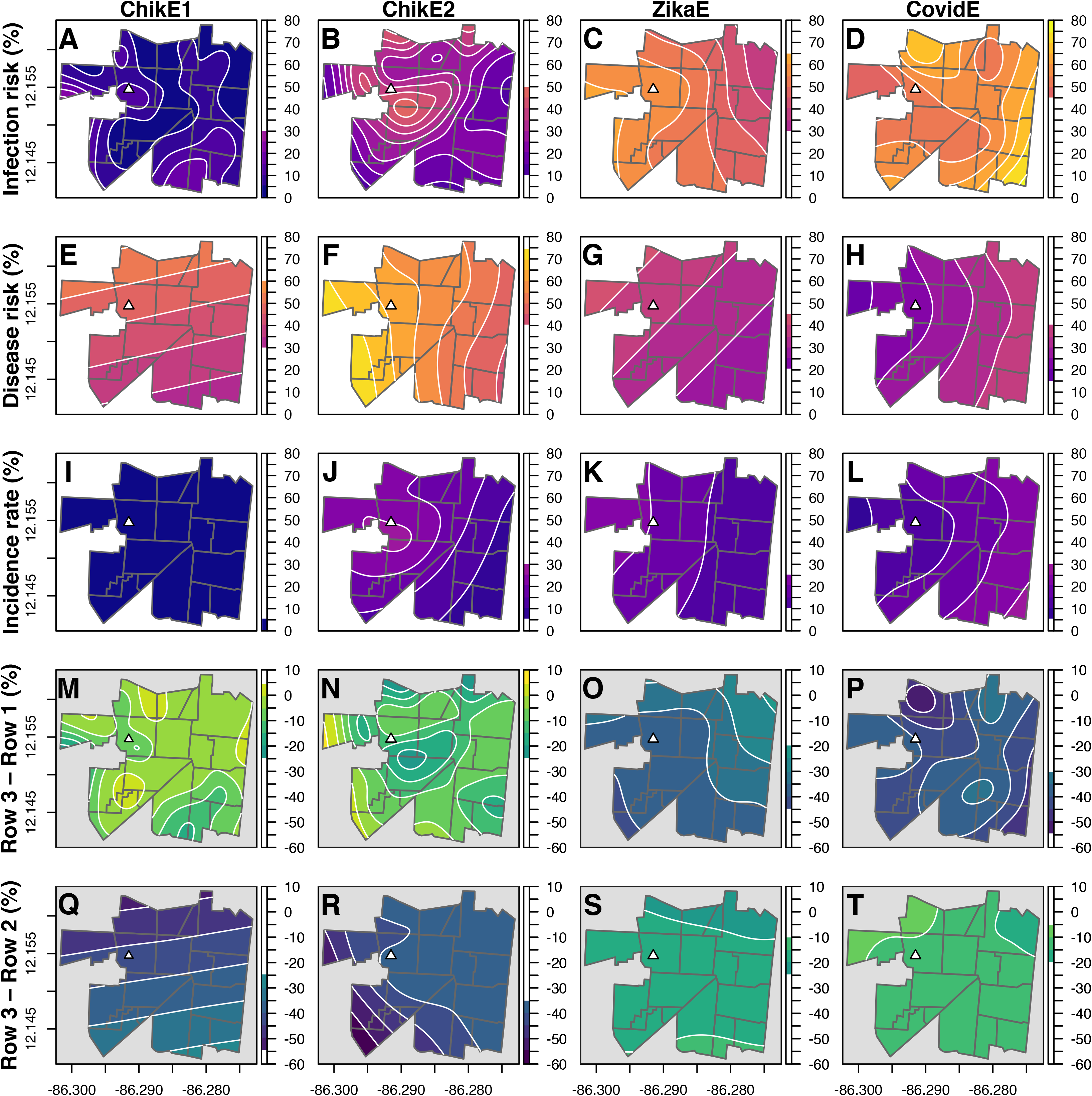
Maps of the infection risk, disease risk, case-based incidence rate, and bias. The infection risk (A-D), disease risk (E-H), and incidence rate (I-L) across four epidemics are shown in one color palette, with warmer colors indicating higher values of the appropriate metric, and are set against a white background. The difference between infection risk and the incidence rate (bias induced by treating the incidence rate as the infection risk) (M-P) and the corresponding bias for the disease risk (Q-T) are shown in another color palette. Bias panels, as they have a different scale, are set against a gray background. Contour lines show changes in infection, disease, and bias metrics corresponding to the scale bar of percentages to the right of each plot. Maps were generated from generalized additive mixed models. A white triangle indicates the study health center. Neighborhoods are outlined in gray. Columns in the figure correspond to the chikungunya epidemics (2014, 2015), Zika epidemic (2016), and COVID-19 epidemic (2020) in Managua, Nicaragua (left to right).

Across all epidemics, disease risk also varied at small spatial scales (Fig. 3E-H). After adjusting for age and sex (Figs S17-24), spatial patterns of disease risk remained non-uniform and distinct from spatial patterns of infection risk. This demonstrates that disease risk can vary spatially and that areas of high infection risk may not have commensurate levels of disease risk.

As the case-based incidence rate is the product of two risks (Eq. 1) with different spatial patterns (Fig. 3A-H), maps of the incidence rate (Fig. 3I-L) underestimated infection and disease risk-based maps and did not recapitulate spatial patterns of either risk. We quantified the bias resulting from treating the incidence rate as infection and disease risk by subtracting incidence maps from risk-based maps (Fig. 3M-T). The average spatial bias for the disease risk was -40.1 and -41.2 percentage points for ChikE1 and ChikE2, respectively; the average spatial bias for the infection risk was -34.9 and -40.3 percentage points for ZikaE and CovidE, respectively. The incidence rate underestimates risk-based metrics, inducing negative biases. Additionally, bias varied substantially across neighborhoods. For example, the *range* of bias for ChikE2 and CovidE infection risks was 31.2 and 19.2 percentage points across the study area. Thus, the inferential bias induced by treating the incidence rate as a risk-based metric was high and spatially heterogeneous across epidemics.

### Cluster detection

We then identified hierarchical and Gini clusters of infection risk, disease risk, and incidence (Fig. 4, Table S2). Each epidemic had ≥1 significant infection or disease cluster. Clusters of elevated infection risk for the larger mosquito-borne epidemics, ChikE2 and ZikaE, encompassed the cemetery and study neighborhoods adjacent to it. Large clusters of diminished infection risk in ChikE1, ChikE2, and ZikaE highlighted areas with excess uninfected persons who remained susceptible to future infection. Such clusters are only identifiable after ascertaining the infection status of a population, regardless of disease presentation. In contrast to the mosquito-borne epidemics, CovidE exhibited small clusters of elevated and diminished infection risk. In general, clusters of infection risk were in different locations and of different sizes than clusters of disease risk, demonstrating that infection and disease risk cluster differently in space. Indeed, we detected no disease risk clusters during both chikungunya epidemics despite finding large clusters of infection risk.

**Figure 4.**
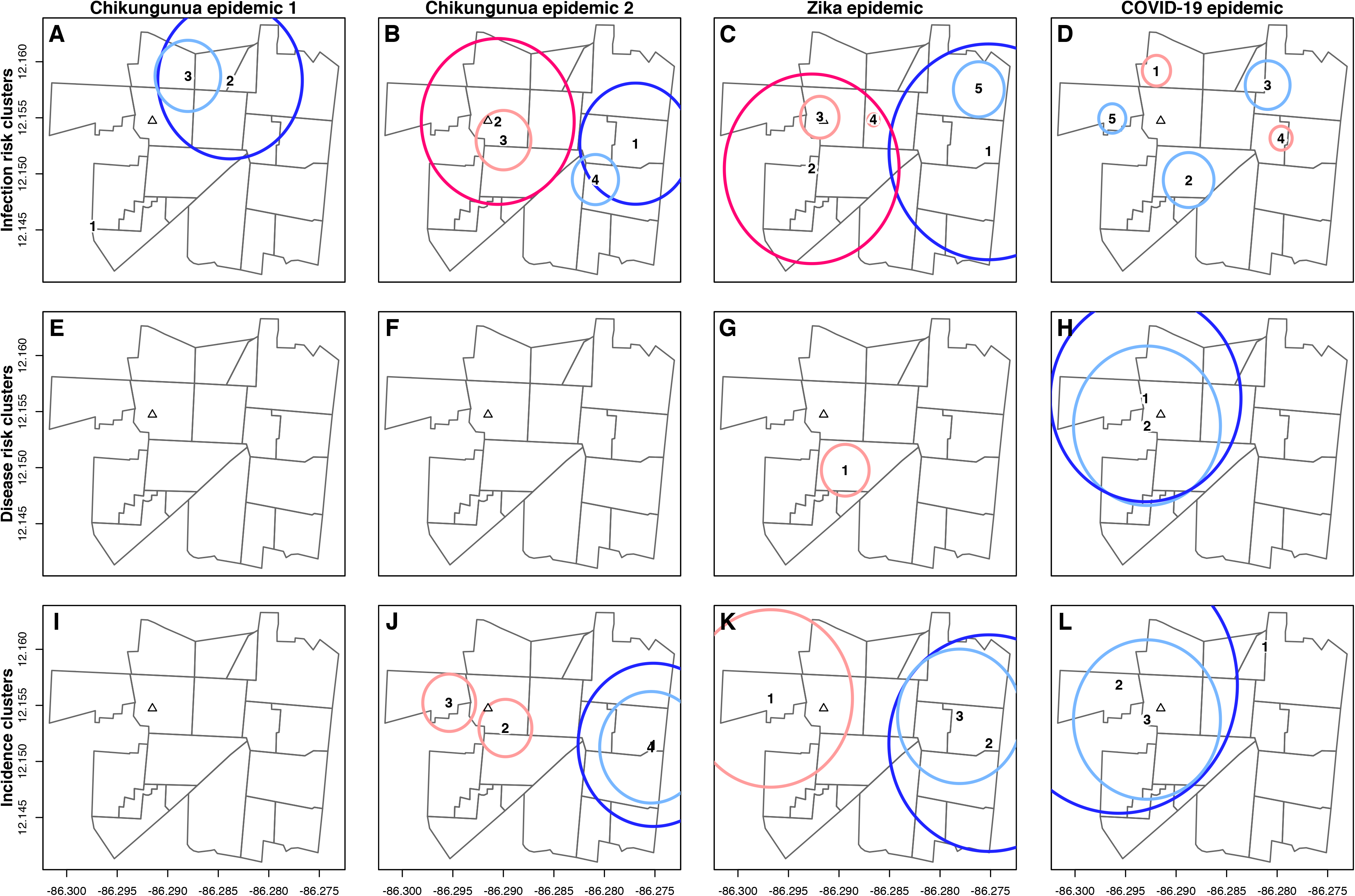
Cluster detection analyses of the infection risk, disease risk, and case-based incidence rate. Clusters of infection risk (A-D), disease risk (E-H), and the incidence rate (I-L) across four epidemics are shown. Panels depict the results of Kulldorf’s spatial scan statistic conducted in SaTScan. Hierarchical clusters are shown in dark colors; Gini clusters are shown in light colors. Hierarchical clusters identify the most statistically likely clusters; Gini clusters maximize outcome rates. Hotspots are shown in pink; coldspots are shown in blue. Cluster centers are numerically labeled. Arrows show the kind of risk clusters that incidence clusters resemble. A white triangle indicates the study health center. Neighborhoods are outlined in gray. Columns in the figure correspond to the chikungunya epidemics (2014, 2015), Zika epidemic (2016), and COVID-19 epidemic (2020) in Managua, Nicaragua (left to right). Table S2 contains additional information for this analysis.

Standard incidence clusters, which identify areas of elevated or diminished case counts among the total population, sometimes missed large risk-based clusters (Fig. 4). More surprisingly, incidence clusters resembled infection risk clusters only for ChikE2 and CovidE, whereas they resembled disease risk clusters for ChikE1 and ZikaE. Thus, incidence clusters failed to display a reproducible pattern, inconsistently resembling either infection or disease risk clusters for a given epidemic.

### Geostatistical modeling

We next conducted geostatistical multivariable modeling. We first describe model-based inferences for correlated outcomes within households and across space. Surprisingly, analyses that did and did not account for household-based correlation yielded very similar results for all epidemics, suggesting that participants’ infection and disease outcomes were poorly correlated within homes (Tables S3-S4). This observation was directly confirmed by low values of the intracluster correlation coefficient (Tables 1, S5). We further observed that infection risk did not scale with household size (Fig. S25). Altogether, the data demonstrated that our participants’ infection and disease outcomes were weakly correlated within households across epidemics.

Likewise, the similarity of estimates from models that did and did not account for spatial autocorrelation (Tables S3-S4) suggested that infection and disease outcomes were only spatially correlated across short distances. This observation was confirmed by estimated Matérn correlation functions (Fig. S26) that demonstrated that infection and disease outcomes were spatially correlated across short distances (<200m) for all epidemics, strengthening earlier findings (Fig. 3) regarding the importance of the local spatial environment.

Indeed, we observed that distance to the cemetery was significantly associated with ZIKV *infection*, such that the odds of ZIKV infection among participants living 1 km from the cemetery were 0.63 (95% CI: 0.55, 0.73) times that of participants living next to the cemetery, conditional on age, sex, and indoor water availability; a similar 1-km odds ratio was observed during ChikE2 (Table S3). However, using geostatistical models, we did not identify variables that were consistently related to *disease* risk across epidemics (Table S4). Rather, model results were epidemic-specific.

### Spatiotemporal dynamics

Spatiotemporal analyses depict epidemic progression across time and space. By harnessing Eq. 1, we estimated the spatiotemporal dynamics of infection risk (Fig. 5A-D), which were substantially underestimated by the less dynamic standard spatiotemporal patterns of case incidence (Fig. 5E-H). Each of the mosquito-borne epidemics featured elevated infection risk around cemetery-adjacent neighborhoods for ≥2 months, with particularly high infection risk during ZikaE. In contrast, cemetery-adjacent neighborhoods were never the focal point of SARS-CoV-2 infection risk. Because CovidE featured the lowest disease risk (Table 1), its spatiotemporal *infection* dynamics differed most from its *incidence* dynamics, underscoring the substantial differences of risk-based mapping compared to case-based mapping.

**Figure 5.**
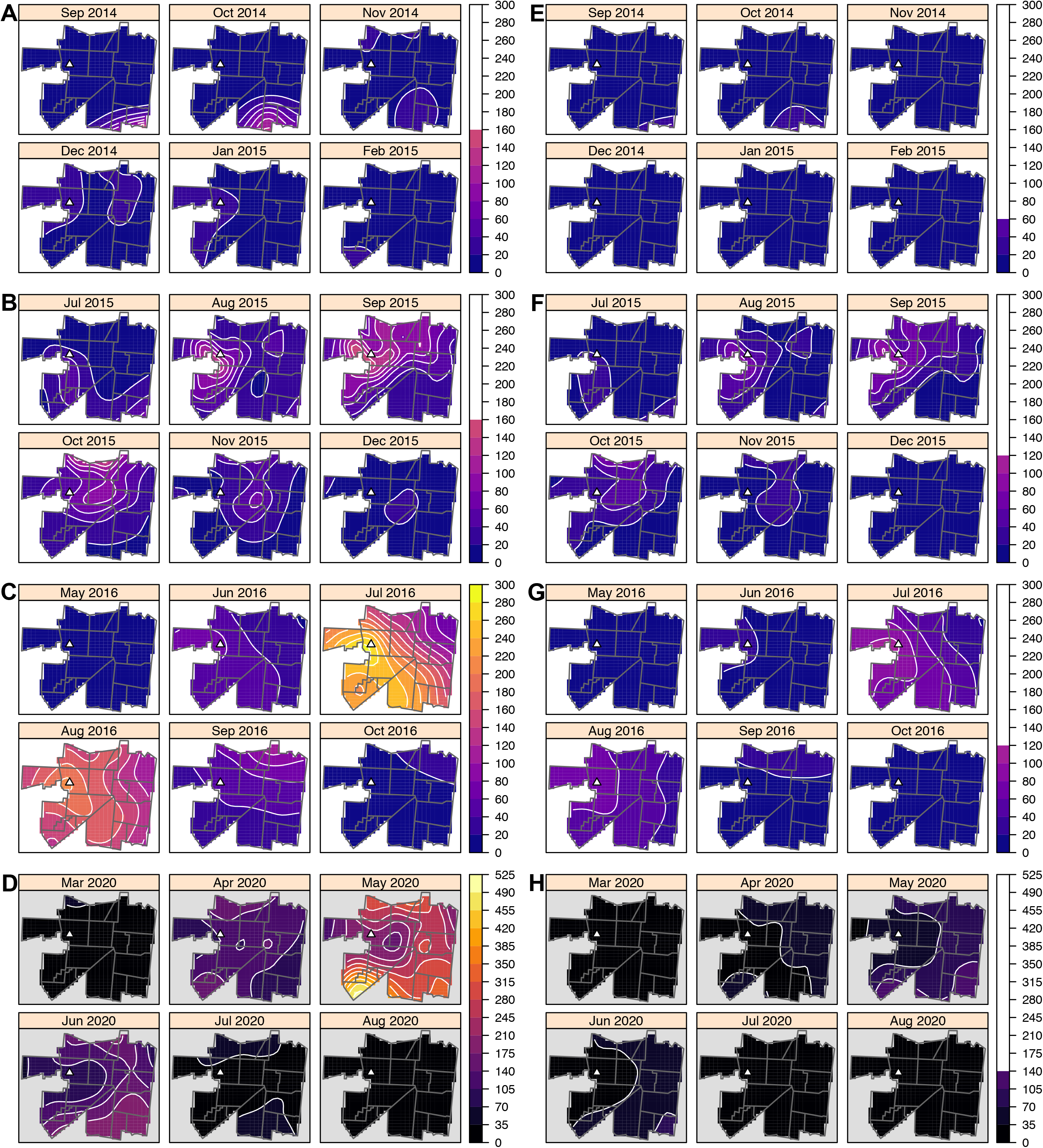
Spatiotemporal dynamics across four epidemics in our study area. Model predictions of the infection risk (A-D, first column) and incidence rate (E-H, second column) are reported per-month and per-1,000 population. Due to space constraints, data for months with few cases are not shown. The PDCS epidemics (ChikE1, ChikE2, and ZikaE) are shown in a different color palette than CovidE as the range of the infection dynamics for CovidE is so much higher than that of the PDCS epidemics. Contour lines show changes in infection and incidence metrics corresponding to the scale bar of percentages to the right of each plot. A white triangle indicates the study health center. Neighborhoods are outlined in gray. Rows in the figure correspond to the chikungunya epidemics (2014, 2015), Zika epidemic (2016), and COVID-19 epidemic (2020) in Managua, Nicaragua (top to bottom).

### Active versus passive surveillance

We quantified case ascertainment bias spatially by complementing the ZikaE serosurvey with either all Zika cases or only cases captured by passive surveillance. Compared to our active surveillance, using passive surveillance altered the clinical profile of captured Zika cases (8) and decreased the case count, thereby increasing the number of subclinical ZIKV infections. The infection risk was unbiased under passive surveillance as its calculation only required serosurvey data; however, estimates of the disease risk and incidence rate were biased (Table 2). The bias from passive surveillance is conceptually and numerically distinct from that induced by treating the incidence rate as a risk. However, these two biases synergized when the incidence rate, estimated from passive surveillance data, was interpreted as a risk. For example, inferring the true disease risk from the incidence rate induced -4.9 percentage points of bias from incorrect inference *and* -26.1 percentage points of bias from incomplete case ascertainment (Table 2). Importantly, this compounded bias would be present irrespective of conducting a serosurvey (Table 2). Moreover, whether biases arose from misinterpretation, incomplete case data, or both, they tended to be high and spatially heterogenous (Figure 6). Thus, inferring risk from passive surveillance data was prone to multiple biases with different spatial patterns.

**Table 2.**
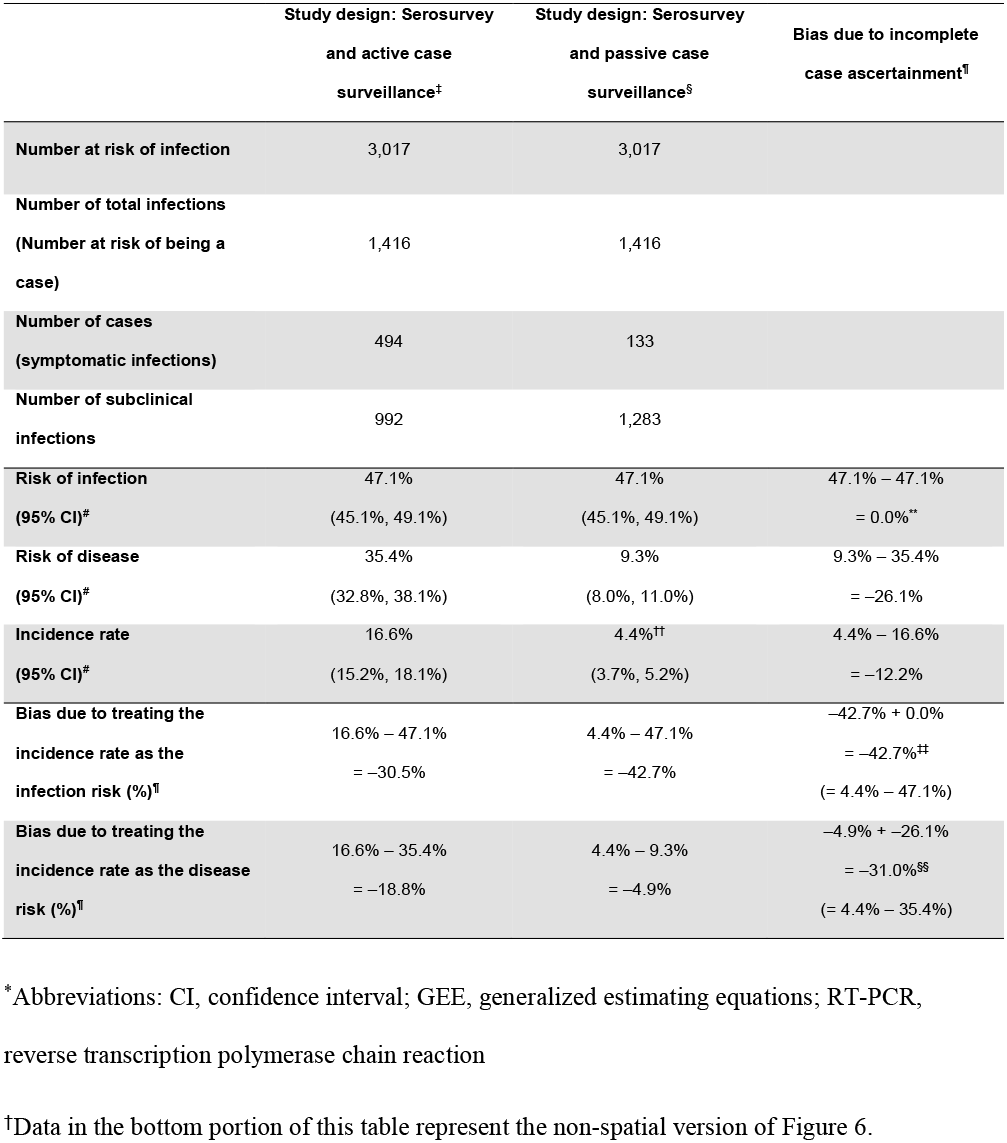
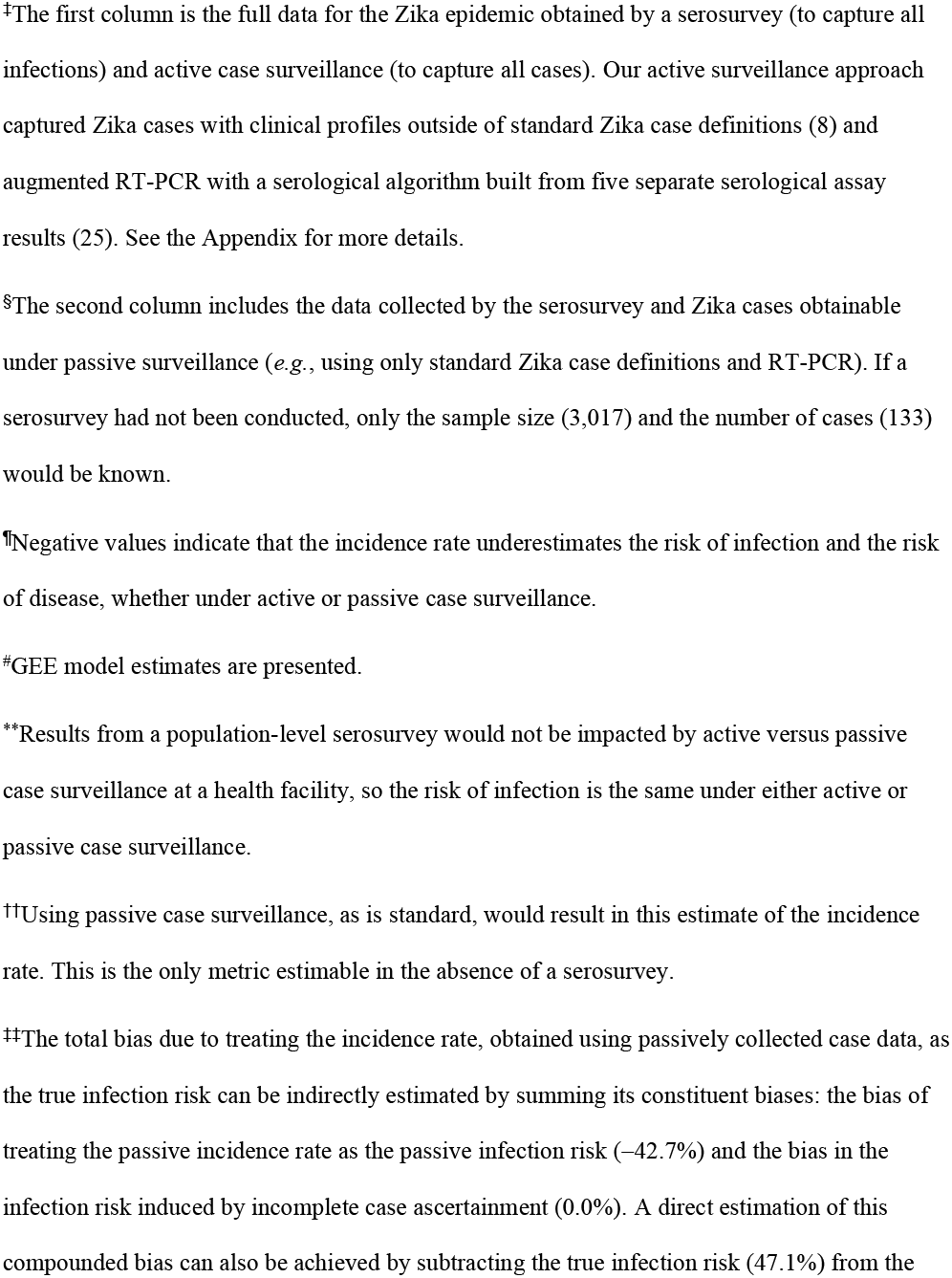
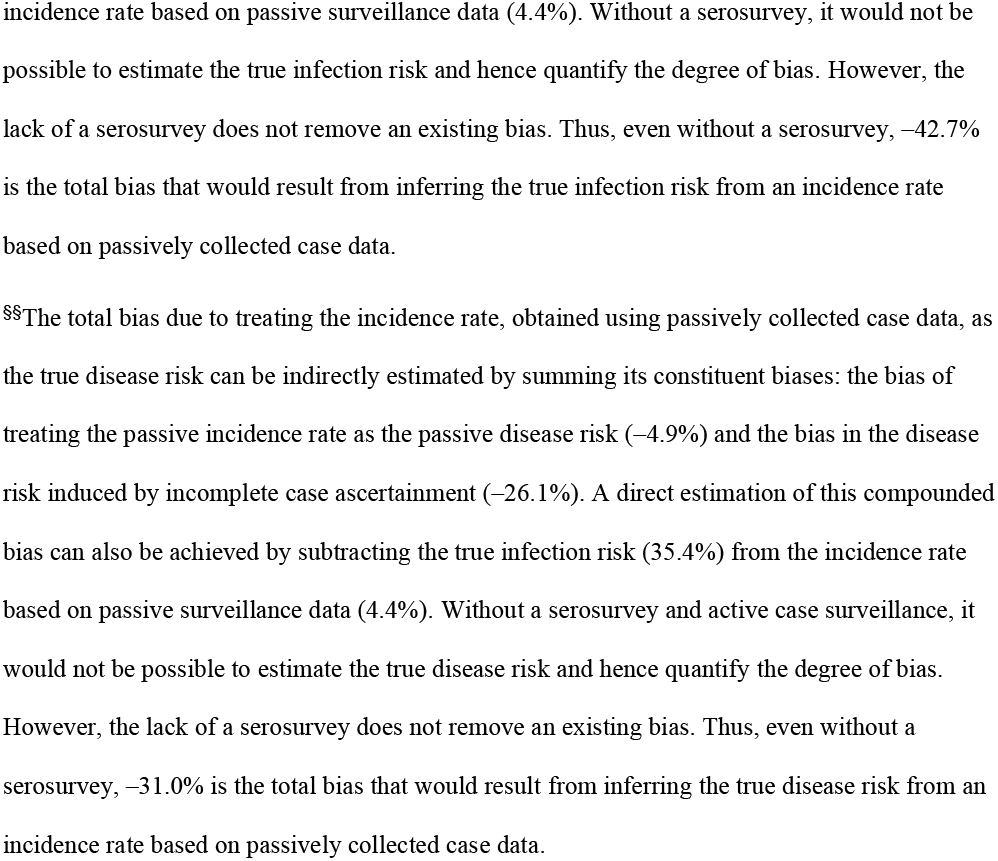
Sample size, infection, disease, incidence, and bias metrics from a serosurvey augmented by cases identifiable by either active or passive surveillance for the 2016 Zika epidemic in the Pediatric Dengue Cohort Study^*,†^

**Figure 6.**
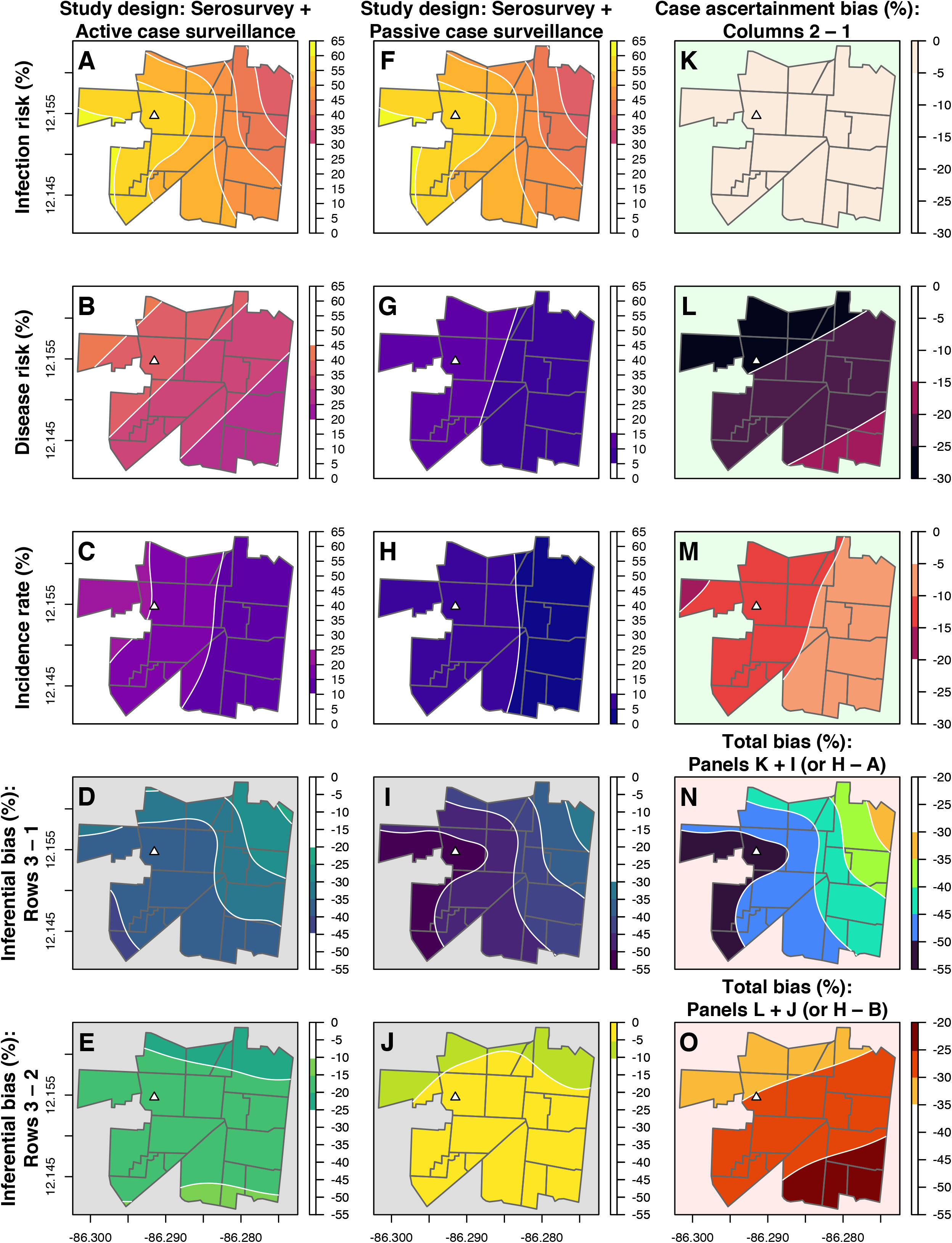
Comparisons of infection, disease, incidence, and bias metrics for the 2016 Zika epidemic in the PDCS using passive and active case surveillance. Panels in this figure are displayed in the same sequence as, and represent the spatial version of, data in the bottom portion of Table 2. Columns in this figure correspond to a study design using a serosurvey and active case surveillance (column 1), a study design using a serosurvey and passive case surveillance (column 2), and the bias induced by passive versus active case surveillance (column 3). Column 1 repeats the data shown in Figure 3, column 3 for the sake of comparing the full data to that obtained under passive case surveillance. Maps of the infection risk (A, F), disease risk (B, G), and the incidence rate (C, H) are shown under active and passive case surveillance in the first color palette and are distinguished by a white map background. The bias induced by active versus passive case surveillance for these three metrics (K-M) is shown in a second color palette distinguished by a green map background. The bias induced by treating the incidence rate as the infection risk (D, I) and the incidence rate as the disease risk (E, J) is shown in a third color palette distinguished by a grey map background. The total bias incurred from incomplete case ascertainment and inferring a risk from the incidence rate (N-O) is shown in a fourth color palette distinguished by a pink map background. Contour lines show changes in infection, disease, incidence, and bias metrics corresponding to the scale bar of percentages to the right of each plot. Maps were generated from generalized additive mixed models. A white triangle indicates the study health center. Neighborhoods are outlined in gray.

## DISCUSSION

Across multiple analyses and four epidemics of three viruses in two cohorts, we showed that the traditional case-based incidence rate considerably underestimated infection and disease risks, broadly impacting how epidemics were characterized. We further demonstrated that case-based analyses did not recover either the magnitude or spatial pattern of infection risk, which critically conveys the landscape of natural immunity. In general, we observed that case-based incidence had more limitations than traditionally assumed. For example, although ChikE2, ZikaE, and CovidE had comparable incidence rates, their underlying infection and disease risks were very different. Similarly, case-based incidence clusters inconsistently captured different risks across epidemics, an observation not apparent without analyzing multiple epidemics in parallel. Together, our results demonstrate how complex, epidemic-specific spatial patterns of infection and disease risk, critical for the design of effective interventions, can be obscured and underestimated by relying solely on case-based analyses. Importantly, this underestimation was distinct from bias due to incomplete case ascertainment, suggesting that the inferential biases we quantify for the incidence rate are exacerbated in typical settings with limited active surveillance and laboratory testing capacity.

Paradoxically, the limitations of the incidence rate are obvious yet underappreciated. It is well-known that incidence estimates based on incomplete case data are underestimated. Here, we showed that a separate bias, with its own spatial pattern, arises when the incidence rate is misinterpreted as conveying infection or disease risks, and we quantified the extent to which this biased estimate deviates from more accurate estimates of infection and disease risk. Correctly interpreting measures of epidemic impact is important for policy decisions. While interventions will vary depending on the pathogen and available countermeasures, areas prone to high *infection* risk generally require interventions that limit transmission (*e*.*g*., mosquito control, masking, and social distancing), whereas areas prone to high *disease* risk require interventions that limit disease occurrence and boost access to care.

Incidentally, if the disease risk were spatially uniform, as some studies have assumed (21), then the spatial pattern of incidence would equal that of the infection risk and the degree of underestimation (and hence bias) would be similar across a given area. However, disease risk was not spatially uniform across epidemics, and its bias also varied spatially. Thus, just as others have found that disease risk can vary across populations (6,22), we find that disease risk can vary within a single population.

The case-based incidence rate is the disease risk when all individuals are susceptible to an outcome (*e*.*g*., cardiovascular disease, death). However, for pathogens that cause subclinical infections, incidence rate maps only convey *where disease occurred*, not the *spatial risk* of infection or disease. Many pathogens of global health importance give rise to substantial quantities of subclinical infections (including *Plasmodium*; *Mycobacterium tuberculosis*; and many pathogens transmitted by sex, air, vectors, and soil). Thus, our findings concerning the limitations of case-based spatial mapping likely generalize to many infectious diseases that disproportionately affect neglected populations.

The pediatric nature of the PDCS precluded spatially analyzing adults in the catchment area of the study health center during ChikE1, ChikE2, and ZikaE. However, previous analyses compared ZIKV infection risk for children and adults in this area (9). The two groups’ spatial patterns were comparable, although ZIKV infection risk was higher among adults. Thus, analyses of the adult population during ChikE1, ChikE2, and ZikaE would likely reveal similar spatial trends as those in PDCS participants.

We found little evidence that infections were clustered within households. Lacking entomological data, our analyses indirectly suggested that viral transmission infrequently occurred within study households. However, this suggestion is directly supported by a study of full-length sequencing of ZIKV genomes in our cohort (23), which found that many households had Zika cases whose most recently sampled viral ancestral strains derived from different households. Together, the evidence suggests that non-household transmission played an important role in the epidemics we assessed.

The geographic extent of our study is small. However, capturing all infections and cases, and hence accurately measuring bias, is only cost-feasible in constrained geographical areas. Spatial studies with incomplete infection and case data, whether small or large, may be subject to inferential and case ascertainment biases despite being unable to measure such biases. Ascertaining infection status and exhancing case surveillance, where possible, may help to mitigate and correct for such biases.

Measuring a population’s infection status has many additional benefits, especially in directing infection control interventions to areas of high transmission. Conversely, knowledge of areas with a high proportion of uninfected individuals is also critical for advancing public health goals, such as prioritizing these areas for epidemic-preventive measures (*e*.*g*., vaccine rollout in areas with low SARS-CoV-2 transmission). Others have shown how combining regional serosurvey data with real-time hospitalization data can estimate infection risk in near real-time at larger spatial scales, thereby improving critical estimates for decision-makers (24). As epidemic management necessitates evaluating the risks of infection and disease across space, our data supports the expanded use of serosurveys to overcome the inherit limitations of case-based spatial measures.

## Data Availability

Individual-level data may be shared with outside investigators following UC Berkeley and UM Ann Arbor IRB approval. Data has been deposited in a HIPAA-compliant Dropbox account hosted by the University of Michigan and will be available by request, as is required by the IRB-approved protocols for the PDCS and the HICS. Please contact the UC Berkeley Center for the Protection of Human Subjects, Eva Harris, the University of Michigan Health Sciences and Behavioral Sciences IRB, and Aubree Gordon to arrange for data access. Databases without names and other identifiable information were used for all analyses. Collaborating research groups and institutions will be sent coded data with all personal identifiers unlinked as well as data dictionaries and the accompanying R code.

## ACKNOWLEDGMENTS

We are extremely appreciative of our dedicated study team at the Centro de Salud Sócrates Flores Vivas and the Laboratorio Nacional de Virología at the Centro Nacional de Diagnóstico y Referencia, Nicaraguan Ministry of Health; and the Sustainable Sciences Institute in Nicaragua. We are grateful to Art Reingold and Tulika Singh for their thoughtful reviews of the manuscript, and we thank Burke Bundy and Suzanne Default at the University of California, Berkeley, for enabling us to use the computer cluster of the Division of Epidemiology and Biostatistics for our geostatistical modeling. We thank François Rousset for expert consultation regarding spatial generalized linear mixed models and their implementation in the spaMM R package. Most importantly, we thank the PDCS and HICS study participants and their families for engaging with us in the endeavor of science. This study was supported by grants R01 AI099631 (AB), P01 AI106695 (EH), R01 AI120997 (AG), and U19 AI118610 (EH) from the National Institute of Allergy and Infectious Diseases of the National Institutes of Health; the National Institutes of Health Centers of Excellence for Influenza Research and Surveillance [contract: HHS 272201400006C (AG)]; and the Open Philanthropy Project Fund for the production of recombinant SARS-CoV-2 spike protein, its receptor binding domain, and antibodies at the University of Michigan Center for Structural Biology. FBC was partially supported by a supplement to grant P01 AI106695.

## ABOUT THE AUTHOR

Dr. Fausto Andres Bustos Carrillo is an epidemiologist in Washington, DC, collaborating with the National Institute of Allergy and Infectious Diseases as an Emerging Leader in Data Science Fellow. His primary research interests are the epidemiological, clinical, and spatial aspects of explosive epidemics caused by SARS-CoV-2, Zika virus, chikungunya virus, dengue virus, and influenza virus, which were the topics of his doctoral dissertation at the University of California, Berkeley.

