## Appendix for "Epidemics of chikungunya, Zika, and COVID-19 reveal bias in case-based mapping"

### Supplementary materials and methods

#### Study design

*Study setting and cohort establishment procedures*

The catchment area of the study health center, the Health Center Sócrates Flores Visas (HCSFV), consists of 18 neighborhoods; however, neither Pediatric Dengue Cohort Study (PDCS) participants nor Household Influenza Cohort Study (HICS) participants resided in the smallest neighborhood, Mantica, during the study period. The PDCS was originally designed to study dengue virus (DENV) infection and disease, but was expanded to include CHIKV and ZIKV when they were introduced into the Americas but before they entered the study area.[1–4] Chikungunya and Zika were first detected in Managua in August 2014 and January 2016, respectively. Chikungunya and Zika were first detected in the study area in September 2014 and January 2016, respectively. The HICS was established in 2017 to study influenza virus and other respiratory pathogens. It was expanded to include severe acute respiratory syndrome coronavirus 2 (SARS-CoV-2) in 2020 shortly before the first case of coronavirus disease 2019 (COVID-19) was detected in Managua (in March 2020) and in the study area (in March 2020). The population served by the HCSFV is approximately 62,000 persons.[5] The catchment area of the HCSFV and hence the study area is approximately 5 $km^2$ and is approximately 3 km long at its widest. The 18 neighborhoods that constitute the study area reside in District II of Managua, Nicaragua. The study area is low-lying, located below Lago Xolotlán (Lake Managua), and quite flat; altitude data from the R raster package[6] indicates that the range of elevation for the study area is 42-106 meters above sea level.

In terms of sanitation and water services, about 88% of PDCS households have garbage collection services.[5] Approximately 95% of PDCS homes have sewage systems and tap water, although tap water may not be potable or available for all hours of the day. In Managua, the Ministry of Health sometimes treats water containers in households of arboviral cases with Temephos, an organophosphate insecticide that eliminates *Aedes* larvae and pupae. However, *Aedes* resistance to Temephos is widespread, increasing, and well documented.[7] While the Ministry of Health recommends that Temephos be used for two months after the initial application, 46% of households in our study area discard Temephos within two weeks, rising to 82% within one month.[8] A randomized cluster trial in Managua and three Mexican sites showed that households with Temephos had significantly higher levels of DENV infections, likely because members of such households felt a false sense of security and were thus significantly less likely to participate in household activities to eliminate potential mosquito breeding containers or in community-based environmental management strategies, including clean-up campaigns that targeted vacant spaces, ravines, streets, and public premises such as cemeteries.[9]

As previously published,[10] participants were initially recruited into the PDCS during door-to-door visits of households served by the HCSFV in 2004, during which eligible children were invited to participate. In addition to parental and participant consent, eligibility criteria included the following: aged between 2 and 9 years old, living in the catchment area of the HCSFV, no plans to leave the catchment area within a period of 3 years, attending the HCSFV for all medical needs, and the lack of immune-compromising medical conditions. Since that time, the PDCS has been expanded to include children between the ages of 2 and 14. Recruitment into the PDCS occurs every year to ensure the age structure remains constant, as detailed below.

In 2007, a pediatric influenza cohort study[11] that only had PDCS participants was established. In 2011, the Nicaraguan Pediatric Influenza Cohort Study (NPICS)[12] was established from a random selection of participants in the earlier influenza cohort study.[11] A random selection of NPICS households that had children aged 12 or younger were then invited to participate in the HICS, which was initiated in 2017. Thus, the vast majority of HICS households are also PDCS households, all of which are serviced by the HCSFV and the same study personnel.[12]

*Annual sampling procedures and demographic/household survey data*

During March and April of each year, healthy participants in the PDCS and HICS visit the HCSFV to provide a blood sample.[10,13] In October and November 2020, both studies conducted a mid-year sampling to obtain additional samples from study participants. In the HICS, the interval between the 2020 annual and midyear samplings corresponded to the first wave of the COVID-19 epidemic (CovidE) experienced by participants in the study area. These blood samples were used to ascertain SARS-CoV-2 infection status in the HICS. PDCS and HICS families agree to bring study participants to the study health center at the first indication of any illness. PDCS participants with undifferentiated febrile illness or with suspected dengue, chikungunya, or Zika provide acute and convalescent blood samples during clinical visits.[1] HICS participants exhibiting febrile illnesses with respiratory signs and symptoms, as well as HICS participants with signs and symptoms consistent with a lower respiratory illness provide respiratory samples for testing. Participants who are ill at the time of the annual sample are instructed to return at least 3 days later to give their annual sample. If a participant has not returned within a month and a half of their scheduled date, attempts are made to conduct home visits to the participant's household to collect the annual sample. Due to our long partnership with the PDCS study population, the annual sampling is very successful: During the 2020 annual sampling, which was conducted as the COVID-19 pandemic began and featured an expanded sample size of 3,950 participants, 93.36% of the PDCS participated during the annual sampling, a slight decrease from the average participate rate of 94%. Only 98 participants (2.5%) provided a sample after the formal period of the 2020 annual sampling concluded.

During the annual sampling, surveys are administered orally to PDCS and HICS participants as well as their family members.[10,12] The answers are recorded on tablet computers and smartphones.[10,14] Age, sex, level of education, and other demographic/anthropometric information is collected on participants' questionnaires. A household questionnaire is used to collect information on assets and conditions of the household, such as the number of cars and motorcycles as well as the construction material of the walls and roof. The household questionnaire also asks participants to list the average number of hours they lack indoor access to tap hours. Participants can list any whole number between 0-24 for this question.

Two-year-olds are primarily recruited into the PDCS during the annual sampling. New participants aged +3 years are recruited into the PDCS every year to maintain the age structure of the cohort. The recruitment period of new enrollees may overlap with the initial period of an epidemic. During the last satisfaction survey we conducted of the entire cohort, 96% of participants rated the medical attention received at the HCSFV as either excellent or very good.[10] In yearly participant surveys, an average of 2% of participants reported attending a health care provider other than the HCSFV, and an average of 3% of participants reported not seeking any medical attention during an acute illness.[10]

At the time the NPICS (the cohort from which HICS households were sampled) was established, children under 2 years of age were recruited into the study by house-to-house visits of the study area.[12] Infants born to mothers enrolled in the HICS are enrolled into the HICS on a monthly basis, as are any other individual that joins the households. HICS does not intentionally seek to enroll new households. However, new households are added to the HICS when participants move and form new households.

*Inclusion and exclusion criteria*

Inclusion and exclusion criteria were determined in advance. All PDCS and HICS participants who were at risk of initial infection due to CHIKV, ZIKV, and SARS-CoV-2 during the chikungunya, Zika, and COVID-19 epidemics were eligible to participate in this study. Individuals at risk of initial infection from chikungunya virus (CHIKV), Zika virus (ZIKV), SARS-CoV-2 were defined as those participants who had not yet been infected at the beginning of a particular epidemic period. Exclusion criteria were enforced to minimize epidemiological biases and to ensure that spatial analyses could be conducted (*e.g.*, households without GPS points were excluded as no spatial analyses could be conducted on these household). In particular, we excluded participants from this analysis who were enrolled after the start of a given epidemic (including babies born to mothers in the HICS who were themselves enrolled into the HICS upon birth) because, although their infection status could be serologically determined after the epidemic (with paired pre- and post-epidemic annual samples), they were not under disease surveillance between the start of the epidemic and their enrollment date. As a result, participants who were enrolled after an epidemic started and who had an acute illness of either chikungunya, Zika, or COVID-19 before their enrollment date would have been incorrectly recorded as having experienced subclinical (clinically inapparent) infections. The bias toward subclinical infections among this set of participants led us to exclude them. This criterion was the major reason for the exclusion of participants; it also led to the analysis of a closed cohort of initially uninfected participants who subsequently experienced an epidemic, even though both the PDCS and the HICS are open cohort studies.

For example, there are 3,808 PDCS participants for whom blockade-of-binding enzyme-linked immunosorbent assay (BOB ELISA) ZIKV infection data are available for analysis of the Zika epidemic (ZikaE). Of those, 39 were dropped because they were not under disease surveillance during any point between January 1, 2016 and January 31, 2017 [a time period that covers the full duration of the Zika epidemic[1]]. Ten participants were dropped for lack of global positioning system (GPS) points (which were necessary for all spatial analyses). An additional 22 individuals were dropped for having moved outside the catchment area of the HCSFV since enrollment into the study, due to the eligibility criteria of the PDCS.[10] Of the remaining 3,737 participants, 720 participants were dropped for being enrolled after the Zika epidemic started, resulting in a total of 3,017 PDCS participants who were at-risk of ZIKV infection at the beginning of the epidemic, under disease surveillance at that time, and for whom infection and GPS data were available to analyze for the present study.

The application of these criteria resulted in a similar number of exclusions for PDCS participants during the first chikungunya epidemic (ChikE1) and the second chikungunya epidemic (ChikE2). The at-risk population for ChikE2 (n=2,864) is the smallest among the three PDCS study populations we considered because all eligible PDCS participants who were infected during ChikE1 were no longer at risk of incident CHIKV infection during ChikE2; they were consequently excluded from analysis of the ChikE2.

Between the beginning of the 2016 annual sampling and the end of the 2017 annual sampling, we detected five new CHIKV infections. However, because these five CHIKV infections that occurred in 2016 numbered too few for spatial analyses, they were also excluded from the present study.

**Laboratory methods**

*Laboratory testing procedures*

Annual PDCS samples that are collected every March and April serve as the basis for testing by CHIKV Inhibition ELISA (iELISA)[15] and ZIKV NS1 blockade-of-binding (BOB) ELISA,[16,17] which primarily determined infection status of PDCS participants in this study. Testing of annual and midyear 2020 samples from HICS participants was facilitated by adapting the "Mount Sinai ELISA" protocol,[18] which served to primarily determine the SARS-CoV-2 infection status of HICS participants. Almost all samples were tested at the National Virology Laboratory; however, a small number of SARS-CoV-2 ELISAs on samples collected in 2019 or March 2020 were run at the University of Michigan, Ann Arbor. The is no immunological cross-reactivity between the assays we used to detect CHIKV and ZIKV infection because CHIKV is an alphavirus and ZIKV is a flavivirus. The SARS-CoV-2 ELISA has been tested and found not to detect seasonal coronaviruses circulating in Managua.

As is standard practice for detecting DENV infections in the PDCS, we used paired CHIKV iELISAs,[15] which detect total immunoglobulin antibodies, to measure infection status on annual samples (2014-2015 and 2015-2016). Due to performing the iELISAs in pairs to measure pre- and post-epidemic infection status in the PDCS, each sample is run twice by iELISA: first as the post-epidemic sample (as occurred for the 2015 sample in the 2014-2015 comparison) and then as the pre-epidemic sample in the following year (as in the 2015-2016 comparison). Participants who exhibited seroconversion, a negative result on the first ELISA followed by a positive result on the second, were categorized as having experienced a CHIKV infection.

As infection with DENV and ZIKV produce cross-reactive antibodies, and dengue is endemic in Nicaragua, ZIKV infection status was predominantly confirmed by the ZIKV NS1 BOB assay, a competition ELISA.[16] The BOB ELISA can accurately detect the presence of anti-ZIKV antibodies even in persons with existing antibodies to DENV,[16,17] a closely related flavivirus. Positivity in the ZIKV BOB ELISA is determined by a percentage of blockade-of-binding value that meets or exceeds 50%. The 2016-2017 paired samples were originally processed using the biotinylated version of the ZIKV BOB ELISA.[3] Since then, the (first generation) biotinylated BOB ELISA has been surpassed by the (second generation) HRP35 version. To ensure that the ZIKV infection variable reflects the best laboratory method available, ZIKV infection status was primarily determined by the 2017 result on the 2017-2018 paired annual sample, which was conducted using the HRP35 version of the BOB. The infection status of eligible PDCS participants who lacked a 2018 sample, and hence did not have an HRP35 BOB result for the 2017 sample, was determined by the pre-existing result of the biotinylated BOB ELISA for the 2017 sample.

The ELISA used on HICS samples detects anti-SARS-CoV-2 IgG antibodies. SARS-CoV-2 receptor binding domain protein was used to screen samples and measure antibody titers. Receptor binding domain antigens were produced at the University of Michigan, Ann Arbor. Optical densities above 0.38 defined ELISA-positive samples. SARS-CoV-2 ELISAs were run on the midyear 2020 blood sample for HICS participants. (See Figure S2 for an overview of the annual and midyear 2020 samples in relation to the COVID-19 epidemic in the HICS.) If the midyear 2020 sample was positive, the annual 2020 blood sample was processed by SARS-CoV-2 ELISA. If seroconversion or a $\geq$4-fold increase in titers levels was observed, the individual was considered ELISA-positive and hence SARS-CoV-2-infected. Some participants did not provide an annual 2020 blood samples but were positive on the midyear 2020 samples. These few individuals were considered ELISA-positive and hence SARS-CoV-2-infected.

Suspected chikungunya cases (symptomatic infections) were confirmed by 1) a multiplex pan-DENV and CHIKV real-time RT-PCR (rRT-PCR) on acute blood samples,[19] 2) seroconversion using a CHIKV IgM ELISA on paired acute and convalescent samples,[15] and/or 3) seroconversion with a $\geq$4-fold increase in titers levels on paired acute and convalescent samples as measured by a CHIKV iELISA.[5,15]

Suspected Zika cases were confirmed by either rRT-PCR in acute blood and/or urine samples or a serological algorithm[1,20] based on acute and convalescent serum samples measured by the ZIKV and DENV iELISAs, the IgM-capture ELISAs, and the ZIKV NS1 BOB ELISA. The algorithm was the product of recursive partitioning of classification trees and was cross-validated.[20] Confirmed cases were considered to be infected, regardless of their ELISA result. We previously estimated the ZIKV infection risk to be 36% in the PDCS population.[3] In this work, we revise this estimate to 47%, primarily due to an improved second-generation ZIKV NS1 BOB ELISA and the use of a multi-assay algorithm that captured rRT-PCR-negative, serology-positive Zika cases.[1,20]

In October and November of 2020, HICS participants were asked if they had experienced any of a list of COVID-19-related signs and symptoms since February 2020. COVID-19-related illness was defined as having 1) loss of smell or taste; 2) at least two other COVID-related signs and symptoms from the following list: runny nose, couch, headache, sore throat, fever, arthralgia, myalgia, diarrhea, vomiting, fatigue, rash, lethargy, conjunctivitis, nasal congestion, itchy throat, poor appetite, chills with or without shaking, abdominal pain, fainting, difficulty sleeping, and lightheadedness or dizziness; 3) at least one of the following more severe signs or symptoms: breathing difficulty, accelerated breathing, shortness of breath, chest pain, a sensation of tightening in the chest; or 4) being admitted to a hospital with COVID-19-like signs and symptoms. COVID-19 cases were defined as participants 1) who experienced COVID-19-related illness and were SARS-CoV-2-positive by RT-PCR, 2) had SARS-CoV-2 infections confirmed by ELISA and who reported COVID-19-related illness during the peak of the transmission period in the study population (March-July 2020, as indicated by RT-PCR results), or 3) experienced COVID-19-related illness and had an epidemiological connection such that someone in the household tested positive for SARS-CoV-2 by RT-PCR.

For PDCS datasets, participants with laboratory-confirmed infections who did not seek medical care were categorized as having had subclinical infections. For HICS datasets, participants with laboratory-confirmed infections who did not report loss of smell or taste, did not report at least two COVID-related signs and symptoms, did not report at least one more severe COVID-19-consistent signs and symptoms, and were not admitted to the hospital with COVID-19-like signs and symptoms were categorized as having experienced a subclinical infection.

*Testing criteria for cases*

PDCS participants who were ill and reported to the HCSFV during the study period were tested for an acute, symptomatic CHIKV and ZIKV infection if they exhibited certain clinical profiles. During the two chikungunya epidemics, these clinical profiles were: 1) fever and at least two of the following: headache, retro-orbital pain, myalgia, arthralgia, rash, hemorrhagic manifestations, and leukopenia [the 1997 World Health Organization (WHO) dengue case definition[21]]; 2) fever and at least two of the following: nausea or vomiting, rash, aches and pains, positive tourniquet test, leukopenia, and any dengue warning sign [the 2009 WHO dengue case definition[22]]; and 3) undifferentiated fever without evident cause, with or without any other sign, symptoms, or complete blood count finding. The three clinical profiles that constituted the study's testing criteria reflected the PDCS's original goal of studying DENV infections; however, the clinical profiles we used were expansive enough to capture the known manifestations of chikungunya and the official WHO case definition for chikungunya, which is simply fever and arthralgia.[23]

The PDCS was expanded to include Zika in August 2015, which in turn led to the addition of one additional clinical profile that triggered arboviral testing: 4) afebrile rash, with or without any other sign, symptoms, or complete blood count finding. This change to the testing scheme was enacted because early reports[24] and preliminary guidance from the Pan American Health Organization (PAHO) stated that Zika could infrequently present without fever.

From February to June 2020, HICS participants reported to the HCSFV with 1) fever or feverishness with cough, sore throat, or runny nose or 2) a lower respiratory illness with or without fever were tested for an acute SARS-CoV-2 infection by RT-PCR. In June 2020, after approval was obtained from an institutional review board, the testing criteria was expanded to include loss of test or smell, rash or conjunctivitis, and fever without a defined focus for all HICS participants.

**Statistical and computing considerations**

*GPS coordinates*

Nicaragua does not use a house numbering system, whereby every home in a given city is assigned a unique number along a street to define the household address. Instead, addresses are recorded in reference to common landmarks (*e.g.,* the large tree in the Mantica neighborhood) and geographical aspects (*e.g.,* three blocks toward the lake and one block up [East]). The address of each PDCS and HICS household is recorded upon participant recruitment into the cohort studies and confirmed at subsequent annual samples. When a participant moves to a new house, the new address and the date of the move is logged into the cohort studies' computer systems. GPS points are taken upon site visits to the approximate 2,000 PDCS households and 430 HICS households for annual field visits, follow-up questionnaires, and medical checks at the household, as needed. The annual field visits serve to encourage continued participation in the cohort studies, receive feedback from participants, collect annual samples for participants who did not visit the study health center on time, and survey participants regarding their use of both study and non-study health centers for medical attention.[10] Household address and GPS information for all members of the PDCS and HICS for all years we considered were systematically examined, validated, and geolocated by a group of study authors who live in Managua and are extensively familiar with the study area.

Individuals who lived in the same household were initially assigned the same GPS points. However, some statistical methods we used are not designed to work with different individuals who have identical GPS points; doing so would cause some of the algorithms we used to return an error. We therefore slightly jittered the points of all study participants. The average displacement (distance from original GPS point to jittered GPS point) was 0.09 meters (3.5 inches), with the maximum displacement being 0.16 meters (6.5 inches). The jittering was performed using the jitter function in R, which added a uniform amount of noise to the original GPS points. Spatial analyses used the EPSG:4326 coordinate reference system.

*Household correlation*

During ChikE1, ChikE2, and ZikaE, approximately 35-40% of PDCS households had a single participant. The HICS study design is explicitly household-based. Correlation of outcomes at the household level, due to shared environments, genetics, and behaviors, was therefore possible. This could lead to a violation of the independence assumption embedded in standard regression approaches and some of the other methods we used. The possibility of household-based correlation necessitated the use of techniques that could account for or measure this across many of the analyses we performed. As detailed further below, this was accomplished through calculation of the intra-cluster correlation coefficient, mixed-effects geospatial models, generalized estimating equations, and generalized additive models with random effect spline terms. All analyses accounted for household-based correlation, unless otherwise noted.

*At-risk populations*

For each epidemic, the main analyses examined the risk (*i.e.,* the proportion of an eligible population that experienced an outcome of interest in a defined period of time) of infection among the at-risk population. Because CHIKV and ZIKV were introduced into the PDCS during ChikE1 and ZikaE, respectively, every participant enrolled in the PDCS at the time of ChikE1 and ZikaE was at risk of infection. Similarly, all participants in the HICS were at risk of infection from SARS-CoV-2 when the virus entered the study area. During ChikE2, those PDCS participants who had been CHIKV-infected during ChikE1 were no longer at risk of incident infection, so they were removed from consideration for all analyses of ChikE2. Main analyses also examined the risk of disease by measuring the proportion of the infected population to experience disease, as only infected persons are at risk of experiencing illness. In the arboviral field, this second risk is typically expressed as an odds, the symptomatic-to-inapparent (infection) ratio. However, the risk (as it is a probability) is a more mathematically tractable quantity with better statistical properties. We therefore did not present any analyses, including those of disease status, on the odds scale.

Typical spatial analyses are case-based as they rely on a sample of cases (often obtained from a Ministry of Health or a registry of confirmed or suspected cases) and population data (often obtained from a census). The typical metric of interest is the case-only incidence rate (also known as the attack rate or incidence proportion) in a given area, which is usually expressed as a scaled version (often by 10,000 or 100,000) of the proportion of cases among the overall population in the same area. Formally, the ratio of new cases to the total population is termed the incidence proportion in epidemiology; however, we use the term incidence rate here as that is the most commonly used term in spatial health research to refer to the same metric. To highlight the properties of its unscaled version, compare it with the risk of infection and the risk of disease, and showcase its relationship with the risk-based measures (Eq. 1), we use the incidence rate's unscaled version. However, by virtue of having many uninfected persons in the denominator, which by definition cannot experience the disease of interest, case-based spatial studies estimate neither the risk of infection nor the risk of disease. The proportion of disease among the overall population is often interpreted as conveying the risk of infection[25–27] and/or the risk of disease.[28,29] The main analyses were re-run to examine how different our conclusions would be if we had ignored infection status (as is typical) and treated the overall population (rather than only the infected population) as being at-risk for disease occurrence.

*Generalized linear models*

Generalized linear models (GLM) generalize linear regression (the general linear model) by allowing the linear predictor (systematic component) to be related to possibly non-continuous outcomes through a variety of link functions. Instead of directly modeling the outcome variable as in linear regression, a GLM models a function of the mean of the outcome variable. We used logistic regression (a GLM with a logistic link function and an assumed binomial distribution for the outcome variable [random component]) to estimate odds ratios (ORs) and corresponding 95% confidence intervals (CIs) for factors plausibly related to infection and disease status among their respective at-risk populations. This, as well we every other analysis except the cluster detection, was performed in R (version 3.6.2).[30]

The GLM estimation framework returns unbiased measures of association under a critical assumption of independence. In a spatial context, this assumption may be violated two different ways: by correlated data among persons living in the same household and/or by correlated data among people living close to each other in space. Both of these possible sources of bias necessitated the use of more complex model types to formally address.

The risk difference is the difference between two risks (proportions). For each epidemic we considered, we calculated the risk difference for the infection risk and the disease risk, with those risks being compared among males and females. To perform this analysis, we used the traditional approach, a binomial linear model (binomial distribution, identity link function) by way of the blm R package.[31]

*Generalized estimating equation models*

Models employing generalized estimating equations (GEE)[32] are extensions of GLMs[33] that allow for modeling of more complex data structures, including correlated data. Intercept-only logistic GEE models were used, for each epidemic, to estimate 1) the risk of an initial infection (infections / total population), 2) the risk of disease (cases / infected population), and 3) the case-based incidence rate (cases / total population). GEE models were run using the geepack R package (version 1.3-1)[34] with an exchangeable correlation structure, Huber-White sandwich standard error estimators,[35] and the scale parameter for a generalized binomial distribution fixed to 1.

Model results were backwards-transformed from the logit scale to the probability scale. In these models, household-based clustering was dealt with by setting the household ID variable as the clustering variable, such that infection and disease outcomes across households are assumed to be independent. Estimation of infection risk accounting for household-based clustering was performed with a GEE model instead of a mixed-effects model because estimates from logistic GEE models, unlike logistic mixed-effects models, are interpretable as population-level averages across the clustering variable.[36,37]

To account for household-based clustering in the estimation of the risk difference, we used modified Poisson regression within a GEE framework in addition to the aforementioned binomial GLM approach. The modified GEE approach involved using a Poisson distribution, the identity link function, the household as the clustering variable, and robust standard errors.[38,39]

*Generalized additive models*

Generalized additive models (GAMs)[40,41] are semi-parametric extensions of GLMs that fit smooth functions to the data to account for possible non-linear trends. GAMs are particularly well-suited to model continuous variables, such as exact age or GPS coordinates, because they can capture complex, non-linear relationships that would be missed by standard GLMs. All GAMs were estimated with the mgcv R package (version 1.8-31).[41] GAMs used restricted maximum likelihood (REML), as recommended,[42,43] to estimate the optimal smoothing parameters,[43,44] and used the outer, Newton numerical optimization method for smoothing parameter estimation. Thin plate splines were used.

Household-based clustering was dealt within the GAM estimation framework by including a random-effect penalized smoothing basis on the household ID variable. These smooth terms are parametric and ridge-penalized, such that the model coefficients are independent and identically distributed Gaussian random effects. Model predictions from the GAMs to the probability scale excluded the random effect terms, so that they were zeroed out in the prediction process.

Due to the connections between GAM estimation theory and Bayesian statistics,[41,45] uncertainty intervals produced by mgcv around a fitted penalized spline are Bayesian credible intervals. These intervals also have a surprising frequentist interpretation as across-the-function intervals,[45,46] such that 95% of these 95% confidence intervals will contain the true function of interest under repeated sampling of the same size from the same population of interest.

Logistic GAMs were used to assess the overall trend of exact age with the risk of infection and the risk of disease. To account for the higher percentage of adult females than adult males in the HICS, the logistic GAMs to assess the risk-based trends by age were weighted by the inverse probability of being female. First, a GAM was constructed that modeled the probability of being female across age for the HICS population. Second, the predicted probabilities for each HICS participant were obtained, which were used to generate the inverse probability weights for being female. Then the weights were normalized and applied to logistic GAMs to assess the trend of exact age with the risk of infection and the risk of disease. The weights were also applied to GEE models for the overall risk of infection and the risk of disease.

Spatial logistic GAMs (for both contour and perspective plots) used a bivariate spatial smooth (two-dimensional spline) that took the jittered longitude and latitude coordinates for a given individual as the input. The contour and perspective plots display the predicted probability of the outcome after holding the relevant covariates constant at their median values across all epidemics. The use of a single set of median values for contour and perspective plots allow for cross-epidemic and cross-plot comparisons for PDCS data. As the HICS study population has a much larger age range than the PDCS, median values for the HICS data were used to parameterize GAMs for HICS data. Contour and perspective spatial GAMs were plotted using a modified vis.gam() function from the mgcv R package to enhance the aesthetic properties of the output. For ease of comparisons, all perspective plots were visualized at an azimuthal angle of 47.5º and a colatitude angle of 30º.

Spatial logistic GAMs were used to estimate the risk of infection, the risk of disease, and the incidence rate for the spatial extent of the study area (Figure 3, rows 1-3). To calculate the spatial bias that would result from interpreting the case-only incidence rate as the risk of infection (Figure 3, row 4), the raster for the infection risk was subtracted from the raster for the incidence rate, as the former would represent the expected value of the latter under the stated interpretation. A similar approach was taken to estimate the bias that would result from interpreting the case-only incidence rate as the risk of disease (Figure 3, row 5).

*Intracluster correlation coefficient*
The intracluster correlation coefficient (ICC) measures the similarity of outcomes within clusters. The ICC is calculated as the ratio of the between-cluster variance to the sum of the within- and between-cluster variance. The value of the ICC ranges between 0 and 1; the value of the ICC indicates the magnitude of correlation among the outcomes within clusters, in the case, participants' households.

An ICC of 0 indicates that outcomes within the cluster are uncorrelated with each other, as the values in the cluster are not similar to each other (maximum within-cluster variation, complete lack of a cluster effect). An ICC of 1 indicates that outcomes within a cluster have the same outcomes as each other (minimum within-cluster variation, complete presence of a cluster effect). If households served as the site of viral transmission, we would expect the ICC to be closer to 1 (reflecting an abundance of either all-infected households or all-uninfected households) than 0. To not bias the ICC by the inclusion of households that only have 1 participant (and hence have a mathematically degenerate within-household variation), only households with at least 2 participants were used in the estimation of the ICC. However, when results were run with all households, they did not differ appreciably.

Three different ICC estimators (one based on an analysis of variance (ANOVA) methodology, one based on a GEE model, and one based on resampling) were used. There is a vast literature on different ICC estimators' properties,[47–49] and we aimed to demonstrate that our results were robust to the specification of the ICC estimator. First, we estimated the ICC using the traditional one-way ANOVA approach, which is based on a common correlation model. Searle's exact confidence limit equations[50] were used in the ICCest() function of the ICC R package (version 2.3.0)[51] to calculate the corresponding 95% CIs. Second, we used a multivariable logistic GEE model with an exchangeable correlation structure to measure the ICC. The variance of the correlation coefficients were extracted from geese() function output, specifically the valpha term. GEE models for CHIKV and ZIKV infection adjusted for exact age, sex, water availability, and distance to the cemetery. GEE models for SARS-CoV-2 infection adjusted for exact age and sex. GEE models for disease given CHIKV, ZIKV, and SARS-CoV-2 infection conditioned on exact age and sex for all comparisons. The GEE models for Zika occurrence given ZIKV infection further conditioned on prior DENV infection status for participants examined during ZikaE. Because the GEE approach used a parametric model to estimate the correlation coefficient, CIs from this approach tend to be the smallest of the three uncertainty intervals we assessed. Third, the ICC was estimated using the modern approach of Chakraborty and Sen,[52] which is based on resampling and U-statistics. ICC estimation with this approach was achieved using the iccbin() function of the ICCbin R package (version 1.1.1).[53] CIs were also estimated using the resampling-based approach, which tends to result in the largest CIs as a result of its estimation procedure. While new and not as commonly used as the ANOVA estimator, the resampling estimator has been shown to more accurately estimate the population-level ICC than the ANOVA estimator.[52]

*Household infection risk vs household size*

If households functioned as major sites of CHIKV and ZIKV transmission, then household-level infection risk (*i.e.*, number of infected participants in a household/number of participants in a household) could be larger for households of larger sizes, as larger households would have more opportunities for mosquito breeding grounds and a larger concentration of individuals that mosquitoes could feed on.[54,55] Similarly, if households functioned as major sites of SARS-CoV-2 transmission in our study area, it is plausible that household infection risk could be higher for larger households, as subclinical and pre-symptomatic household members could infect many persons in the households before precautionary steps to isolate a household member with COVID-19 are taken upon illness onset. To examine this association in our study setting, we plotted household size vs household infection risk, both with respect to cohort participants only, as they are the only members of the households for whom we have data. We plotted the population-averaged, intercept-only GEE estimate of infection risk, which averages the estimated infection risk across households of different sizes. We then overlaid the result of a LOESS (locally estimated scatterplot smoothing) regression[56,57] to show the underlying trend in the data. The span of the LOESS algorithm was set to the default value of 0.75.

*Kulldorff's spatial scan test*

Kuldorff's spatial scan test[58] was used to detect spatial clusters based on a significant excess or deficit of outcomes (whether infections or cases) within a moving window. This window visits all spatial locations and varies in size to detect small and large clusters. A purely spatial analysis was conducted with a Bernoulli probability model within SaTScan[59] (version 9.4.4). Clusters of high and low rates were scanned for with a circular spatial window. The maximum allowable size of a cluster was set at 50% of the population at risk (the default) for the outcome of interest, and 999 Monte Carlo simulations were run to generate p-values. By virtue of the estimation procedure, SaTScan examines millions of possible clusters, and inferential statistics are calculated for each of them. To correct for this, SaTScan adjusts p-values for the millions of multiple comparisons it conducts. By default, SaTScan maximizes the likelihood to identify hierarchical clusters, which are therefore the most statistically likely clusters. Hierarchical clusters are the most common type of clusters that are reported in spatial health research. They can be any size, but they tend to be large and may miss smaller, true clusters. As a result, we also used the SaTScan option to identify clusters by considering the Gini index.[60] These Gini clusters maximize outcome rates when comparing cluster and non-cluster areas. Due to the way Gini clusters are calculated, they tend to be smaller than hierarchical clusters, but they have higher rates of the outcome of interest. Statistically significant, non-geographically overlapping hierarchical clusters as well as significant Gini clusters were visualized, as is the norm in spatial health research. Numerical SaTScan output corresponding to the cluster analysis is presented. To identify clusters of high and low infection risk, we compared infected (cases) vs uninfected (controls) participants. (Here we use case-control nomenclature that is standard in the cluster detection literature. This nomenclature assigns a different meaning to *case* that does not imply a symptomatic infection, as we use *case* throughout most of the text. See Table S2 for further details.) To identify clusters of high and low disease risk, we compared symptomatic infections (cases) vs subclinical infections (controls). To identify clusters of high and low incidence, we compared symptomatic infections (cases) vs everyone else in the total population, which consisted of uninfected persons and those with subclinical infections (controls). This method does not account for household-based clustering.

*Moran's I test*
Global Moran's I[61] test was used to assess whether outcomes were spatially correlated (were autocorrelated). If autocorrelation of outcomes remains after conditioning on non-spatial variables, spatial correlation should be accounted for through spatial modeling. For Moran's I test, deviance residuals from non-spatial, logistic mixed-effects models were used as the numerical vector to assess whether outcomes remained autocorrelated after adjusting for covariates. The weights for Moran's I test corresponded to inverse distances between participants' locations. As Moran's I test is intended for use with continuous outcomes and the infection and disease outcome data was binary, we did not necessarily infer that spatial models were called for if Moran's I test indicated that outcomes were autocorrelated. We holistically considered the p-values, test statistics, other statistical output from Moran's I tests for all models, and the plausibility that the outcomes were autocorrelated to decide whether to use spatial regression models, ultimately concluding that they were needed. Because Moran's I test was used as an intermediary step to determine whether spatial regression models were needed, we do not present the results of Moran's I tests. To characterize whether the outcomes were autocorrelated, we conducted Moran's I tests using the Moran.I() function from the ape R package (version 5.3).[62]

*Statistical models for estimating measures of association*
A series of regression models were constructed to estimate measures of association for a distinct set of risk factors related to each outcome of interest, which were chosen *a priori* (see the "Covariate selection" section below). This was done to evaluate the impact on point and interval estimates after accounting for household-based clustering and spatial correlation. First, standard logistic GLMs were built, which account for neither clustering nor for autocorrelation. GLMs were estimated with the glm() function in base R.

Second, logistic generalized linear mixed models (GLMMs)[63,64] were built with the same covariate set (detailed below) as GLMs, incorporating household-based clustering by including the household ID as a random intercept. GLMMs are very flexible extensions of GLMs that can account for a variety of clustered and longitudinal data structures, similar to GEE models. However, GLMMs account for clustering through the use of random effects. GLMMs were estimated by way of the glmer() function of the lme4 R package (version 1.1-21).[65] GLMMs were fit by maximum likelihood approaches and used 25 Gauss-Hermite quadrature iterations for parameter estimation, as adaptive Gauss-Hermite quadrature approximation is appropriate for one random effect and is a more accurate approach than the default Laplace approximation. Unless otherwise noted, we used the default bobyqa optimizer for the first phase of optimization (random effects parameters only) and the Nelder-Mead optimizer for the second phase of optimization (random effects and fixed effects parameters). To avoid model convergence issues, we ran some models using the bobyqa optimizer for both optimization steps; when this was done, the maximum number of function evaluations to try was set to 100,000. GLMMs were used to estimate measures of association instead of GEE models because spatial versions of GLMMs exist. In addition, GLMs, non-spatial GLMMs, and spatial GLMMs share an overarching likelihood-based statistical framework that GEE models do not.

Third, spatial GLMMs were implemented with the fitme() function of the spaMM R package (version 3.1.2).[66] Spatial GLMMs are geostatistical extensions of GLMMs that accommodate spatial correlation by modeling autocorrelation directly. Logistic spatial GLMMs were constructed with a random intercept on the household ID variable. The spatial structure of our observations was modeled with a Matérn covariance function on the jittered longitude and latitude variables. Laplace maximum likelihood approximation was used to estimate the correlation parameters and $\lambda$, the variance term of the random effects. Fixed effects were estimated by $h$-likelihood approximation using the penalized quasilikelihood (PQL),[64] specifically the variant that does not use the leverage corrections of REML (PQL/L); by circumventing these corrections, PQL/L uses a marginal likelihood approximation for the estimation of the dispersion parameters. For fixed effect estimation, the PQL/L method was chosen over the default Laplace approximation due to concerns that the former (including its second-order variant[67]) could lead to separation, introducing bias into the estimates.[66] For nugget estimation, the initial value of the parameter was set to 0.5. To examine the impact of household-based correlation vs spatial correlation, a spatial GLMM without accounting for household clustering was constructed to be the spatial version of a GLM. Similarly, we used a spatial GLMM with household adjustment to be the spatial version of the non-spatial GLMM.

Using the MaternCorr() function of the spaMM R package and the estimated spatial correlation parameters $\nu$ and $\rho$, as well as the nugget parameter, from the spatial GLMMs, we visualized the estimated spatial correlation of two points across distance. Implementing spaMM functions with the size of our data was very computationally expensive on modern laptops, so all spaMM models were run for several days on the UC Berkeley Department of Biostatistics's computer cluster, which has with 288 processor cores, 768 gigabytes of memory, and over 10 terabytes of local storage.

*Covariate selection*

The covariate set of GLMs, non-spatial GLMMs, and spatial GLMMs for infection and disease were determined by our knowledge of the infection and disease processes for CHIKV, ZIKV, and SARS-CoV-2; prior literature regarding these outcomes; and our knowledge of the geographical conditions for the spatial extent of the study area. The covariate set for CHIKV and ZIKV infection was comprised of exact age, sex, estimated number of hours per day without tap water in the household, and distance (in 100m) from the slightly jittered GPS points to the closest boundary of the polygon representing a local cemetery. The covariate set for SARS-CoV-2 infection only included exact age and sex. The covariate set for disease included age and sex for chikungunya, Zika, and COVID-19 among infected individuals; the covariate set for Zika additionally included previous infection with DENV.

Exact (fractional) age was based on participants' birthdays and the date that participants last provided an annual sample within a given epidemic period. Exact age accounted for leap years. Age and sex were included in the covariate sex as they are standard demographic factors of interest with regard to infection and disease outcomes, and previous work has shown differences in outcomes, particularly for CHIKV and ZIKV infection in children[3,5] and SARS-CoV-2 infection across the life course.[68] Although it is known that the relationship between age and ZIKV infection in children is mostly reflective of mosquitoes' preference for larger targets as well as older children having a larger body surface area (BSA) and expelling more carbon dioxide (a mosquito chemoattractant),[3] we included age instead of BSA in the CHIKV and ZIKV infection models on the basis of interpretability. (It is difficult to interpret the additional risk of ZIKV infection for every 1 square meter of a child's BSA.)

Historical anecdotes from Managua-based study authors as well as many studies in the literature[69] suggested that the local cemetery abutting our study center may serve as a possible source of mosquitoes and hence CHIKV and ZIKV infection in the PDCS population. A site visit to the cemetery by the lead author in late August 2017 confirmed that many of the vases, gardens, water containers, artificial containers, crypts, and other aspects of the cemetery's built environment served as mosquito breeding sites. We included distance to the cemetery in 100-meter units in the infection models for CHIKV and ZIKV, as we hypothesized that mosquito-driven infection risk would be a function of Euclidean distance for the PDCS epidemics. As SARS-CoV-2 is not transmitted by mosquitoes, we did not include distance to the cemetery in the infection model corresponding to CovidE in the HICS population.

Repeated entomological inspections of the shoreline of Lago Xolotlán (Lake Managua) by members of the Department of Entomology of the Nicaraguan Ministry of Health have revealed breeding sites of *Anopheles* (*albimanus* and *pseudo punctipennis*) and *Culex quinquefasciatus* mosquitoes, but no *Aedes* breeding sites. *Anopheles* mosquitoes commonly found in North America are not known to transmit or be infected by ZIKV;[70] similarly, *Anopheles* mosquitoes are not known to be infected by CHIKV.[71] Despite early debate in the literature, at present, *Culex* mosquitoes are not believed to be competent vectors of ZIKV.[70,72] *Culex* mosquitoes are not thought to transmit CHIKV, and are very poor vectors of the closely related alphavirus, Mayaro virus.[73] As a result of this collective evidence, we did not include distance to the lake in the infection models for CHIKV and ZIKV. As we know of no relationship between SARS-CoV-2 transmission and large bodies of water, we also did not include distance to the lake for the SARS-CoV-2 infection model.

We hypothesized that water availability might be related to CHIKV and ZIKV infection risk because, in Managua, households with low water availability often store intermittent piped water as well as rainwater in barrels or other receptacles for indoor (*e.g.*, drinking, washing dishes) and outdoor (*e.g.*, gardening) activities. These water barrels can become ideal mosquito breeding sites if the water container walls are not scrubbed, if the water is not frequently replaced, or if larvicide is not used in them to control larvae growth.

Recent work has identified existing anti-DENV antibodies from a prior DENV infection as lowering the risk of a subsequent ZIKV infection becoming symptomatic.[20,74,75] While the exact causal mechanisms of this observation are unknown, this cross-protection is hypothesized to result from the similar immune responses induced to different flaviviruses, particularly cross-reactive antibody responses and/or cross-protective T-cell mediated responses.[20] Because there is no known cross-reactivity between CHIKV (an alphavirus), SARS-CoV-2 (a betacoronavirus), and DENV (a flavivirus), we only included prior DENV infection in the covariate set for the disease models of Zika.

To the best of our knowledge, there is no existing work showing that distance to cemeteries or other mosquito hotspots influences whether a CHIKV or ZIKV infection becomes symptomatic. However, we added distance to the cemetery in contour plots of the risk of chikungunya and Zika to examine the hypothesis that the spatial variation in disease risk is related to elevated viremia from repeated mosquito bites, which are likely to occur close to the cemetery. Direct adjustment for viremia was not possible because viremia levels during acute infections can only be known for the cases; persons with subclinical infections are not serologically assessed during their period of acute infection as they have no reason to report to the study's health center.

Previous work has shown that elevation is related to arboviral infection risk, as *Aedes* mosquitoes can vary substantially by elevation.[76] However, we did not believe that elevation would impact the distribution of the mosquito population, and hence infection risk, in our study area because the elevation gradient across the study area is too small (42-106m above sea level, per the R raster package[6]), to affect the spatial density of mosquitoes. Therefore, we did not include elevation in the models for CHIKV and ZIKV infection.

Historically, the household questionnaire that is administered during the annual sampling has been used to construct a validated, principal-components-based proxy variable for socioeconomic status (SES).[77] For the construction of the SES proxy, the following variables are used: whether the household floor is made of earth, whether the household walls are made of concrete, the number of refrigerators/freezers in the household, the number of fans in the household, the number of televisions in the household, and whether someone in the household owns a car or motorcycle. However, due to improving economic conditions over the years, household wealth as measured by this method has become relatively homogenous in recent years, explaining earlier null results for the association of ZIKV infection risk and SES.[3] For example, only 6 PDCS households are clearly separated from the others along the first principal component. All other households cluster close to each other on the first and second principal component, indicating that little variation exists among households with respect to the household SES proxy variable. Inclusion of a variable with such little variable could result in statistical separation, leading to bias. Consequently, we did not include a proxy variable for SES in models for either infection or disease.

*Spatiotemporal GAMs*

Temporal data were only available for the subset of participants in each outbreak who were cases, as they reported to health center when they were sick with chikungunya, Zika, or COVID-19, and provided an estimated date of illness onset in a subset of the HICS population. The infection date of subclinical infections is unknown. Case data were treated like a Poisson point process to investigate the overall spatiotemporal dynamics of cases and infections among the overall population, with only the latter representing a measure of risk.[78] In particular, cases were treated as a realization from an intensity surface, and the overall study population was used as the offset since its size remained constant across the various months of the epidemic periods.

Cases' reported date of illness onset was used as the temporal variable. For each epidemic, a data frame of case data (*i.e.*, date of illness onset, jittered longitude, jittered latitude, and coordinate reference system) was turned into an sf object using the sf R package (version 1.4-1),[79] which was then used as the points argument in the space_time_ppmify() function of the disarmr R package (version 0.0.3).[80] The spatial distribution of the overall study population was aggregated on a 33x33 raster, and this raster was used as the exposure argument in the space_time_ppmify() function. The periods argument, defining the time slices of the analyses, corresponded to a vector of the first date of each month for the duration of a given epidemic period. The approximate number of integration points was set to 10,000 and the output returned a rasterStack for prediction purposes. Poisson regression was applied to the resulting data frame, using the log of the exposure as the offset and the regression weight supplied by space_time_ppmify(). Poisson regression was performed using the bam() function of the mgcv R package,[41] with a tensor product on the bivariate (two-dimensional) spatial smooth given by participants' longitude and latitude and a univariate (one-dimensional) smooth on time period as measured by the month of participants' illness initiation. Thin plate and cubic regression splines were used, with 50 and 5 respective basis functions, for these two- and one-dimensional smooths, respectively.

Predictions for case counts across the raster for the overall population were made for each month and visualized as level plots by way of the levelplot() function from the rasterVis R package (version 0.47).[81] The predicted number of cases were visualized per 1,000 persons in the overall study population.

To back-calculate the spatiotemporal dynamics of infection risk, we relied on our unique data structure: a cross-sectional understanding of infection risk and the temporal case data for each epidemic. We also exploited the algebraic relationship between the three quantities we repeatedly estimate across this analysis: the product of the risk of infection (infected / total) and the risk of disease (cases / infected) equals the incidence rate (cases / total), which is Equation 1 in the main text. The spatiotemporal analysis previously outlined gave a spatiotemporal estimate of the incidence rate. The use of spatial GAMs had given an estimate of the risk of disease across the study area for each epidemic (Figure 3, row 2). We assumed this risk, while it varied in space, was constant over time. As far as we know, there is no evidence in the literature that this measure varies over the timescale of a single, short epidemic caused by CHIKV, ZIKV, or SARS-CoV-2. Further, we could identify no host or environmental factors that plausibly and substantially changed the value of the risk of disease over the few months of the three epidemics we assessed. While our assumption was untestable with the available data, we judged it probable that factors influencing disease occurrence were temporally stable over each of the short epidemic periods that peaked during a 3-4-month timeframe.

To estimate the spatiotemporal dynamics of infection risk, we used the space_time_ppmify() function to make a purely spatial Poisson model of the risk of disease. This analysis thus used the infected population instead of the overall study population in space_time_ppmify() to replicate the results of the binomial GAM model (Figure 3, row 2) but in raster form. The resulting raster was stacked to create a RasterStack object of the same number of rasters as the months of spatiotemporal predictions for the corresponding epidemic. The spatiotemporal predictions of case occurrence among the overall study population, also a RasterStack object, was then divided by the RasterStack object with multiple rasters of the risk of disease. As before, the data was visualized as level plots. The predicted number of infections were visualized per 1,000 persons in the overall study population. Arranging of the spatiotemporal levelplots, created using rasterVis,[81] into a single figure was facilitated by the gridExra R package.[82]

For all spatiotemporal maps, neighborhood boundaries and contour lines were superimposed on model predictions for ease of interpretation. Based on information from PDCS families regarding their use of medical services from the HCSFV,[10] as well as the results of the clustering analyses for disease outcomes, we did not believe that treatment-seeking behavior spatially varied among PDCS participants over the course of the three PDCS epidemics we assessed. As many HICS households are also PDCS households, the same treatment-seeking behavior in PDCS household likely applies to HICS households. Therefore, we did not seek to account for this factor in the spatiotemporal models.

The spatiotemporal dynamics were only estimated for the infection risk and the incidence rate, which both have the total population in their denominator. The spatiotemporal dynamics of the disease risk was not estimated as its denominator, the infected population, rapidly changes over the course of an epidemic, unlike the total population. Consequently, accurately estimating the spatiotemporal dynamics of the disease risk would be statistically intractable without further assumptions and data.

*Comparison of active versus passive data*

Zika is known to exhibit a broad clinical spectrum, including a very mild clinical profile.[1] In addition, the viremic period of ZIKV is shorter and peak ZIKV viremia is lower than other arboviruses. Together, the clinical and serological features of an acute Zika case complicate the detection of ZIKV. As a result, we instituted additional measures during ZikaE in our testing criteria for suspected cases and in our laboratory testing methods to capture all Zika cases in the PDCS. As previously detailed,[1,20] we expanded our testing criteria, beyond those cases meeting the 1997 and 2009 WHO case definitions for dengue[82] and those presenting with undifferentiated fever, such that all participants presenting to the study health center with a clinical profile of afebrile rash were tested for the presence of ZIKV. In addition to detection by rRT-PCR, we used a serological algorithm to identify acute Zika cases. As many PDCS participants have been previously infected with DENV, anti-DENV and anti-ZIKV antibodies cross-react, and there was DENV transmission during ZikaE, we could not rely on standard serological assays to complement rRT-PCR and help us identify additional Zika case. Thus, we developed and validated a serological algorithm to accomplish this goal. Due to our enhanced methods, we captured more Zika cases than would be possible with standard approaches, which has been noted previously.[1]

To compare the results of our active surveillance to a more typical approach, we considered the design of a standard a passive surveillance system that would capture cases during ZikaE. Standard studies would likely have tested suspected cases meeting either the WHO[83] or PAHO[84] case definitions for Zika, as those case definitions were then, and are still today, the official guidance by global health bodies on the clinical presentation of Zika. In addition, standard studies would likely have relied on rRT-PCR for confirmation, as IgM assays were unreliable given the epidemiological conditions of ZikaE at the time. The use of more specific plaque reduction neutralization tests (PRNT), while helpful, would have been extremely time-consuming because of the large number of cases who met the testing criteria during ZikaE (N=1,110).[1] Moreover, evidence suggests that PRNTs cannot unambiguously distinguish between ZIKV and DENV infections in populations, such as the PDCS, with a high proportion of prior DENV infections.[85]

A standard passive surveillance system in place during ZikaE would thus capture the 133 rRT-PCR-positive Zika cases who met the Zika case definitions established by either the WHO or PAHO. The total number of Zika cases captured by our active surveillance system, representing the complete case count, is 494, as per Table 2. A clinical comparison of Zika cases meeting the WHO and PAHO case definitions and those captured by our full active surveillance procedures has been published.[1]

We calculated the infection risk, disease risk, and incidence rate using the ZikaE serosurvey data and the Zika case count from active surveillance. These three metrics were then compared to those estimated from the ZikaE serosurvey data and the Zika case counts from a standard passive surveillance system. Importantly, this comparison also reveals what would have occurred if a serosurvey had not been conducted, as the incidence rate is the only metric estimable in the absence of infection data. As before, the bias arising from treating the incidence rate as a risk was calculated as the difference between the incidence rate and each of the infection and disease risks. This was done with the data attainable under both study designs (*i.e.,* serosurvey paired with active case surveillance and the serosurvey paired with passive case surveillance). Similarly, the bias arising from incomplete case ascertainment data was calculated as the difference between each of the three metrics under the study design with passive case surveillance and their true value under the study design with active case surveillance. The compounded bias of treating the incidence rate obtained under passive surveillance as the true infection risk was estimable directly as the difference between those two metrics. It was also indirectly estimable by summing the bias of the infection risk under passive surveillance and the bias arising from treating the incidence rate (under passive surveillance) as the infection risk (also under passive surveillance). A similar calculation was used to estimate the compounded bias for the disease risk. Maps of the infection risk, disease risk, and incidence rate (under both study designs); the inferential bias; the case ascertainment bias; and the compounded bias were created using generalized additive models, as previously described.

*Data visualization*
When possible, data were visualized with the ggplot2 R package (version 3.3.0)[82,83] with uncertainty intervals (pointwise 95% CIs or confidence bands, as appropriate). The arranging of different ggplot2 plots into panels was facilitated by the patchwork R package (version 1.1.0).[84] When possible, data corresponding to the four epidemic periods are graphed on the same plot with the same aesthetics settings to enable cross-epidemic comparisons. However, for some plots, we separately graphed data from the PDCS and HICS cohorts to convey important differences between the populations. Smoothed density plots of exact (fractional) age were constructed with Gaussian kernels, and smoothing bandwidths were set equal to the standard deviations of the kernels. For various figures, outcomes automatically output in decimal degrees were transformed to meters for ease of interpretation. As Nicaragua is close to the equator, 1 decimal degree was taken to be equal to 111.32 kilometers for conversion purposes.

Predicted outcomes on the absolute scale were visualized with the plasma, inferno, magma, and viridis color palettes of the viridis R package (version 0.5.1)[85] and the turbo color palette in the viridisLite R package (version 0.4.0).[86] These color palettes were specifically chosen as they provide perceptually uniform, sequential, grayscale-friendly, and color-blind friendly options for the visualization of spatial data. All analysis operations were performed on the continuous output of spatial models. However, mapped continuous data were discretized during the data visualization step at increments of five percentage points to better interpret color-coded outcomes at specific locations. Contour lines were overlaid on maps.

Visualizations of the same phenomenon across the four epidemics were shown using the same color range to facilitate cross-epidemic comparisons. All scales bars in the main figures were dynamic such that they only contained the colors of the corresponding panel, even if the range of the scale bars were larger. Whenever a figure contained panels with different sets of data (*e.g.*, the panels with the infection risk, disease risk, and incidence rate data in addition to the bias panels in Figure 3), a different color palette and scale bar range were used to differentiate visually between the different types of panels. To further aid the reader in making visual contrasts, the background color of panels was also changed from white to another color in these circumstances. Adding a background color to some panels was necessary to aid the visual differentiation because the limited number of perceptually uniform color palettes results in some of those color palettes have somewhat overlapping color ranges. The addition of a background color to the panels aids in distinguishing color palettes that may have overlapping colors. Thus, panels of similar data are visually united by three separate elements: the color palette, the range of the scale bar, and the background color.

When using the vis.gam() function of the mgcv R package, it was not possible to clip the resulting contour plot to the polygon of the study area. Thus, analyses that relied on this function for output retained the default continuous color output and overlaid contour lines to better demarcate intervals on the prediction surface.

*Data management*
Data management was performed in the RStudio integrated development environment.[87] R packages not previously mentioned that contributed significantly to the data wrangling and cleaning include readxl (version 1.3.1),[88] WriteXLS (version 5.0.0),[89] dplyr (version 1.0.3),[90] Hmisc (version 4.3-1),[91] varhandle (version 2.0.5),[92] and magrittr (version 1.5).[93]

### Supplementary text: Limitations of the case-only incidence rate

The purpose of this text is to illustrate the limitations of the common, case-only incidence rate (cases / total population) in the context of pathogens that cause subclinical (clinically inapparent) infections. In particular, we focus on interpretability issues that arise when conducting risk factor analyses of the incidence rate in this context.

We did not conduct multivariable analyses of disease status among the entire population since it was unclear how to biologically interpret a resulting odds ratio (OR). As an extended example, imagine restricting to the CHIKV-infected population and conducting a logistic regression analysis of the association of red hair (yes / no) with disease status (yes / no) during ChikE1. If the OR associated with having red hair had been 2, its interpretation would be: CHIKV-infected redheads have twice the odds as CHIKV-infected non-redheads of experiencing chikungunya. Assuming the regression result was not otherwise biased, this OR would biologically suggest that something about having red hair (or related to being a redhead) was associated with an increased risk of CHIKV-infected persons experiencing chikungunya. Thus, the risk-based OR has a clear interpretation, both epidemiologically and biologically.

However, with an analysis that considers the disease status among the entire population, the interpretation of a similar OR would be: redheads in the overall population (including those who are CHIKV-naïve) have twice the odds as non-redheads in the overall population (including those who are CHIKV-naïve) of experiencing chikungunya. First, this is an unusual epidemiological interpretation because persons who are CHIKV-naïve are not at risk for experiencing chikungunya. That is, mathematically (measure theoretically), the probability of CHIKV-naïve persons experiencing chikungunya is 0 because the outcome (chikungunya occurrence) is impossible for CHIKV-naïve persons; their potential outcomes do not include chikungunya. The disease is only a potential outcome, in the measure theoretic sense, among those that are CHIKV-infected. It is therefore not clear what it means to have twice the odds of chikungunya occurrence when the majority of both exposure groups (redheads and non-redheads) were impervious to exhibiting chikungunya signs and symptoms because they were never CHIKV-infected in the first place.

Second, there are two kinds of non-cases: uninfected persons and infected persons who nevertheless did not exhibit a disease phenotype. The factors the gave rise to the first set of non-cases may be different than those relevant to the second set of non-cases. Moreover, there is no guarantee that factors shared by both sets of non-cases would have similar strengths of association, let alone the same significance or directionality of association. More broadly, this implies that the same factors can have the different strengths of association, statistical significance, and directionality of association across the infection risk, disease risk, and incidence rate. Consequently, risk factors identified for the incidence rate – even if such factors have unclear interpretations – may be associated differently with the infection risk and disease risk. An explicit example of this has been published (see Table 3 of the cited reference).[20]

Third, estimates calculated from statistical models, such as the OR of 2, typically correspond to averages. But that OR does not necessarily translate to an increased risk of chikungunya occurrence among the average participant, who would be uninfected (as only 6% of the ChikE1 population was CHIKV-infected).

Finally, it is also not the case that the OR of 2 would hypothetically and necessarily apply, in the future, if the uninfected persons were infected. There may be systematic differences between uninfected and infected persons that impact the risk of disease. Thus, there is no guarantee that if all persons in the overall population would be infected, then the OR of 2 among the infected population would also be the average OR across the overall, now-infected population. Moreover, the risk of disease cannot be calculated among a sample of participants whose risk of the event is always 0. The risk of chikungunya occurrence for CHIKV-naïve persons remains 0 until a CHIKV infection occurs, as it is the qualifying event to be at risk for the outcome of interest. Just as a two-year-old who is assigned female at birth is not at risk for experiencing pregnancy until pubertal changes (the qualifying event), a person is not at risk for chikungunya occurrence until the qualifying event, infection from CHIKV, has occurred.

In sum, whether a factor is related to the infection process or the disease process is not knowable from an analysis of the incidence rate. Such analyses are unable to distinguish insights related to the separate processes of infection and disease. As a result, it is difficult to draw biological inferences from incidence rate in the context of a pathogen capable of causing clinically subclinical infections.

### Supplementary Tables

**Table S1.** Distribution of age and sex among the PDCS and HICS populations, by epidemic.

| | ChikE1 | ChikE2 | ZikaE | CovidE |
|---|---|---|---|---|
| **Female (%)** | 1,571 (50.3%) | 1,441 (50.3%) | 1,516 (50.2%) | 1,109 (61.9%) |
| **0-1 years old (%)** | 0 (0.0%) | 0 (0.0%) | 0 (0.0%) | 31 (1.7%) |
| **2-5 years old (%)** | 886 (28.4%) | 714 (24.9%) | 703 (23.3%) | 174 (9.7%) |
| **6-9 years old (%)** | 1,038 (33.2%) | 1,057 (36.9%) | 1,092 (36.2%) | 275 (15.3%) |
| **10-14 years old (%)** | 1,200 (38.4%) | 1,093 (38.2%) | 1,222 (40.5%) | 308 (17.2%) |
| **15-17 years old (%)** | 0 (0.0%) | 0 (0.0%) | 0 (0.0%) | 138 (7.7%) |
| **18-29 years old (%)** | 0 (0.0%) | 0 (0.0%) | 0 (0.0%) | 258 (14.4%) |
| **30-59 years old (%)** | 0 (0.0%) | 0 (0.0%) | 0 (0.0%) | 509 (28.4%) |
| **60-87 years old (%)** | 0 (0.0%) | 0 (0.0%) | 0 (0.0%) | 100 (5.6%) |

Abbreviations: ChikE1, first chikungunya epidemic; ChikE2, second chikungunya epidemics; CovidE, COVID-19 epidemic; HICS, Household Influenza Cohort Study; PDCS, Pediatric Dengue Cohort Study; ZikaE, Zika epidemic

**Table S2.** SaTScan output for the purely spatial cluster analyses.[1]

| Epidemic | Cluster type, case-control comparison[2] | Cluster number[3] | Radius (km) | Cluster type | N cases[2] in cluster / overall N in cluster | SMR[4] | Relative risk[5] | p-value |
|---|---|---|---|---|---|---|---|---|
| ChikE1 | Infection risk, Infected participants (cases) vs uninfected participants (controls) | 1 | 0.003 (a single house) | Gini | 5 / 5 | 15.78 | 16.16 | 0.003 |
| | | 2 | 0.78 | Hierarchical | 63 / 1517 | 0.66 | 0.49 | 0.014 |
| | | 3 | 0.35 | Gini | 3 / 296 | 0.16 | 0.15 | 0.016 |
| ChikE1[6] | Disease risk, Symptomatic infections (cases) vs subclinical infections (controls) | | | | | | | |
| ChikE1[6] | Incidence, Symptomatic infections (cases) vs uninfected persons & subclinical infections (controls) | | | | | | | |
| ChikE2 | Infection risk, Infected participants (cases) vs uninfected participants (controls) | 1 | 0.60 | Hierarchical | 62 / 477 | 0.52 | 0.48 | < 0.001 |
| | | 2 | 0.82 | Hierarchical | 430 / 1415 | 1.23 | 1.57 | < 0.001 |
| | | 3 | 0.29 | Gini | 70 / 155 | 1.82 | 1.91 | < 0.001 |
| | | 4 | 0.25 | Gini | 13 / 142 | 0.37 | 0.36 | 0.023 |
| ChikE2[6] | Disease risk, Symptomatic infections (cases) vs subclinical infections (controls) | | | | | | | |
| ChikE2 | Incidence, Symptomatic infections (cases) vs uninfected persons & subclinical infections (controls) | 1 | 0.81 | Hierarchical | 40 / 576 | 0.48 | 0.42 | < 0.001 |
| | | 2 | 0.28 | Gini | 47 / 141 | 2.29 | 2.46 | < 0.001 |
| | | 3 | 0.28 | Gini | 49 / 174 | 1.94 | 2.06 | 0.034 |
| | | 4 | 0.55 | Gini | 17 / 284 | 0.41 | 0.39 | 0.039 |
| ZikaE | Infection risk, Infected participants (cases) vs uninfected participants (controls) | 1 | 1.07 | Hierarchical | 456 / 1169 | 0.83 | 0.75 | < 0.001 |
| | | 2 | 0.94 | Hierarchical | 629 / 1149 | 1.17 | 1.30 | < 0.001 |
| | | 3 | 0.21 | Gini | 98 / 143 | 1.46 | 1.49 | 0.003 |
| | | 4 | 0.06 | Gini | 30 / 34 | 1.88 | 1.90 | 0.009 |
| | | 5 | 0.27 | Gini | 41 / 194 | 0.59 | 0.57 | 0.016 |

| | | | | | | | | |
|---|---|---|---|---|---|---|---|---|
| ZikaE | Disease risk, Symptomatic infections (cases) vs subclinical infections (controls) | 1 | 0.26 | Gini | 27 / 37 | 2.09 | 2.15 | 0.037 |
| ZikaE | Incidence, Symptomatic infections (cases) vs uninfected persons & subclinical infections (controls) | 1 | 0.88 | Gini | 197 / 860 | 1.40 | 1.66 | < 0.001 |
| | | 2 | 1.07 | Hierarchical | 129 / 1113 | 0.71 | 0.60 | 0.002 |
| | | 3 | 0.67 | Gini | 103 / 892 | 0.71 | 0.63 | 0.042 |
| CovidE | Infection risk, Infected participants (cases) vs uninfected participants (controls) | 1 | 0.15 | Gini | 87 / 101 | 1.49 | 1.53 | < 0.001 |
| | | 2 | 0.27 | Gini | 3 / 26 | 0.20 | 0.20 | 0.006 |
| | | 3 | 0.24 | Gini | 47 / 125 | 0.65 | 0.63 | 0.020 |
| | | 4 | 0.12 | Gini | 19 / 19 | 1.73 | 1.74 | 0.039 |
| | | 5 | 0.15 | Gini | 11 / 44 | 0.43 | 0.43 | 0.045 |
| CovidE | Disease risk, Symptomatic infections (cases) vs subclinical infections (controls) | 1 | 1.03 | Hierarchical | 115 / 516 | 0.76 | 0.61 | 0.003 |
| | | 2 | 0.79 | Gini | 69 / 343 | 0.68 | 0.59 | 0.012 |
| CovidE | Incidence, Symptomatic infections (cases) vs uninfected persons & subclinical infections (controls) | 1 | 0.00002 (a single house) | Gini | 9 / 11 | 4.79 | 4.91 | 0.016 |
| | | 2 | 1.28 | Hierarchical | 113 / 873 | 0.76 | 0.62 | 0.043 |
| | | 3 | 0.79 | Gini | 69 / 598 | 0.68 | 0.58 | 0.044 |

[1]Only data for statistically significant clusters is presented in this table, as is the norm in spatial epidemiology.

[2]Cluster detection analyses use case-control terminology differently than this paper. Whereas we use *case* to denote a symptomatic infection, *case* means the group of interest and *control* means the control group in cluster detection nomenclature. Thus, when assessing the risk of infection, for example, infected participants constitute the case group and uninfected participants constitute the control. We apply the case-control nomenclature of cluster detection analyses to this table as that is the standard in the field of spatial health research.

[3]Cluster number is with respect to Figure 4.

[4]The SRM is a measure of association that compares the observed number of outcomes in the cluster to the expected number of outcomes in the cluster if (under the null) it had exhibited the same proportion of outcomes as the entire study area.

[5]The relative risk (risk ratio) is a measure of association that compares the proportion of outcomes in the cluster to the proportion of outcomes outside the cluster.

[6]No significant clusters for this outcome were identified.

Abbreviations: ChikE1, first chikungunya epidemic; ChikE2, second chikungunya epidemic; CovidE, COVID-19 epidemic; SMR, standardized mortality ratio; ZikaE, Zika epidemic

**Table S3.** Odds ratios and 95% CIs for infection risk, estimated from increasingly complex logistic models.[1,2]

| Epidemic | Variable | Non-spatial GLM | Non-spatial GLMM[3] | Spatial GLMM, without household adjustment[4] | Spatial GLMM, with household adjustment[5] |
|---|---|---|---|---|---|
| ChikE1 | Distance to cemetery (per 100m) | 0.98 (95% CI: 0.96, 1.01) | 0.98 (95% CI: 0.94, 1.02) | 0.99 (95% CI: 0.93, 1.05) | 0.99 (95% CI: 0.92, 1.05) |
| | Age (per 1 year) | **1.09 (95% CI: 1.05, 1.13)** | **1.13 (95% CI: 1.07, 1.20)** | **1.10 (95% CI: 1.05, 1.15)** | **1.10 (95% CI: 1.05, 1.15)** |
| | Female sex | 0.86 (95% CI: 0.64, 1.14) | 0.79 (95% CI: 0.54, 1.15) | 0.82 (95% CI: 0.61, 1.12) | 0.83 (95% CI: 0.61, 1.13) |
| | Hours w/o water (per 1 hour) | 1.01 (95% CI: 0.98, 1.04) | 1.03 (95% CI: 0.98, 1.08) | 1.01 (95% CI: 0.97, 1.05) | 1.01 (95% CI: 0.97, 1.05) |
| ChikE2[6] | Distance to cemetery (per 100m) | **0.96 (95% CI: 0.94, 0.97)** | **0.94 (95% CI: 0.92, 0.97)** | **0.96 (95% CI: 0.93, 0.99)** | **0.96 (95% CI: 0.93, 0.99)** |
| | Age (per 1 year) | **1.13 (95% CI: 1.10, 1.16)** | **1.17 (95% CI: 1.13, 1.21)** | **1.13 (95% CI: 1.10, 1.17)** | **1.13 (95% CI: 1.10, 1.17)** |
| | Female sex | 1.03 (95% CI: 0.86, 1.22) | 1.00 (95% CI: 0.81, 1.25) | 1.02 (95% CI: 0.84, 1.22) | 1.00 (95% CI: 0.83, 1.21) |
| | Hours w/o water (per 1 hour) | 0.99 (95% CI: 0.96, 1.01) | 0.99 (95% CI: 0.96, 1.02) | 0.99 (95% CI: 0.96, 1.02) | 0.99 (95% CI: 0.96, 1.02) |
| ZikaE | Distance to cemetery (per 100m)[7] | **0.95 (95% CI: 0.94, 0.97)** | **0.95 (95% CI: 0.93, 0.97)** | **0.96 (95% CI: 0.94, 0.97)** | **0.96 (95% CI: 0.94, 0.97)** |
| | Age (per 1 year) | **1.13 (95% CI: 1.11, 1.16)** | **1.18 (95% CI: 1.14, 1.21)** | **1.14 (95% CI: 1.11, 1.17)** | **1.14 (95% CI: 1.11, 1.17)** |
| | Female sex | **1.28 (95% CI: 1.11, 1.49)** | **1.35 (95% CI: 1.12, 1.62)** | **1.28 (95% CI: 1.10, 1.50)** | **1.29 (95% CI: 1.10, 1.51)** |
| | Hours w/o water (per 1 hour) | 1.01 (95% CI: 0.99, 1.03) | 1.02 (95% CI: 0.99, 1.05) | 1.01 (95% CI: 0.98, 1.03) | 1.01 (95% CI: 0.98, 1.03) |
| CovidE | Age (per 1 year) | **1.01 (95% CI: 1.00, 1.01)** | **1.01 (95% CI: 1.00, 1.02)** | **1.01 (95% CI: 1.00, 1.02)** | **1.01 (95% CI: 1.00, 1.02)** |
| | Female sex | 1.21 (95% CI: 0.99, 1.47) | 1.21 (95% CI: 0.94, 1.55) | 1.17 (95% CI: 0.93, 1.48) | 1.17 (95% CI: 0.93, 1.48) |

[1]Statistically significant covariates (at the α-level of 0.05) are bolded. Some of the models do not return p-values, but whether or not covariates are statistically significant at the specified α-level can be inferred from whether the CI includes the null value of 1.

[2]In general, results from generalized linear, mixed, geostatistical, and mixed geostatistical multivariable models were similar because infection and disease outcomes were not clustered within households (Table S5) and both outcomes were spatially correlated over small distances (Figure S26). Accounting for moderate-to-substantial levels of clustering would have resulted in enlarged CIs from the mixed models' coefficients relative to the CIs from the GLM.

[3]This model was the one evaluated using Moran's I test to assess whether autocorrelation (spatial correlation) for infection remained after accounting for the covariates and household-based clustering. As Moran's I test indicated the need to account autocorrelation, we proceeded to build spatial models.

[4]The spatial GLMM that does not account for household clustering is the geostatistical analogue of the non-spatial GLM.

[5]The spatial GLMM that does account for household clustering is the geostatistical analogue of the non-spatial GLMM.

[6]The non-spatial GLMM was run with the bobyqa optimizer for both optimization steps to avoid model convergence issues.

[7]Infection models for CHIKV and ZIKV were parameterized with distance to the cemetery on a 100m basis as the study area is ~3km as its widest. The seemingly small reduction in odds of infections per 100m from the cemetery compounds multiplicatively such that, during ZikaE, the conditional odds of infection among participants living 1 km from the cemetery were 0.63 (95% CI: 0.55, 0.73), as reported in the main text.

Abbreviations: ChikE1, first chikungunya epidemic; ChikE2, second chikungunya epidemic; CovidE, COVID-19 epidemic; CI, confidence interval; GLM, generalized linear model; GLMM, generalized linear mixed model

**Table S4.** Odds ratios and 95% CIs for disease risk, estimated from increasingly complex logistic models.[1,2]

| Epidemic | Variable | Non-spatial GLM | Non-spatial GLMM[3] | Spatial GLMM, without household adjustment[4] | Spatial GLMM, with household adjustment[5] |
|---|---|---|---|---|---|
| ChikE1 | Age (per 1 year) | **1.11 (95% CI: 1.03, 1.20)** | **1.12 (95% CI: 1.03, 1.27)** | **1.11 (95% CI: 1.02, 1.20)** | **1.11 (95% CI: 1.02, 1.20)** |
| | Female sex | 0.77 (95% CI: 0.43, 1.36) | 0.72 (95% CI: 0.28, 1.36) | 0.77 (95% CI: 0.43, 1.36) | 0.76 (95% CI: 0.43, 1.36) |
| ChikE2 | Age (per 1 year) | 1.03 (95% CI: 0.99, 1.08) | 1.05 (95% CI: 0.99, 1.12) | 1.04 (95% CI: 0.99, 1.09) | 1.04 (95% CI: 0.99, 1.08) |
| | Female sex | 0.90 (95% CI: 0.67, 1.22) | 0.86 (95% CI: 0.57, 1.28) | 0.90 (95% CI: 0.66, 1.24) | 0.91 (95% CI: 0.66, 1.24) |
| ZikaE | Age (per 1 year) | 1.02 (95% CI: 0.98, 1.06) | 1.03 (95% CI: 0.98, 1.08) | 1.02 (95% CI: 0.98, 1.06) | 1.02 (95% CI: 0.98, 1.06) |
| | Female sex | 1.14 (95% CI: 0.92, 1.43) | 1.20 (95% CI: 0.89, 1.62) | 1.15 (95% CI: 0.91, 1.45) | 1.15 (95% CI: 0.90, 1.45) |
| | Prior DENV infection | **0.63 (95% CI: 0.49, 0.81)** | **0.54 (95% CI: 0.37, 0.77)** | **0.64 (95% CI: 0.48, 0.84)** | **0.64 (95% CI: 0.48, 0.84)** |
| CovidE | Age (per 1 year) | **1.03 (95% CI: 1.02, 1.04)** | **1.04 (95% CI: 1.03, 1.05)** | **1.03 (95% CI: 1.03, 1.04)** | **1.04 (95% CI: 1.03, 1.04)** |
| | Female sex | 1.10 (95% CI: 0.82, 1.48) | 1.17 (95% CI: 0.81, 1.70) | 1.14 (95% CI: 0.82, 1.60) | 1.16 (95% CI: 0.82, 1.62) |

[1]Statistically significant covariates (at the α-level of 0.05) are bolded. Some of the models do not return p-values, but whether or not covariates are statistically significant at the specified α-level can be inferred from whether the CI includes the null value of 1.

[2]In general, results from generalized linear, mixed, geostatistical, and mixed geostatistical multivariable models were similar because infection and disease outcomes were not clustered within households (Table S5) and both outcomes were spatially correlated over small distances (Figure S26). Accounting for moderate-to-substantial levels of clustering would have resulted in enlarged CIs from the mixed models' coefficients relative to the CIs from the GLM.

[3]This model was the one evaluated using Moran's I test to assess whether autocorrelation (spatial correlation) for disease remained after accounting for the covariates and household-based clustering. As Moran's I test indicated the need to account autocorrelation, we proceeded to build spatial models.

[4]The spatial GLMM that does not account for household clustering is the geostatistical analogue of the non-spatial GLM.

[5]The spatial GLMM that does account for household clustering is the geostatistical analogue of the non-spatial GLMM.

Abbreviations: ChikE1, first chikungunya epidemic; ChikE2, second chikungunya epidemic; CovidE, COVID-19 epidemic; CI, confidence interval; DENV, dengue virus; GLM, generalized linear model; GLMM, generalized linear mixed model

**Table S5.** Intracluster correlation coefficient[1] and 95% CIs for infection outcomes and disease outcomes given infection across the four epidemics.

| Outcome of interest | Method | ChikE1 | ChikE2 | ZikaE | CovidE |
|---|---|---|---|---|---|
| Infection | ANOVA[2] | 0.22<br>(95% CI: 0.17, 0.28) | 0.21<br>(95% CI: 0.16, 0.27) | 0.22<br>(95% CI: 0.17, 0.27) | 0.30<br>(95% CI: 0.25, 0.35) |
| | GEE[3] | 0.24<br>(95% CI: 0.12, 0.35) | 0.20<br>(95% CI: 0.13, 0.26) | 0.18<br>(95% CI: 0.13, 0.23) | 0.27<br>(95% CI: 0.22, 0.32) |
| | Resampling[4] | 0.19<br>(95% CI: 0.00, 0.41) | 0.23<br>(95% CI: 0.14, 0.31) | 0.20<br>(95% CI: 0.13, 0.26) | 0.28<br>(95% CI: 0.21, 0.35) |
| Disease given infection[5] | ANOVA[2] | 0.14<br>(95% CI: 0.00, 0.51) | 0.26<br>(95% CI: 0.09, 0.42) | 0.28<br>(95% CI: 0.18, 0.37) | 0.21<br>(95% CI: 0.15, 0.28) |
| | GEE[3] | 0.00<br>(95% CI: 0.00, 0.28) | 0.25<br>(95% CI: 0.07, 0.42) | 0.25<br>(95% CI: 0.15, 0.36) | 0.25<br>(95% CI: 0.14, 0.36) |
| | Resampling[4] | 0.13<br>(95% CI: 0.00, 0.60) | 0.25<br>(95% CI: 0.05, 0.45) | 0.25<br>(95% CI: 0.13, 0.38) | 0.19<br>(95% CI: 0.08, 0.30) |

[1]The ICC ranges from 0-1. A value of 0 indicates outcomes within homes are not correlated. A value of 1 indicates outcomes within homes are fully correlated.

[2]Point estimates derived from the traditional one-way ANOVA-based approach and interval estimates derived from Searle's exact confidence limit equations.[50]

[3]Point and interval estimates derived from a logistic GEE model using an exchangeable correlation structure.[32] GEE models for CHIKV and ZIKV infection were adjusted for age, sex, water availability, and distance to the cemetery. GEE models for SARS-CoV-2 infection were adjusted for age and sex. GEE models for chikungunya given CHIKV infection were adjusted for age and sex. GEE models for Zika given ZIKV infection were adjusted for age, sex, and prior DENV infection. GEE models for COVID-19 given SARS-CoV-2 infection were adjusted for age and sex.

[4]Point and interval estimates derived from the modern resampling-based approach of Chakraborty and Sen.[52]

[5]The correlation of infection outcomes within households was the primary analysis; we have no reason to believe, based on the existing literature, that disease outcomes given infection clusters within households. However, we present estimates of the ICC for this outcome to compare with the ICC values of infection outcome.

Abbreviations: ANOVA, analysis of variance; ChikE1, first chikungunya epidemic; ChikE2, second chikungunya epidemic; CHIKV, chikungunya virus; CI, confidence interval; CovidE, COVID-19 epidemic; GEE, generalized estimating equations; COVID-19, coronavirus disease 2019; ICC, intracluster correlation coefficient; SARS-CoV-2, severe acute respiratory syndrome coronavirus 2; ZIKV, Zika virus; ZikaE, Zika epidemic

### Supplemental Figures

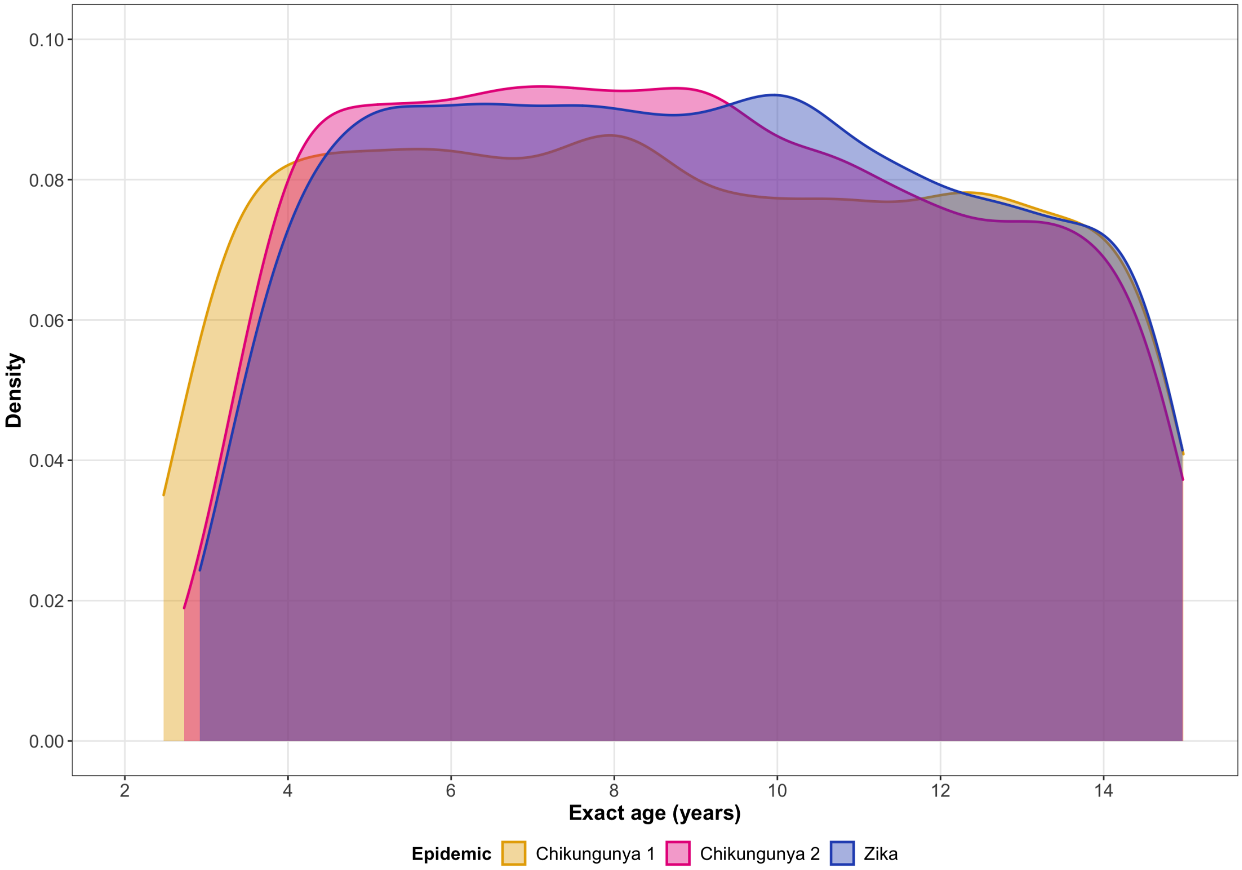


**Figure S1.** Distribution of exact age among PDCS participants by epidemic period.

Abbreviations: PDCS, Pediatric Dengue Cohort Study

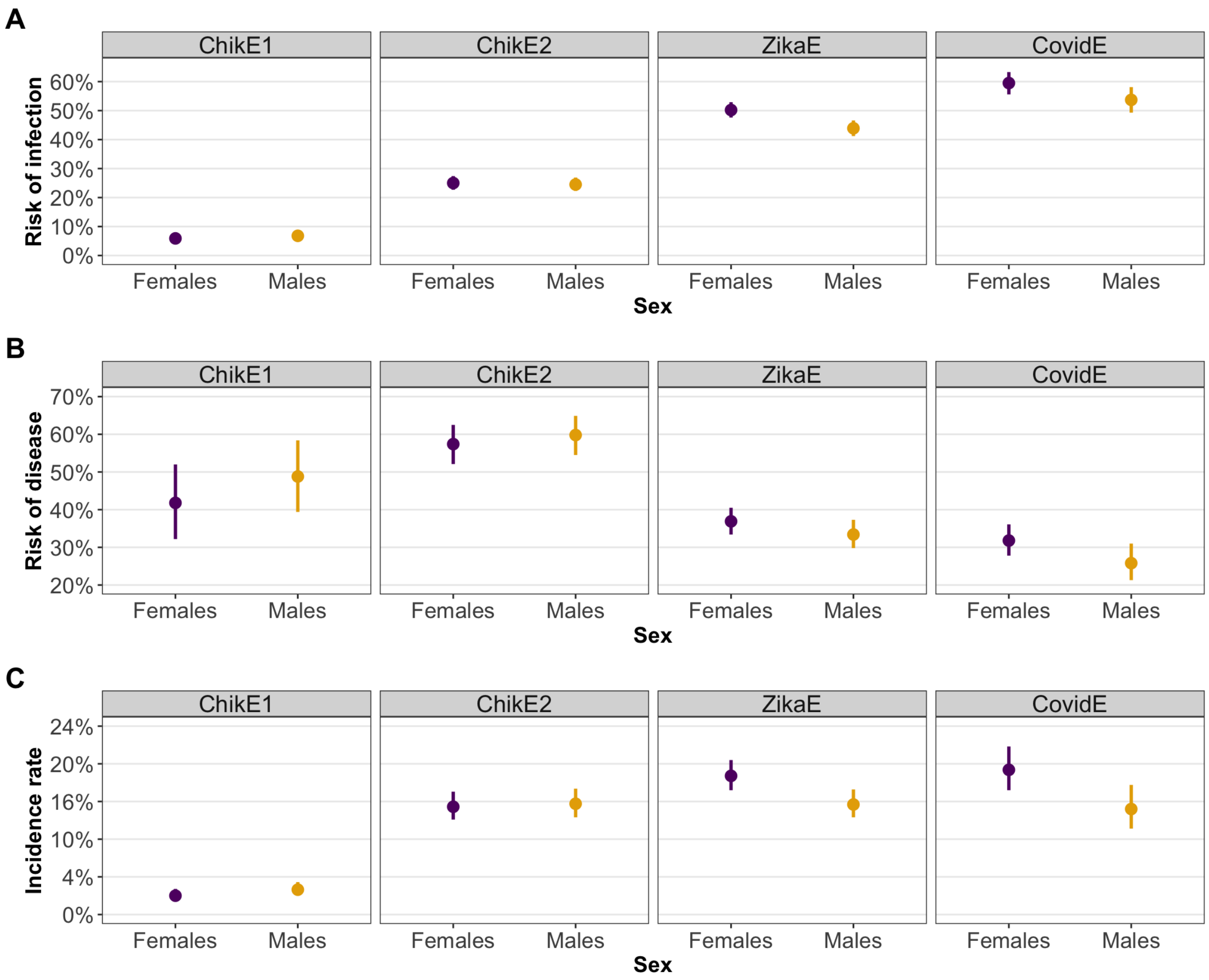


**Figure S2**. Point and interval (95% confidence interval) estimates for the risk of infection **(A)** risk of disease **(B)** and the incidence rate (**C**) by epidemic and sex. The was a significantly different risk difference (difference in proportions) for infection risk during ZikaE; female children had a higher risk of ZIKV infection (risk difference = 6.1%, 95% CI: 2.6, 9.5, p-value < 0.001) than male children by modified Poisson regression. The was a significantly different risk difference for infection risk during CovidE; females had a higher risk of SARS-CoV-2 infection (risk difference = 4.7%, 95% CI: 0.5, 9.0, p-value = 0.029) than males by modified Poisson regression. The was a significantly different risk difference for disease risk during CovidE; SARS-CoV-2-infected females had a higher risk of COVID-19 (risk difference = 6.3%, 95% CI: 0.8, 11.8, p-value = 0.025) than males by modified Poisson regression. The was a significantly different risk difference for the incidence rate during ZikaE; females had a higher Zika incidence rate (risk difference = 3.7%, 95% CI: 1.1, 6.2, p-value = 0.004) than males by modified Poisson regression. The was a significantly different risk difference for the incidence rate during CovidE; females had a higher COVID-19 incidence rate (risk difference = 4.9%, 95% CI: 1.6, 8.2, p-value = 0.003) than males by modified Poisson regression. All other comparisons resulted in non-statistically significant risk differences.

Abbreviations: ChikE1, first chikungunya epidemic; ChikE2, second chikungunya epidemic; CI, confidence interval; CovidE, COVID-19 epidemic; COVID-19, coronavirus disease 2019; SARS-CoV-2, severe acute respiratory syndrome coronavirus 2; ZikaE, Zika epidemic; ZIKV, Zika virus

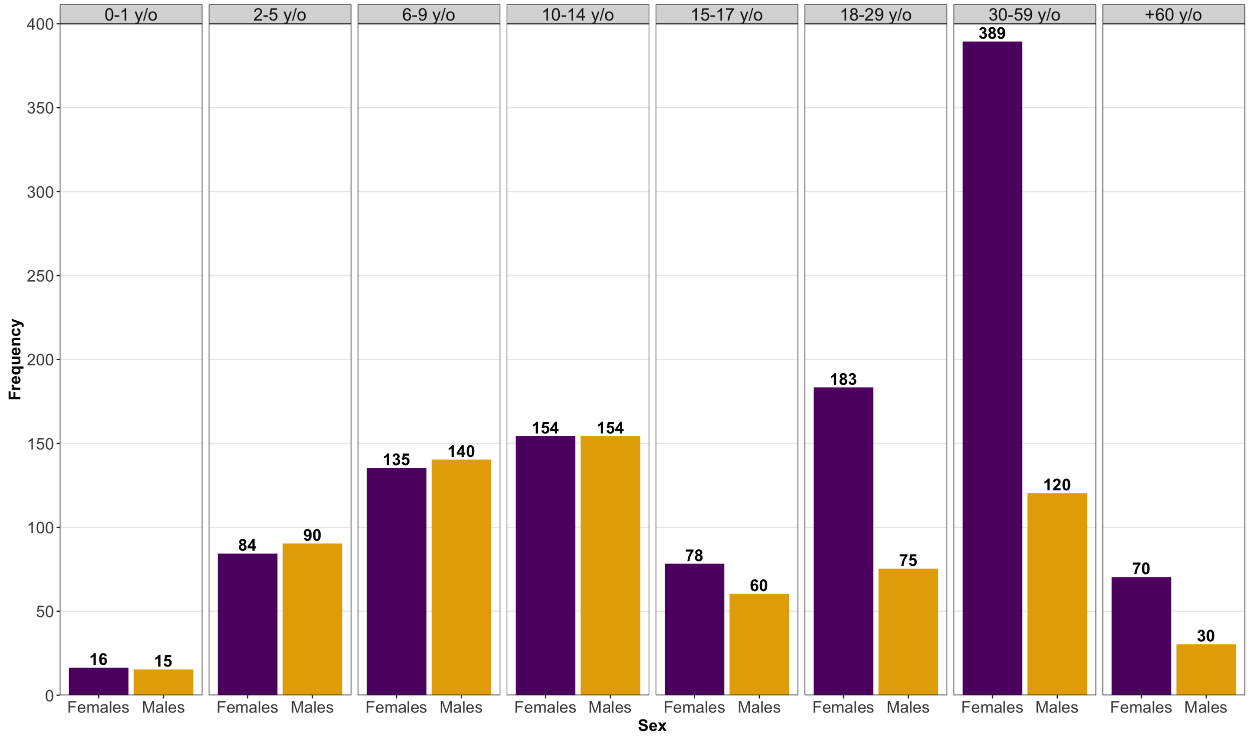


**Figure S3.** Sex distribution of the HICS population by age group.

Abbreviations: HICS, Household Influenza Cohort Study; y/o, year olds

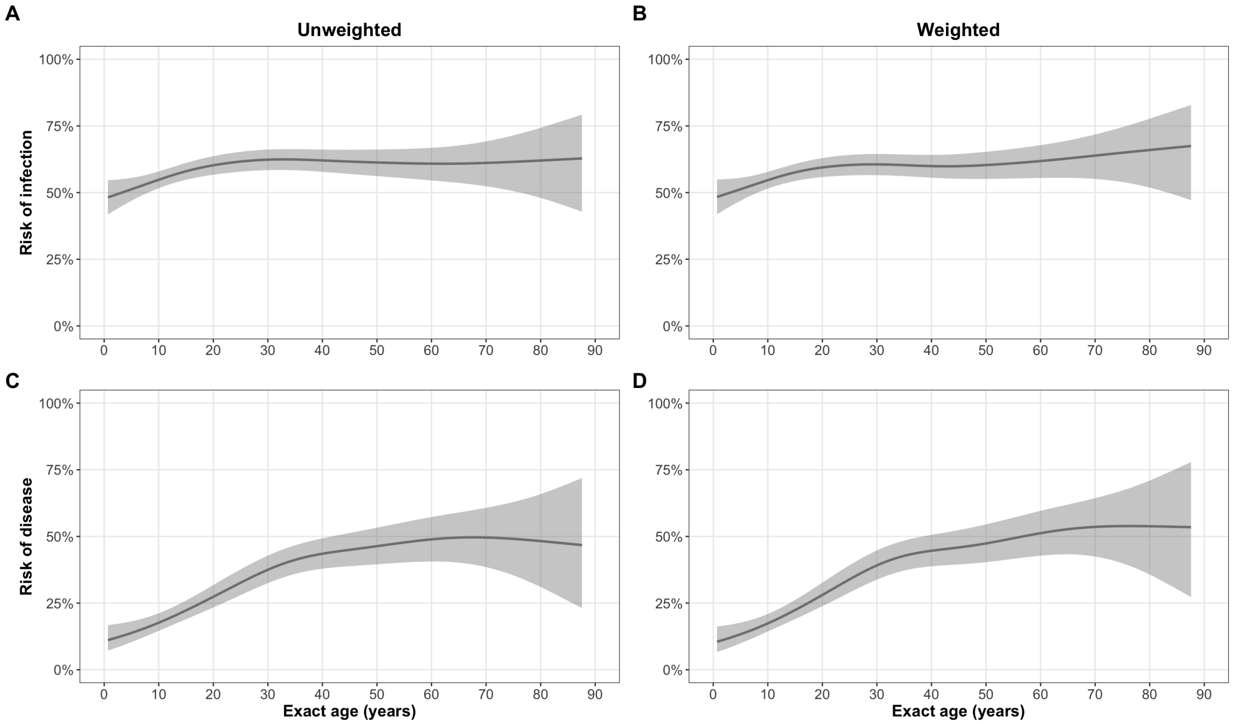


**Figure S4.** Age trends for the risk of infection (**A-B**) and the risk of disease (**C-D**) unweighted (column 1) and weighted (column 2) to account for the higher proportion of adult females in the HICS population. The overall, unweighted infection risk was 57.5% (95% CI: 54.1, 60.9) and the weighted infection risk was 56.6% (95% CI: 52.9, 60.3). The overall unweighted disease risk was 28.9% (95% CI: 25.5, 32.5) and the weighted disease risk was 29.8% (95% CI: 26.0, 33.9). Due to the very similar results, we did not employ the weighting scheme in other analyses.

Abbreviations: CI, confidence interval; HICS, Household Influenza Cohort Study

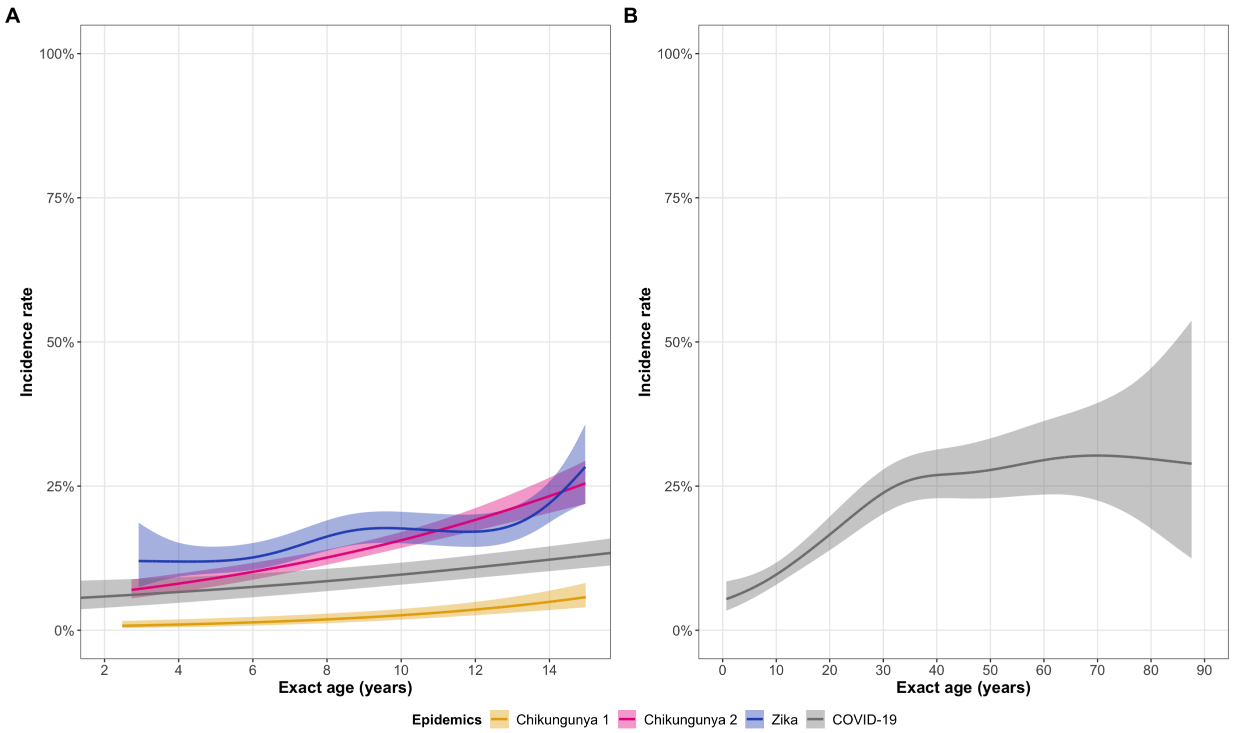


**Figure S5.** Age trends for the incidence rate, along with pointwise 95% confidence bands, for **(A)** the first chikungunya epidemic, the second chikungunya epidemic, the Zika epidemic, and the COVID-19 epidemic for the age range of the PDCS (2-14 years of age) as well as **(B)** the COVID-19 epidemic across the full age range in the HICS (0-87 years of age).

Abbreviations: COVID-19, coronavirus disease 2019; HICS, Household Influenza Cohort Study; PDCS, Pediatric Dengue Cohort Study

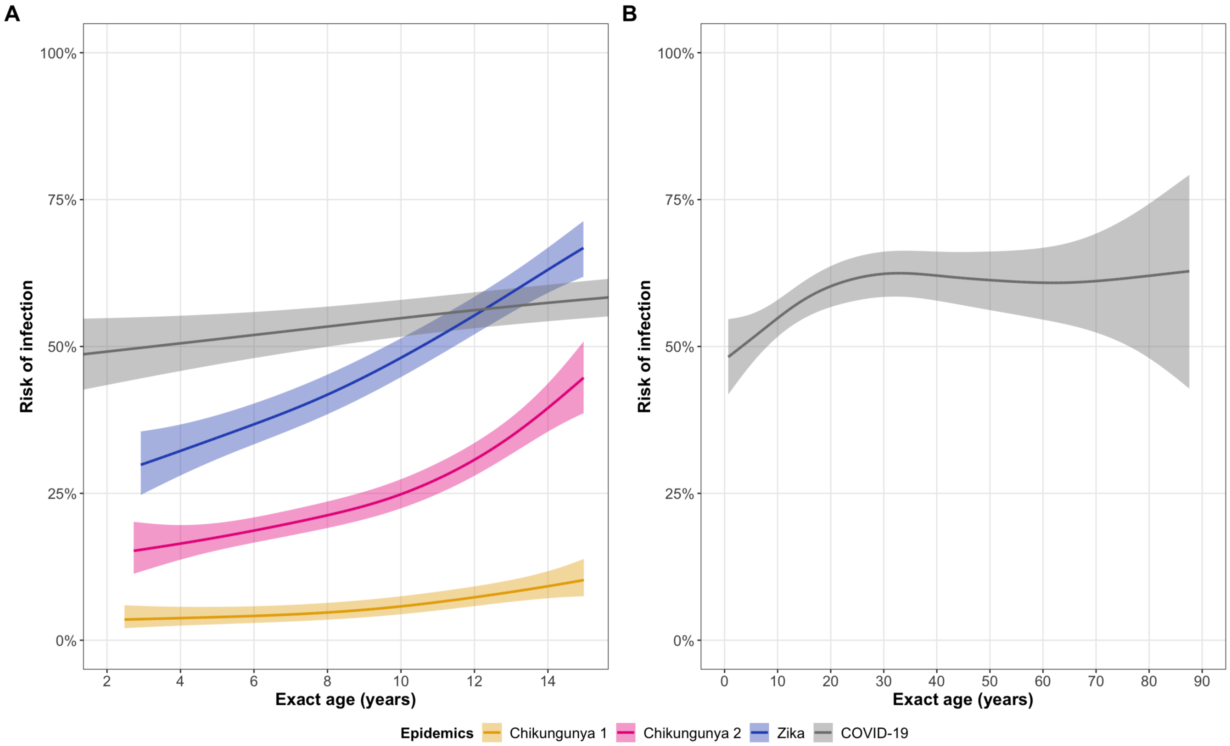


**Figure S6.** Age trends for the risk of infection, along with pointwise 95% confidence bands, for **(A)** the first chikungunya epidemic, the second chikungunya epidemic, the Zika epidemic, and the COVID-19 epidemic for the age range of the PDCS (2-14 years of age) as well as **(B)** the COVID-19 epidemic across the full age range in the HICS (0-87 years of age).

Abbreviations: COVID-19, coronavirus disease 2019; HICS, Household Influenza Cohort Study; PDCS, Pediatric Dengue Cohort Study

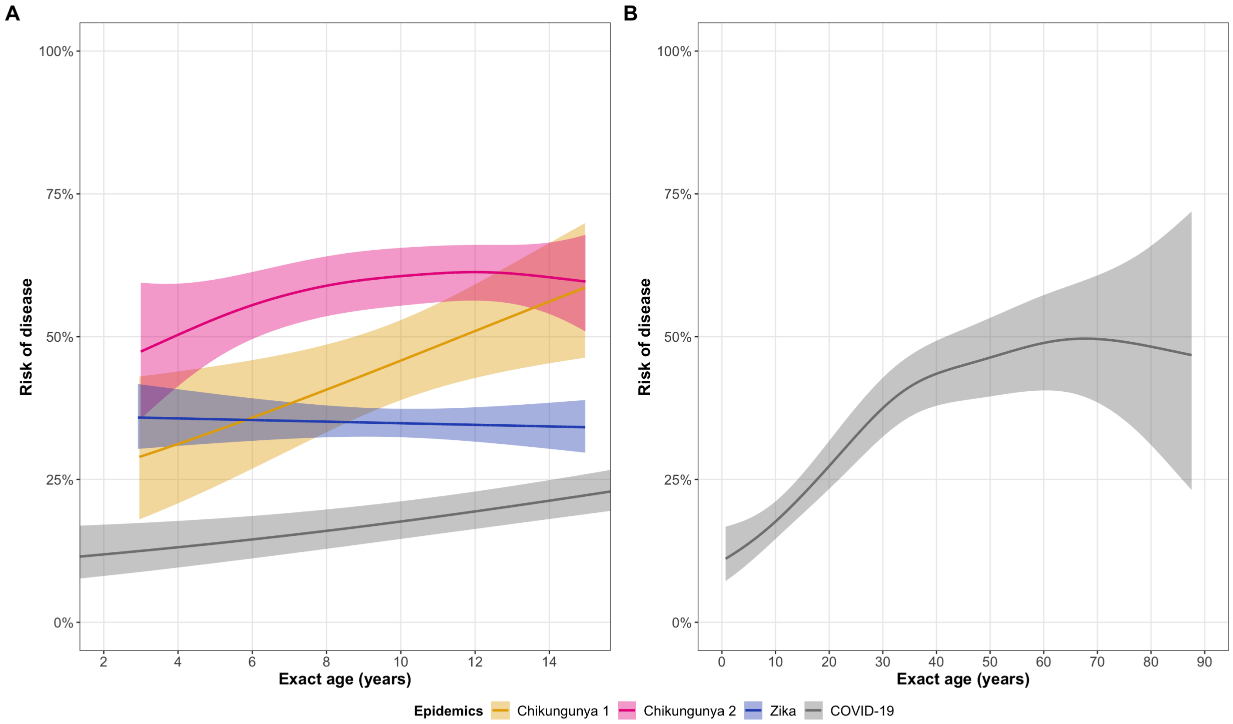


**Figure S7.** Age trends for the risk of disease, along with pointwise 95% confidence bands, for **(A)** the first chikungunya epidemic, the second chikungunya epidemic, the Zika epidemic, and the COVID-19 epidemic for the age range of the PDCS (2-14 years of age) as well as **(B)** the COVID-19 epidemic across the full age range in the HICS (0-87 years of age).

Abbreviations: COVID-19, coronavirus disease 2019; HICS, Household Influenza Cohort Study; PDCS, Pediatric Dengue Cohort Study

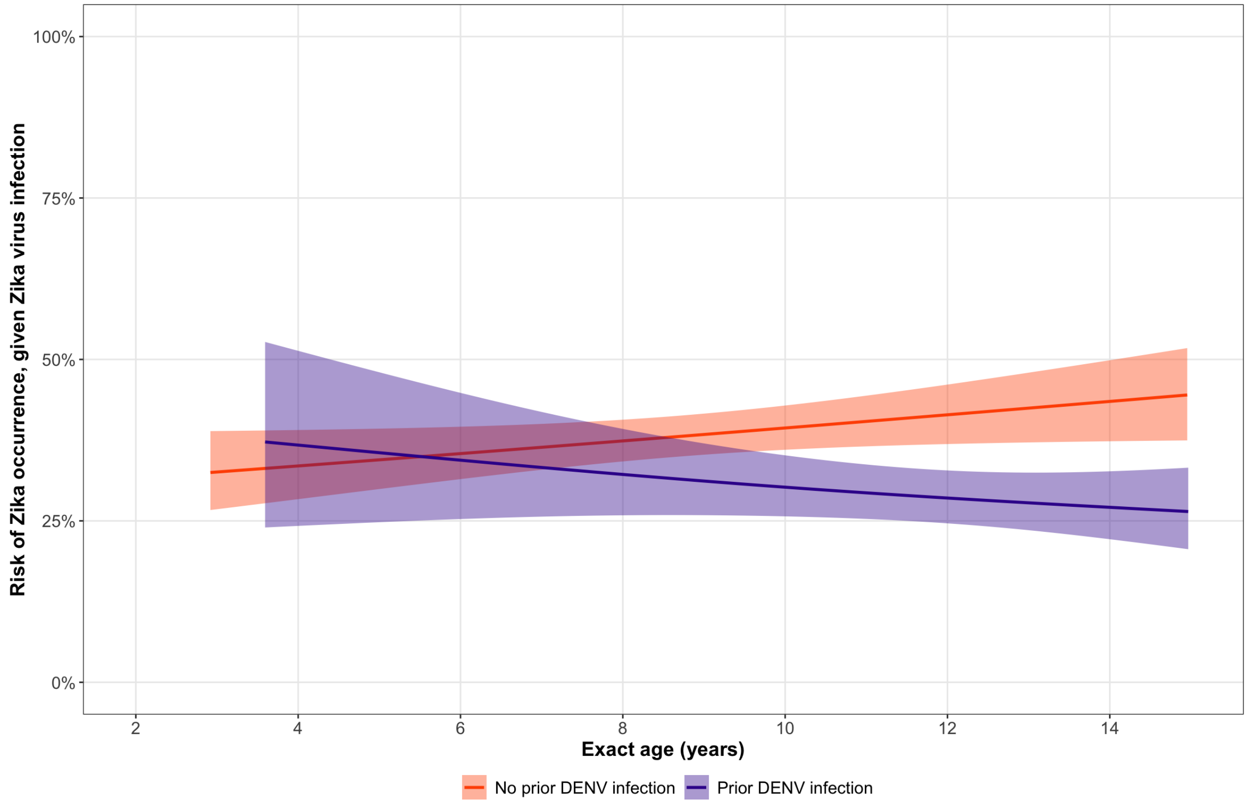


**Figure S8.** Age trends in the risk of Zika occurrence given ZIKV infection, by prior DENV infection status. 95% confidence bands are shown. Disease risk increased with age during all epidemics except ZikaE (as seen in Fig. S7). The overall negative age-disease trend during ZikaE reflects the established cross-protective effect of anti-DENV antibodies on Zika occurrence.[20] Among those without a prior DENV infection, the age-disease trend is positive.

Abbreviations: DENV, dengue virus; ZikaE, Zika epidemic; ZIKV, Zika virus

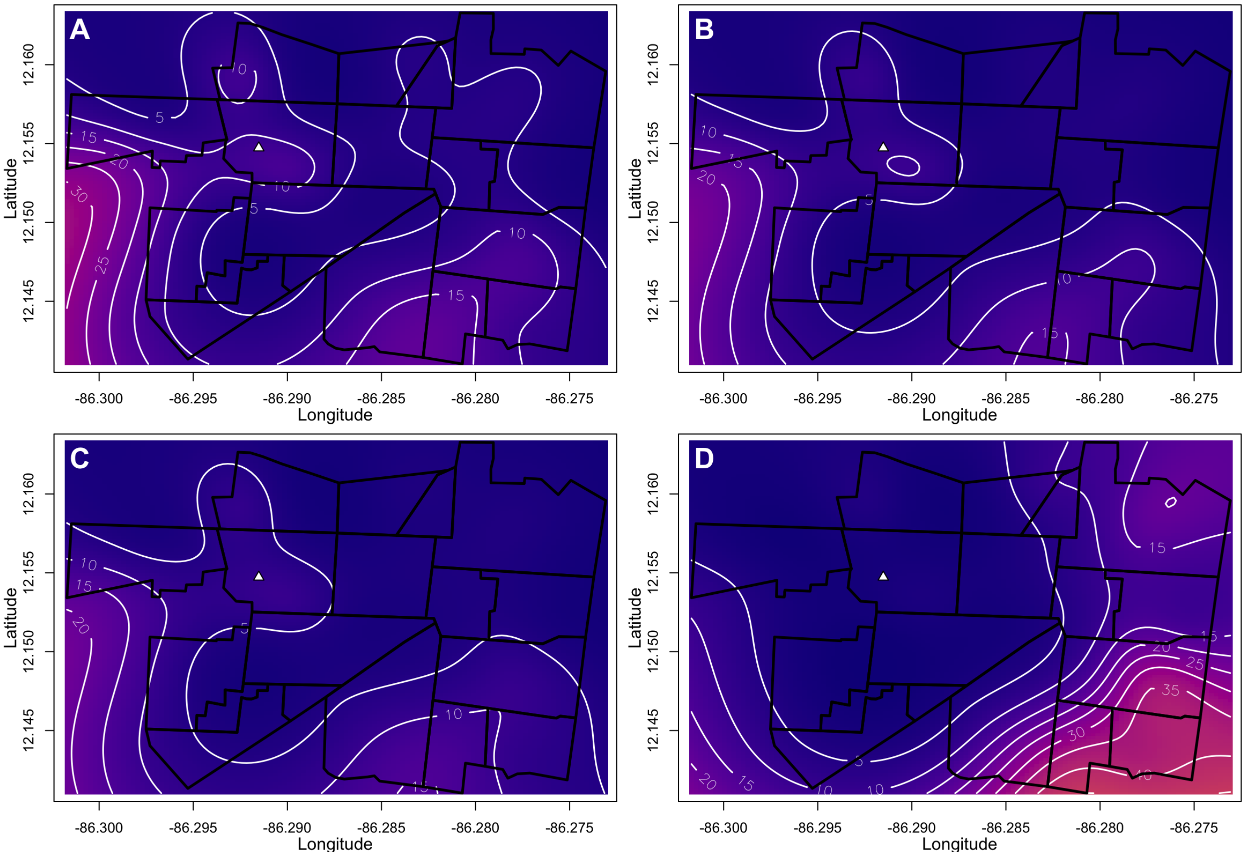


**Figure S9.** CHIKV infection risk estimated from spatial logistic GAMs during the first chikungunya epidemic. Panels show **(A)** unadjusted infection risk; **(B)** infection risk adjusted for age and sex; **(C)** infection risk adjusted for age, sex, and water availability; and **(D)** infection risk adjusted for age, sex, water availability, and distance to the cemetery. The variables in models for Panels B-D have been set to those of the median participant across the three PDCS epidemics to facilitate cross-epidemic comparisons. The median PDCS participant is a female of age 8.71 years living in a household that has 24-hour indoor access to tap water and is located 841.50 meters from the boundary of the local cemetery. Figures S9-11 and S17-19 use the same color scale to facilitate cross-epidemic comparisons in the PDCS population and to ensure visual comparability between maps that share the same median values.

Abbreviations: CHIKV, chikungunya virus; GAM, generalized additive model; PDCS, Pediatric Dengue Cohort Study

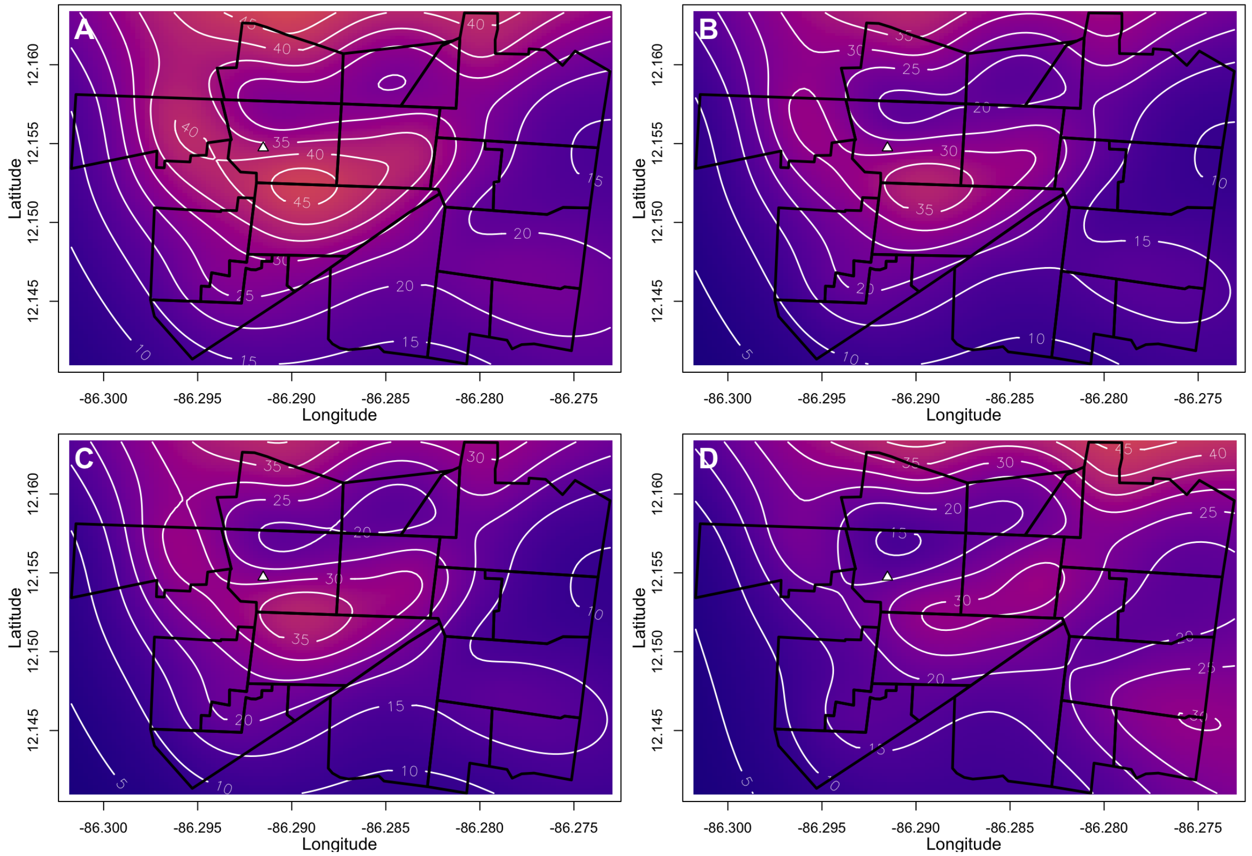


**Figure S10.** CHIKV infection risk estimated from spatial logistic GAMs during the second chikungunya epidemic. Panels show **(A)** unadjusted risk; **(B)** infection risk adjusted for age and sex; **(C)** infection risk adjusted for age, sex, and water availability; and **(D)** infection risk adjusted for age, sex, water availability, and distance to the cemetery. The variables in models for Panels B-D have been set to those of the median participant across the three PDCS epidemics to facilitate cross-epidemic comparisons. The median participant is a female of age 8.71 years living in a household that has 24-hour indoor access to tap water and is located 841.50 meters from the boundary of the local cemetery. Figures S9-11 and S17-19 use the same color scale to facilitate cross-epidemic comparisons in the PDCS population and to ensure visual comparability between maps that share the same median values.

Abbreviations: CHIKV, chikungunya virus; GAM, generalized additive model; PDCS, Pediatric Dengue Cohort Study

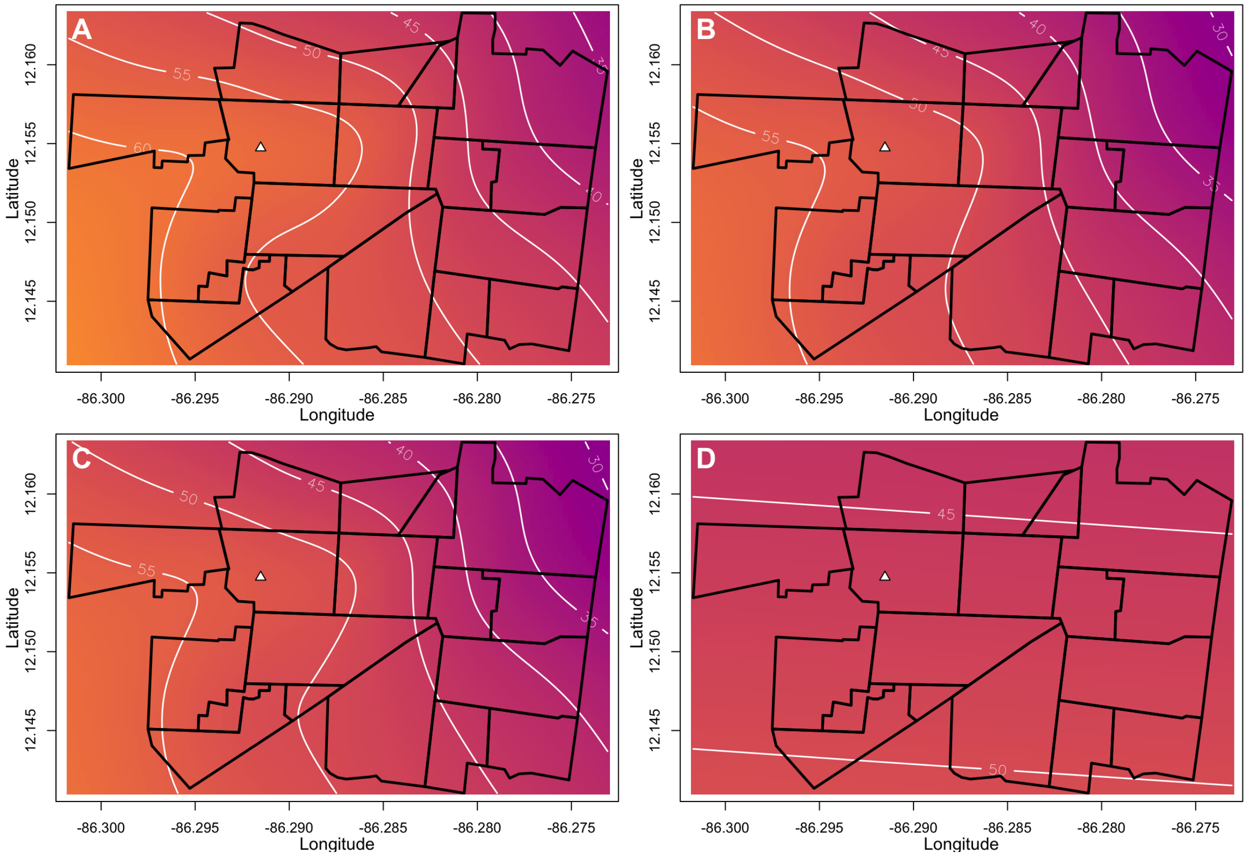


**Figure S11.** ZIKV infection risk estimated from spatial logistic GAMs during the Zika epidemic. Panels show **(A)** unadjusted infection risk; **(B)** infection risk adjusted for age and sex; **(C)** infection risk adjusted for age, sex, and water availability; and **(D)** infection risk adjusted for age, sex, water availability, and distance to the cemetery. The variables in models for Panels B-D have been set to those of the median participant across the three PDCS epidemics to facilitate cross-epidemic comparisons. The median participant is a female of age 8.71 years living in a household that has 24-hour indoor access to tap water and is located 841.50 meters from the boundary of the local cemetery. Figures S9-11 and S17-19 use the same color scale to facilitate cross-epidemic comparisons in the PDCS population and to ensure visual comparability between maps that share the same median values.

Abbreviations: GAM, generalized additive model; PDCS, Pediatric Dengue Cohort Study; ZIKV, Zika virus

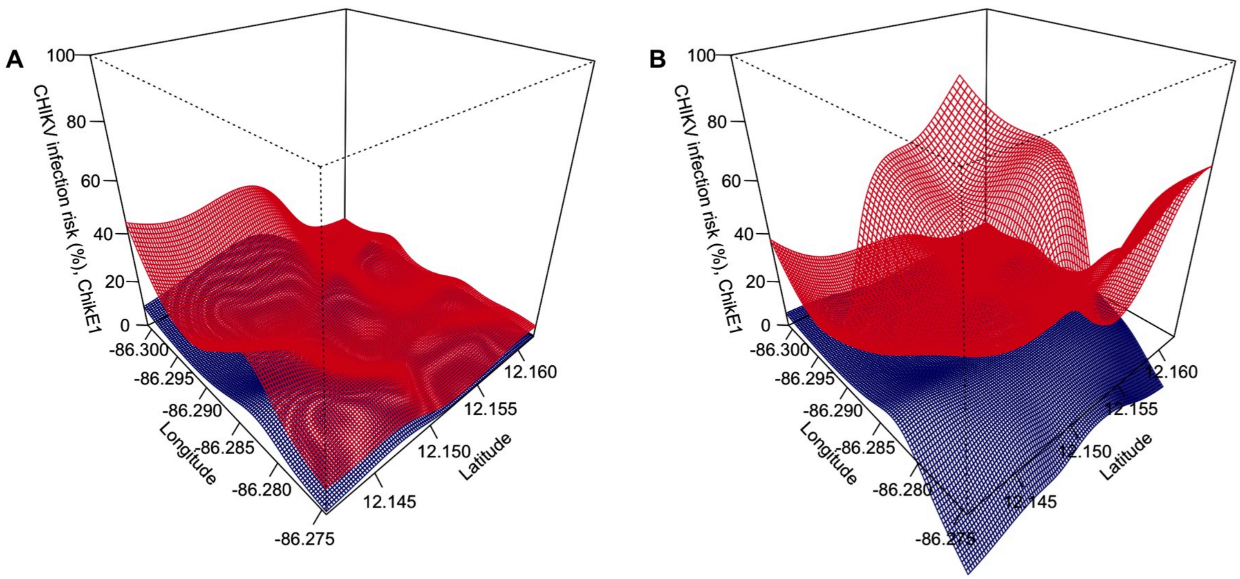


**Figure S12.** Uncertainty bounds for the unadjusted **(A)** and adjusted **(B)** (for age, sex, water availability, and distance to the cemetery) spatial infection risk estimates from the first chikungunya epidemic. The unadjusted and fully adjusted panels from Figure S9A,D lie between the red (point estimate + 1SE surface) and the blue (point estimate – 1SE surface). The surfaces should be interpreted as indicative of where there is more certainty (red and blue surfaces are close to each other) and where there is less certainty (red and blue surfaces are far from each other). Unlike the point estimate surfaces of Figure S9, these uncertainty surfaces are not model-constrained to be between 0-1. As a result, the surfaces may exceed the [0,1] probability space, which is why they should not be interpreted as the 3D equivalent of confidence intervals or credible intervals.

Abbreviations: SE, standard error

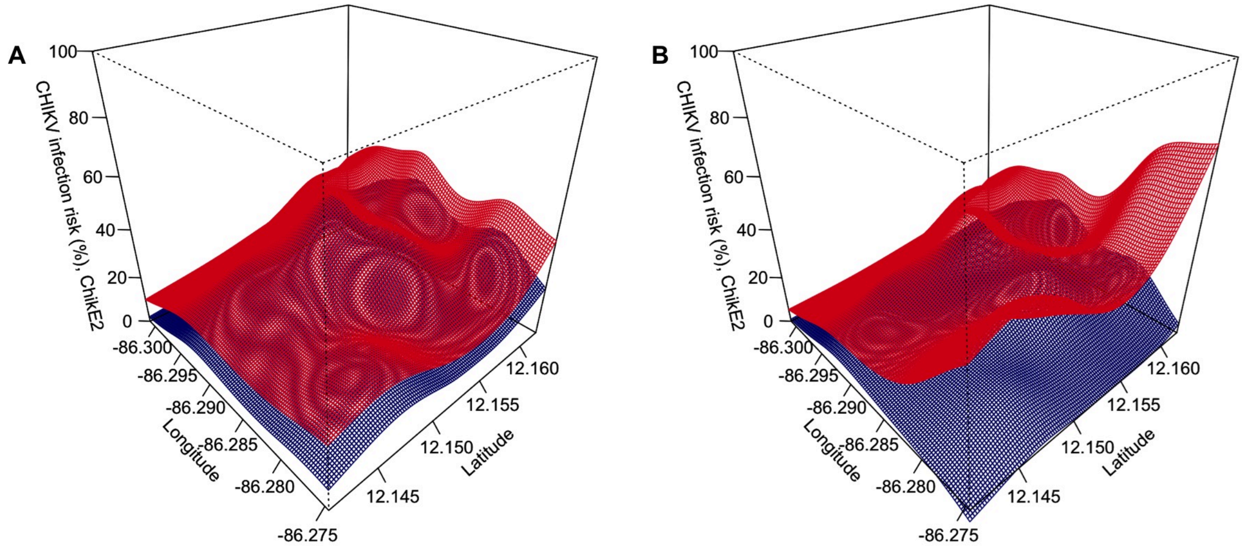


**Figure S13.** Uncertainty bounds for the unadjusted **(A)** and adjusted **(B)** (for age, sex, water availability, and distance to the cemetery) spatial infection risk estimates from the second chikungunya epidemic. The unadjusted and fully adjusted panels from Figure S10A,D lie between the red (point estimate + 1SE surface) and the blue (point estimate – 1SE surface). The surfaces should be interpreted as indicative of where there is more certainty (red and blue surfaces are close to each other) and where there is less certainty (red and blue surfaces are far from each other). Unlike the point estimate surfaces of Figure S10, these uncertainty surfaces are not model-constrained to be between 0-1. As a result, the surfaces may exceed the [0,1] probability space, which is why they should not be interpreted as the 3D equivalent of confidence intervals or credible intervals.

Abbreviations: SE, standard error

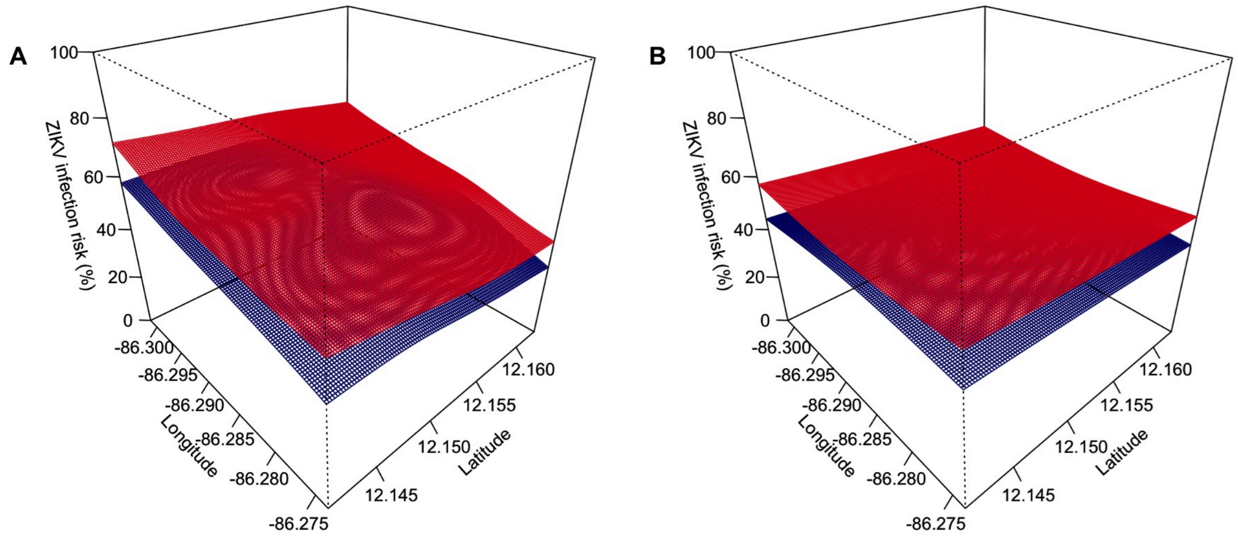


**Figure S14.** Uncertainty bounds for the unadjusted **(A)** and adjusted **(B)** (for age, sex, water availability, and distance to the cemetery) spatial infection risk estimates from the Zika epidemic. The unadjusted and fully adjusted panels from Figure S11A,D lie between the red (point estimate + 1SE surface) and the blue (point estimate – 1SE surface). The surfaces should be interpreted as indicative of where there is more certainty (red and blue surfaces are close to each other) and where there is less certainty (red and blue surfaces are far from each other). Unlike the point estimate surfaces of Figure S11, these uncertainty surfaces are not model-constrained to be between 0-1. As a result, the surfaces may exceed the [0,1] probability space, which is why they should not be interpreted as the 3D equivalent of confidence intervals or credible intervals.

Abbreviations: SE, standard error

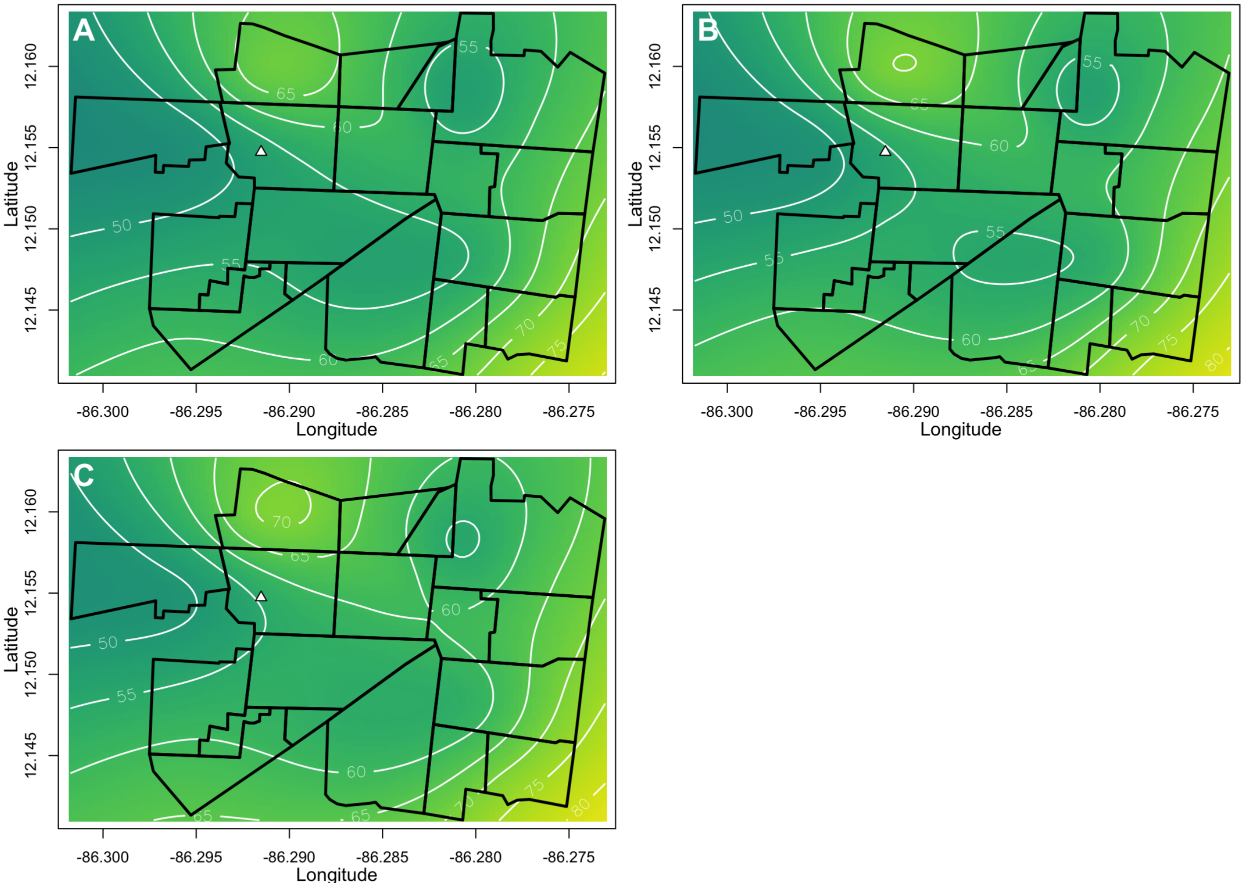


**Figure S15.** SARS-CoV-2 infection risk estimated from spatial logistic GAMs during the COVID-19 epidemic. Panels show **(A)** unadjusted infection risk; **(B)** infection risk adjusted for age, and **(C)** infection risk adjusted for age and sex. The variables in models for Panels B-C have been set to those of the median participant in the HICS population: a female of age 17.1 years. Figures S15 and S20 use the same color scale to facilitate comparisons in the HICS population and to ensure visual comparability between maps that share the same median values.

Abbreviations: COVID-19, coronavirus disease 2019; GAM, generalized additive model; HICS, Household Influenza Cohort Study; SARS-CoV-2, severe acute respiratory syndrome coronavirus 2

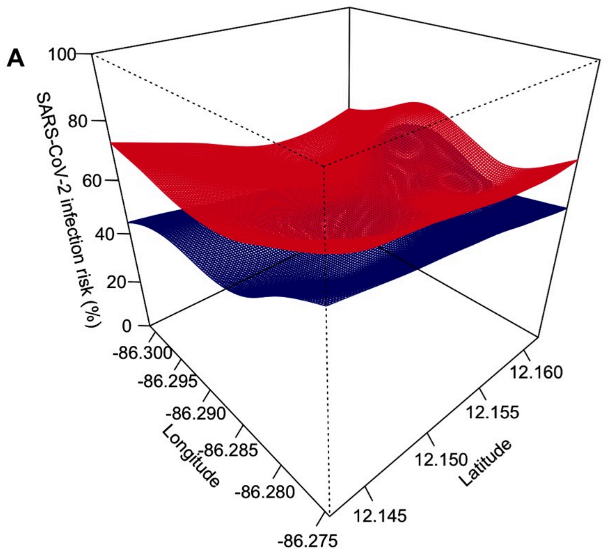


**Figure S16.** Uncertainty bounds for the unadjusted **(A)** and adjusted **(B)** (for age and sex) spatial infection risk estimates from the COVID-19 epidemic. The unadjusted and fully adjusted panels from Figure S15A,C lie between the red (point estimate + 1SE surface) and the blue (point estimate – 1SE surface). The surfaces should be interpreted as indicative of where there is more certainty (red and blue surfaces are close to each other) and where there is less certainty (red and blue surfaces are far from each other). Unlike the point estimate surfaces of Figure S15, these uncertainty surfaces are not model-constrained to be between 0-1. As a result, the surfaces may exceed the [0,1] probability space, which is why they should not be interpreted as the 3D equivalent of confidence intervals or credible intervals.

Abbreviations: SE, standard error

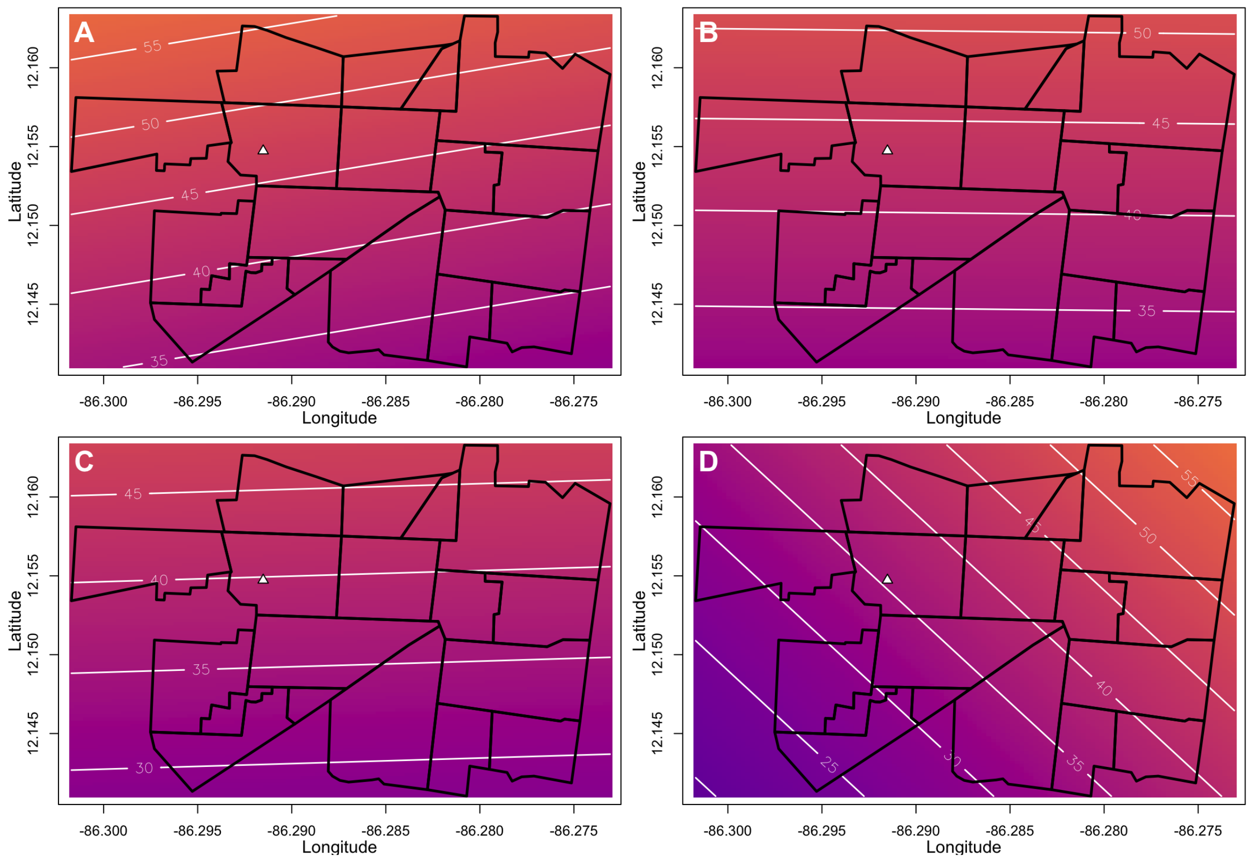


**Figure S17.** The risk of chikungunya given CHIKV infection estimated from spatial logistic GAMs during the first chikungunya epidemic. Panels show the **(A)** unadjusted risk of disease; **(B)** the risk of disease adjusted for age; **(C)** the risk of disease adjusted for age and sex; and **(D)** the risk of disease adjusted for age, sex, and distance to the cemetery. Distance to the cemetery was included in these model visualizations to examine the hypothesis that the spatial variation in disease risk is due to elevated CHIKV viremia from multiple mosquito bites, which would be likelier closer to the cemetery than away from it. The variables in models for Panels B-D have been set to those of the median participant across the three PDCS epidemics to facilitate cross-epidemic comparisons. The median participant is a female of age 8.71 years living in a household that is located 841.50 meters from the boundary of the local cemetery. Figures S9-11 and S17-19 use the same color scale to facilitate cross-epidemic comparisons in the PDCS population and to ensure visual comparability between maps that share the same median values.

Abbreviations: CHIKV, chikungunya virus; GAM, generalized additive model; PDCS, Pediatric Dengue Cohort Study

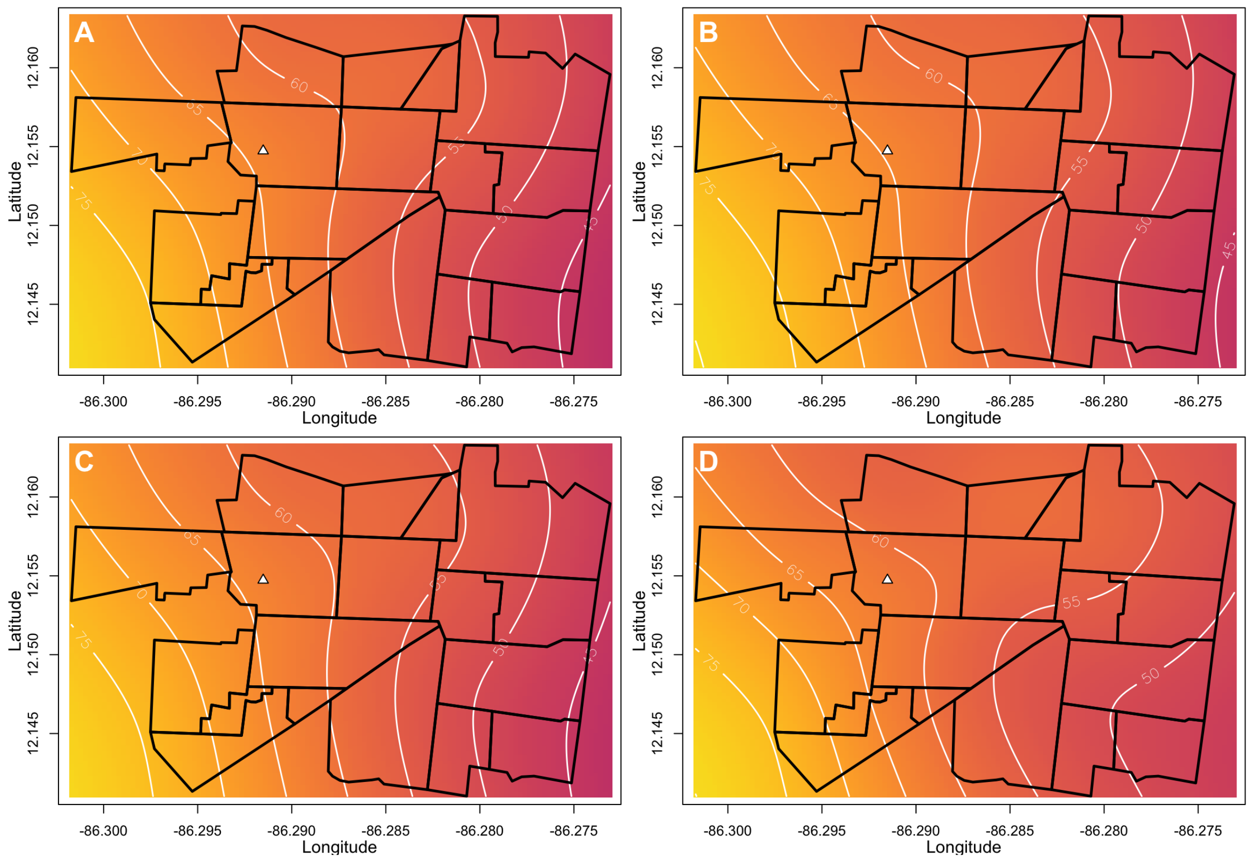


**Figure S18.** The risk of chikungunya given CHIKV infection estimated from spatial logistic GAMs during the second chikungunya epidemic. Panels show the **(A)** unadjusted risk of disease; **(B)** the risk of disease adjusted for age; **(C)** the risk of disease adjusted for age and sex; and **(D)** the risk of disease adjusted for age, sex, and distance to the cemetery. Distance to the cemetery was included in these model visualizations to examine the hypothesis that the spatial variation in disease risk is due to elevated CHIKV viremia from multiple mosquito bites, which would be likelier closer to the cemetery than away from it. The variables in models for Panels B-D have been set to those of the median participant across the three PDCS epidemics to facilitate cross-epidemic comparisons. The median participant is a female of age 8.71 years living in a household that is located 841.50 meters from the boundary of the local cemetery. Figures S9-11 and S17-19 use the same color scale to facilitate cross-epidemic comparisons in the PDCS population and to ensure visual comparability between maps that share the same median values.

Abbreviations: CHIKV, chikungunya virus; GAM, generalized additive model; PDCS, Pediatric Dengue Cohort Study

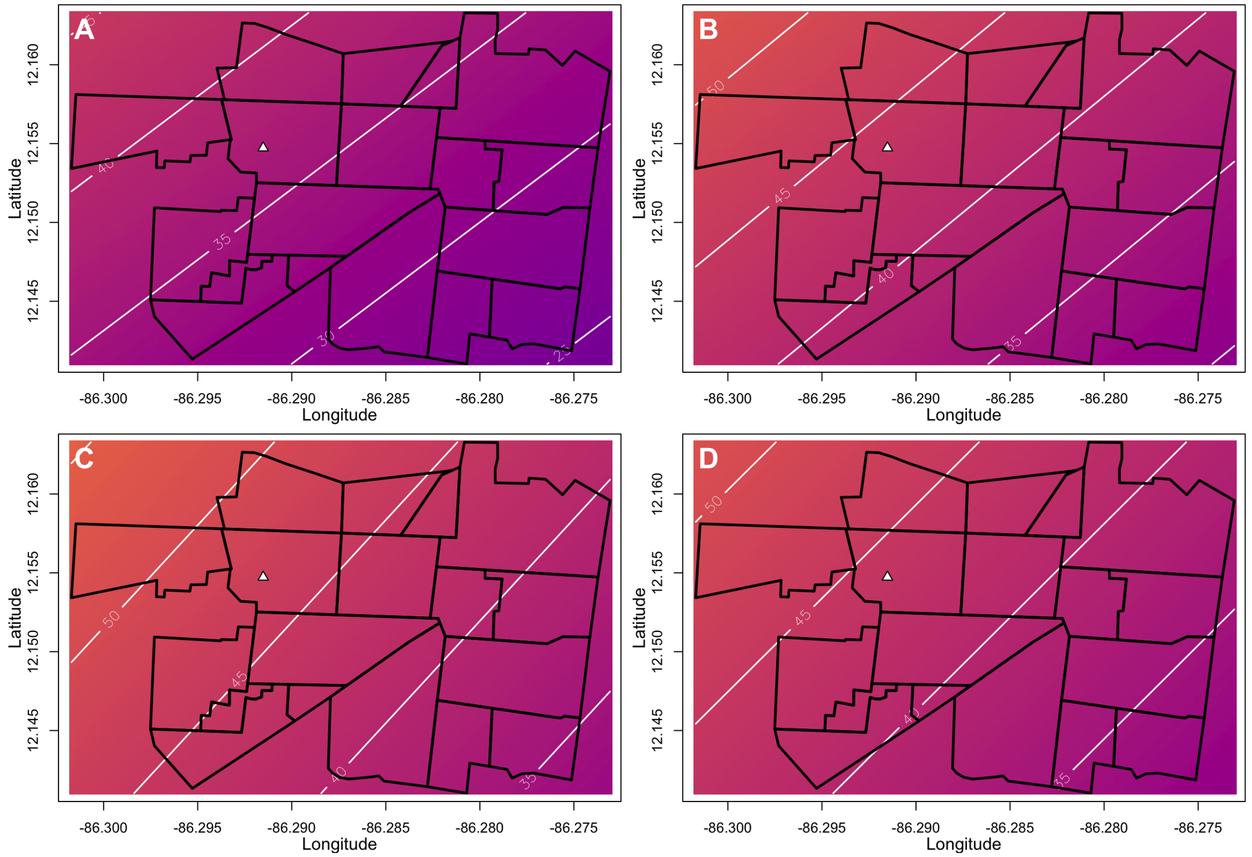


**Figure S19.** The risk of Zika given ZIKV infection estimated from spatial logistic GAMs during the Zika epidemic. Panels show the **(A)** unadjusted risk of disease; **(B)** the risk of disease adjusted for age and sex; **(C)** the risk of disease adjusted for age, sex, and prior DENV infection; and **(D)** the risk of disease adjusted for age, sex, and distance to the cemetery. Distance to the cemetery was included in these model visualizations to examine the hypothesis that the spatial variation in disease risk is due to elevated ZIKV viremia from multiple mosquito bites, which would be likelier closer to the cemetery than away from it. The variables in models for Panels B-D have been set to those of the median participant across the three PDCS epidemics to facilitate cross-epidemic comparisons. The median participant is a female of age 8.71 years living in a household that is located 841.50 meters from the boundary of the local cemetery. Figures S9-11 and S17-19 use the same color scale to facilitate cross-epidemic comparisons in the PDCS population and to ensure visual comparability between maps that share the same median values.

Abbreviations: DENV, dengue virus; GAM, generalized additive model; PDCS, Pediatric Dengue Cohort Study; ZIKV, Zika virus

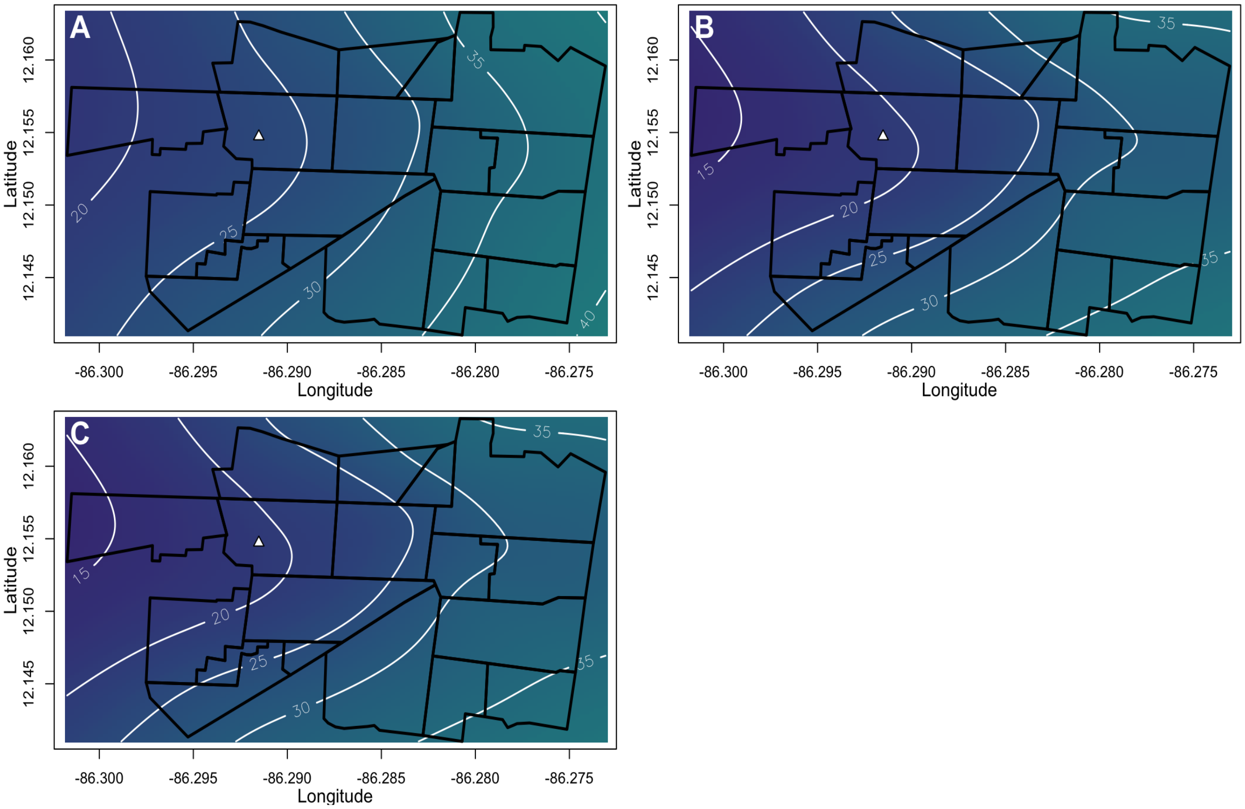


**Figure S20.** The risk of COVID-19 given SARS-CoV-2 infection estimated from spatial logistic GAMs during the COVID-19 epidemic. Panels show **(A)** unadjusted infection risk; **(B)** infection risk adjusted for age, and **(C)** infection risk adjusted for age and sex. The variables in models for Panels B-C have been set to those of the median participant in the HICS population: a female of age 17.1 years. Figures S15 and S20 use the same color scale to facilitate comparisons in the HICS population and to ensure visual comparability between maps that share the same median values.

Abbreviations: COVID-19, coronavirus disease 2019; GAM, generalized additive model; HICS, Household Influenza Cohort Study; SARS-CoV-2, severe acute respiratory syndrome coronavirus 2

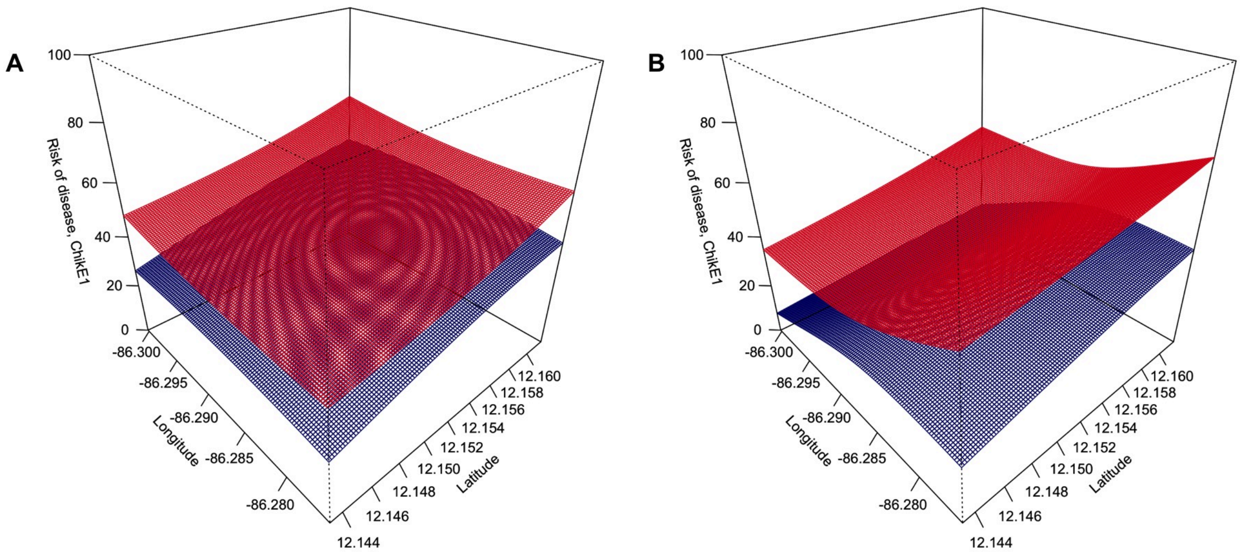


**Figure S21.** Uncertainty bounds for the unadjusted **(A)** and adjusted **(B)** (for age, sex, and the distance to the cemetery) spatial estimates of the risk of chikungunya given CHIKV infection from the first chikungunya epidemic. The unadjusted and fully adjusted panels from Figure S17A,D lie between the red (point estimate + 1SE surface) and the blue (point estimate – 1SE surface). The surfaces should be interpreted as indicative of where there is more certainty (red and blue surfaces are close to each other) and where there is less certainty (red and blue surfaces are far from each other). Unlike the point estimate surfaces of Figure S17, these uncertainty surfaces are not model-constrained to be between 0-1. As a result, the surfaces may exceed the [0,1] probability space, which is why they should not be interpreted as the 3D equivalent of confidence intervals or credible intervals.

Abbreviations: CHIKV, chikungunya virus; SE, standard error

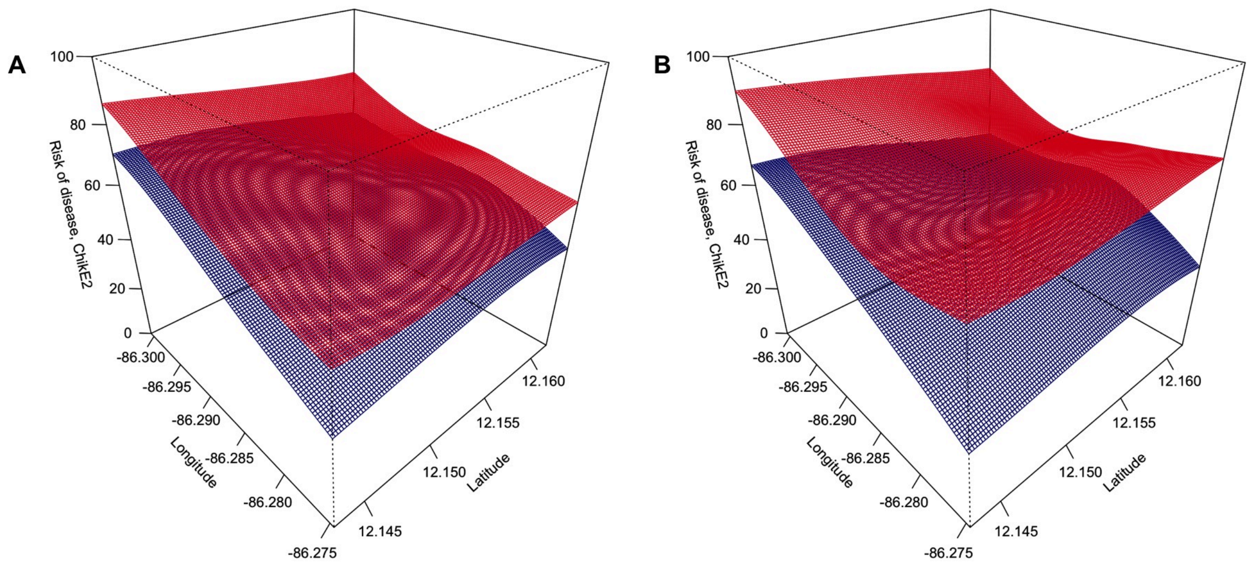


**Figure S22.** Uncertainty bounds for the unadjusted **(A)** and adjusted **(B)** (for age, sex, and the distance to the cemetery) spatial estimates of risk of chikungunya given CHIKV infection from the second chikungunya epidemic. The unadjusted and fully adjusted panels from Figure S18A,D lie between the red (point estimate + 1SE surface) and the blue (point estimate – 1SE surface). The surfaces should be interpreted as indicative of where there is more certainty (red and blue surfaces are close to each other) and where there is less certainty (red and blue surfaces are far from each other). Unlike the point estimate surfaces of Figure S18, these uncertainty surfaces are not model-constrained to be between 0-1. As a result, the surfaces may exceed the [0,1] probability space, which is why they should not be interpreted as the 3D equivalent of confidence intervals or credible intervals.

Abbreviations: CHIKV, chikungunya virus; SE, standard error

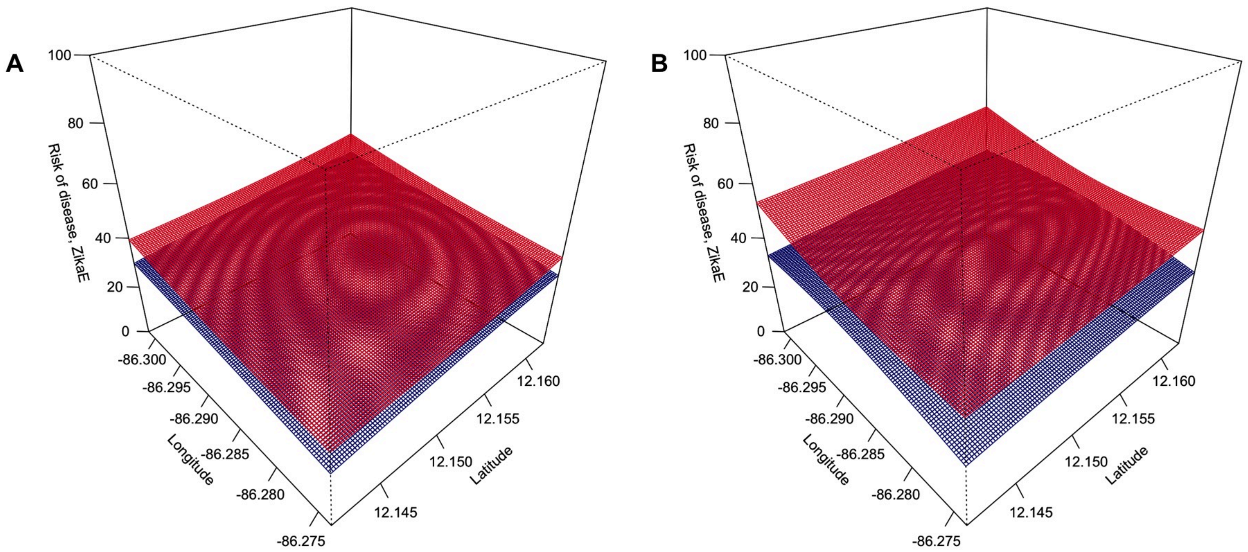


**Figure S23.** Uncertainty bounds for the unadjusted **(A)** and adjusted **(B)** (for age, sex, prior DENV infection, and the distance to the cemetery) spatial estimates of the risk of Zika given ZIKV infection from the Zika epidemic. The unadjusted and fully adjusted panels from Figure S19A,D lie between the red (point estimate + 1SE surface) and the blue (point estimate – 1SE surface). The surfaces should be interpreted as indicative of where there is more certainty (red and blue surfaces are close to each other) and where there is less certainty (red and blue surfaces are far from each other). Unlike the point estimate surfaces of Figure S19, these uncertainty surfaces are not model-constrained to be between 0-1. As a result, the surfaces may exceed the [0,1] probability space, which is why they should not be interpreted as the 3D equivalent of confidence intervals or credible intervals.

Abbreviations: DENV, dengue virus; SE, standard error; ZIKV, Zika virus

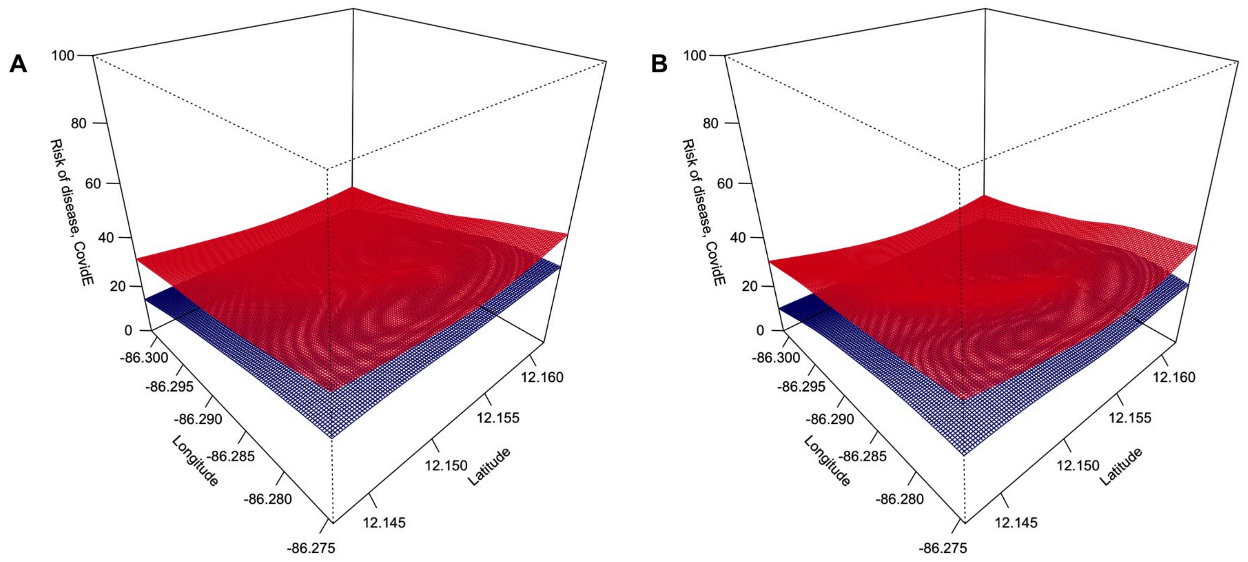


**Figure S24.** Uncertainty bounds for the unadjusted **(A)** and adjusted **(B)** (for age and sex) spatial estimates of the risk of COVID-19 given SARS-CoV-2 infection from the COVID-19 epidemic. The unadjusted and fully adjusted panels from Figure S20A,C lie between the red (point estimate + 1SE surface) and the blue (point estimate – 1SE surface). The surfaces should be interpreted as indicative of where there is more certainty (red and blue surfaces are close to each other) and where there is less certainty (red and blue surfaces are far from each other). Unlike the point estimate surfaces of Figure S20, these uncertainty surfaces are not model-constrained to be between 0-1. As a result, the surfaces may exceed the [0,1] probability space, which is why they should not be interpreted as the 3D equivalent of confidence intervals or credible intervals.

Abbreviations: COVID-19, coronavirus disease 2019; SARS-CoV-2, severe acute respiratory syndrome coronavirus 2; SE, standard error

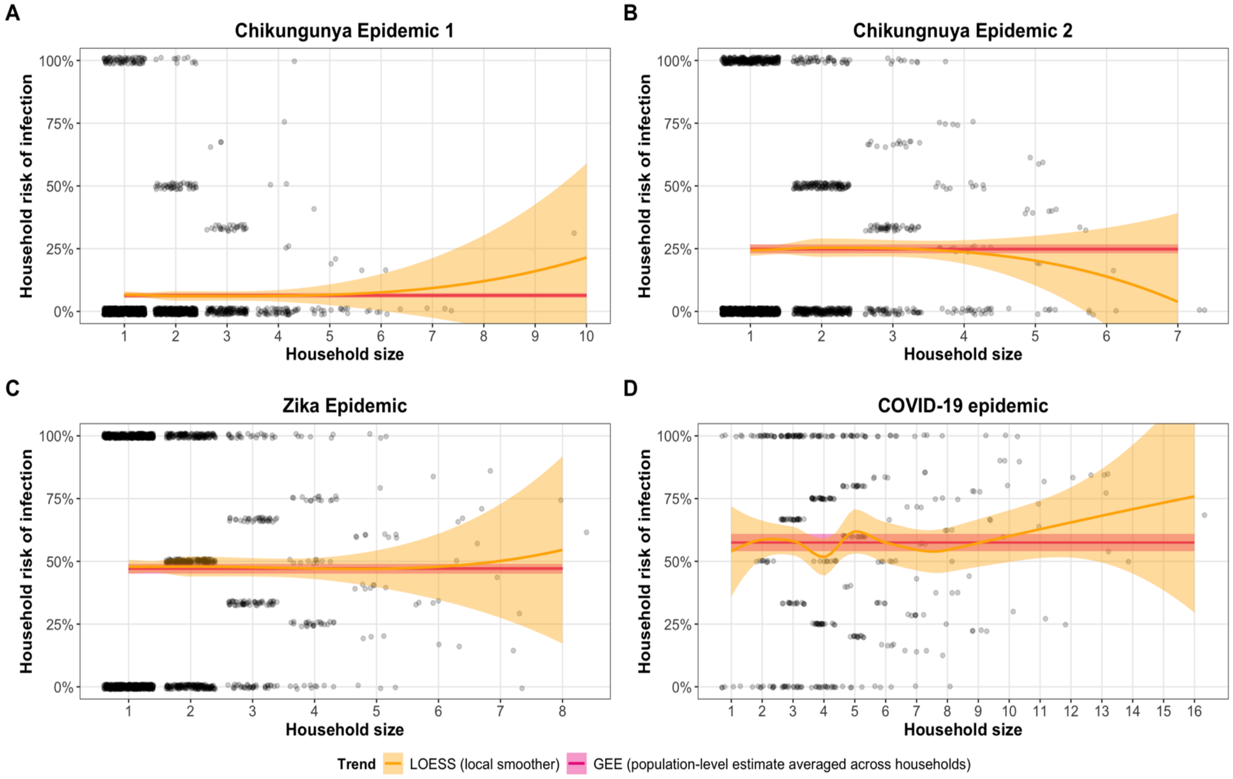


**Figure S25.** The association between household size and household infection risk (number of household members infected / household size) during **(A)** ChikE1, **(B)** ChikE2, **(C)** ZikaE, and **(D)** CovidE. Where there was sufficient data to reliably estimate a trend (PDCS households with ≤5 participants and HICS households with ≤8 participants), there was no evidence of household infection risk scaling with household size. Both the independent and dependent variables refer only to PDCS and HICS participants, as we have no data on household members who are not participants of the cohorts. The yellow line is a smooth LOESS trend. The pink line is the population-averaged intercept-only GEE estimate of infection risk, which averages over households of different sizes. A 95% confidence band is shown for each estimate in the corresponding color. Note that the confidence band for the LOESS trend dips below 0 because there is no numerical constraint on the LOESS smoother. A GAM approach failed to converge because of insufficient variation in values of household infection risk, hence the use of the less robust LOESS smoother.

Abbreviations: ChikE1, Chikungunya epidemic 2; ChikE2, chikungunya epidemic 2; CovidE, COVID-19 epidemic; GEE, generalized estimating equations; LOESS, locally estimated scatterplot smoothing; GAM, generalized additive model; ZikaE, Zika epidemic

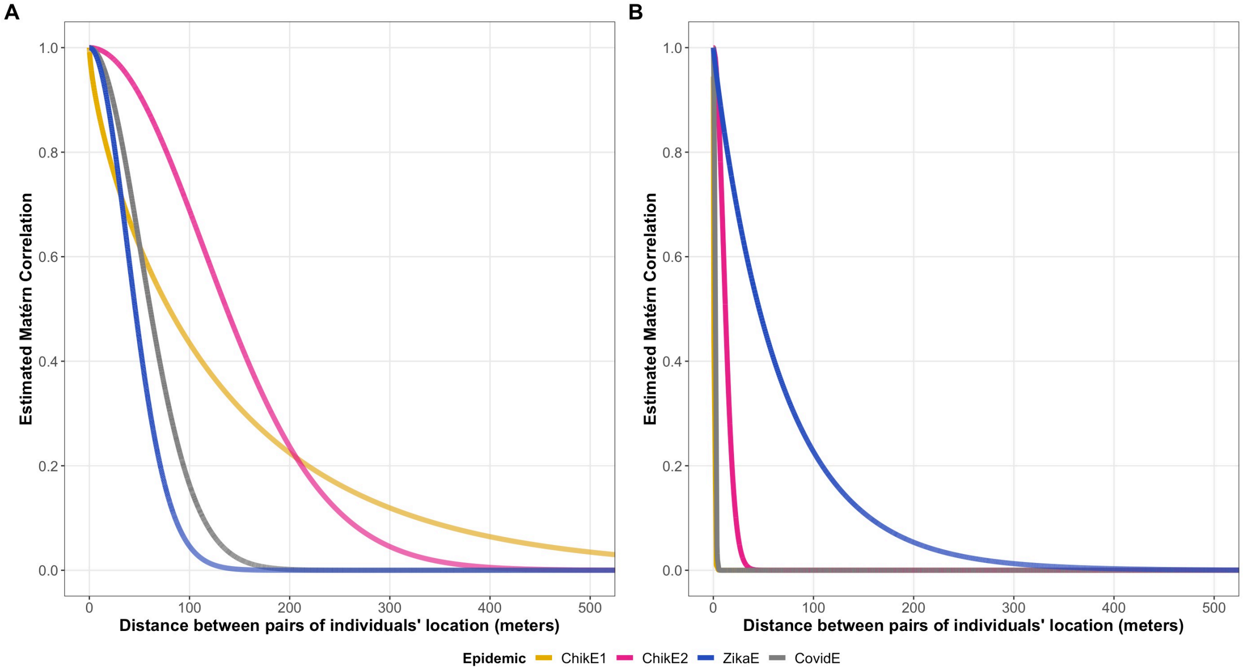


**Figure S26.** Spatial correlation effect of infection outcomes **(A)** and disease outcomes given infection **(B)** across the four epidemics we considered. The spatial correlation of infection outcomes was the primary analysis since CHIKV and ZIKV are spread by *Aedes* mosquitoes, which have a limited flight range; the corresponding analysis for disease outcomes among infected persons is shown for completeness. The estimated Matérn autocorrelation function for infection and disease outcomes between two locations against their distance is shown. For example, the CHIKV infection outcomes of persons living >200 meters apart during ChikE1 and ChikE2 have a correlation <0.2, on average. The Matérn autocorrelation parameters underlying the functions shown were estimated from the logistic geostatistical mixed models for infection and disease given infection that account for spatial and household correlation (Table S5-S6). Unlike all other lines in this figure, the pink and gray lines in B corresponds to geostatistical models that only account for spatial correlation. In the models that account for both sources of possible correlation, the random effects for the household terms had an overinflated variance, resulting in biologically implausible Matérn autocorrelation functions. Given the similarity of all other components of the models (Table S4) and the lack of household-based clustering of disease outcomes (Table S5) during ChikE2 and CovidE, the pink and gray lines in B reflect the estimated Matérn autocorrelation function for geostatistical models without accounting for household-based clustering. The slow decay of the autocorrelation function for disease outcomes during ZikaE suggests either that ZIKV infections within 200 meters of each other mutually affected disease outcomes or (more likely) unknown variables relevant to the spatial distribution of Zika occurrence given ZIKV infection were not included in the model.

Abbreviations: ChikE1, chikungunya epidemic 1; ChikE2, chikungunya epidemic 2; CHIKV, chikungunya virus; CovidE, COVID-19 epidemic; ZikaE, Zika epidemic

21. World Health Organization. Dengue haemorrhagic fever: Diagnosis, treatment, prevention, and control, 2nd ed. Geneva; 1997.

22. World Health Organization. Dengue : Guidelines for Diagnosis Treatment Prevention and Control (New Edition 2009). World Health Organization; 2009. 158 p.

23. World Health Organization. Proposed case definition of Chikungunya Fever (WHO, SEARO). 2008.

24. ProMED. Doença misteriosa assusta população de Camaçari. 2015.

25. Aguiar BS, Lorenz C, Virginio F, Suesdek L, Chiaravalloti-Neto F. Potential risks of Zika and chikungunya outbreaks in Brazil: A modeling study. Int J Infect Dis. 2018 May 1;70:20–9.

26. McHale T, Romero-Vivas C, Fronterre C, Arango-Padilla P, Waterlow N, Nix C, et al. Spatiotemporal heterogeneity in the distribution of chikungunya and Zika virus case incidences during their 2014 to 2016 epidemics in Barranquilla, Colombia. Int J Environ Res Public Health. 2019 May 2;16(10).

27. Campos de Lima EE, Gayawan E, Baptista EA, Queiroz BL. Spatial pattern of COVID-19 deaths and infections in small areas of Brazil. PLoS One. 2021 Feb 1;16(2 Febuary).

28. Bonilla-Aldana DK, Bonilla-Aldana JL, García-Bustos JJ, Lozada CO, Rodríguez-Morales AJ. Geographical trends of chikungunya and Zika in the Colombian Amazonian gateway department, Caqueta, 2015–2018 – Implications for public health and travel medicine. Travel Med Infect Dis. 2020 May 1;35.

29. Vissoci JRN, Rocha TAH, Silva NC da, de Sousa Queiroz RC, Thomaz EBAF, Amaral PVM, et al. Zika virus infection and microcephaly: Evidence regarding geospatial associations. PLoS Negl Trop Dis. 2018 Apr 25;12(4).

30. R Core Team. R: A language and environment for statistical computing. R Foundation for Statistical Computing. Vienna: R Foundation for Statistical Computing; 2019.

31. Kovalchik S, Varadhan R. Fitting additive binomial regression models with the R package blm. J Stat Software2. 2013;54(1):1–18.

32. Liang K-Y, Zeger SL. Longitudinal data analysis using generalized linear models. Biometrika. 1986;73(1):13–22.

33. McCullagh P, Nelder J. Generalized linear models. 2nd ed. Boca Raton, Florida: Chapman and Hall / CRC Press; 1989.

41. Wood SN. Generalized Additive Models: An Introduction with R. 2nd ed. Chapman and Hall / CRC Press;

2017.

50. Searle SR. Linear Models. New York City, USA: Wiley; 1971.

51. Wolak ME, Fairbairn DJ, Paulsen YR. Guidelines for Estimating Repeatability. Methods in Ecology and Evolution. 2012. p. 129–37.

52. Chakraborty H, Sen P. Resampling method to estimate intra-cluster correlation for clustered binary data. Commun Stat - Theory Methods. 2016;45(8):2368–77.

53. Hossain A, Chakraborty H. ICCbin: Facilitates clustered binary data generation, and estimation of intracluster correaltion coefficient (ICC) for binary data. 2017.

59. Kulldorff M. SaTScan. 2016.

60. Han J, Zhu L, Kulldorff M, Hostovich S, Stinchcomb DG, Tatalovich Z, et al. Using Gini coefficient to determining optimal cluster reporting sizes for spatial scan statistics. Int J Health Geogr. 2016 Aug 3;15(1):27.

61. Moran PAP. Notes on Continuous Stochastic Phenomena. Biometrika. 1950 Jun;37(1/2):17.

78. Sturrock H. Fitting point process models to disease case data with mgcv (or whatever you want). GitHub. 2020.

79. Pebesma E. Simple features for R: Standardized support for spatial vector data. R J. 2018;10(1):439–46.

80. Sturrock H. disarmr: A collection of spatial analysis functions related to the DiSARM project at UCSF. 2020.

81. Perpinan Lamigueiro O, Hijams RJ. rasterVis. 2019.

82. Agudie B. gridExtra: Miscellaneous functions for "Grid" graphics. 2017.

83. Horstick O, Jaenisch T, Martinez E, Kroeger A, See LLC, Farrar J, et al. Comparing the usefulness of the 1997 and 2009 WHO dengue case classification: A systematic literature review. Am J Trop Med Hyg. 2014;91(3):621–34.

84. WHO. Zika virus disease | Interim case definition. World Health Organization; 2016.

85. PAHO/WHO. Zika Resources: Case Definitions. 2016.

86. Lindsey NP, Staples JE, Powell K, Rabe IB, Fischer M, Powers AM, et al. Ability To Serologically Confirm Recent Zika Virus Infection in Areas with Varying Past Incidence of Dengue Virus Infection in the United States and U.S. Territories in 2016. J Clin Microbiol. 2017 Jan 1;56(1).

87. Wickham H. ggplot2: Elegant Graphics for Data Analysis. Springer-Verlag New York; 2016.

88. Wickham H. ggplot2. 1st ed. New York City: Springer; 2009. 1–212 p.

89. Pedersen TL. patchwork: The Composer of Plots. 2019.

90. Garnier S. viridis: Default Color Maps from "matplotlib." 2018.

91. Garnier S, Ross N, Rudis R, Camargo A, Sciaini M, Scherer C. viridis - colorblind-friendly color maps for R. 2021.
92. RStudio Team. RStudio: Integrated Development for R. Boston: RStudio, Inc.; 2016.
93. Wickham H, Bryan J. readxl: Read Excel Files. 2018.
94. Schwartz M. WriteXLS: Cross-Platform Perl Based R Function to Create Excel 2003 (XLS) and Excel 2007 (XLSX) Files. 2015.
95. Wickham H, François R, Henry L, Müller K. dplyr: A Grammar of Data Manipulation. 2018.
96. Harrell F, Dupont C. Hmisc: Harrell Miscellaneous. 2020.
97. Mahmoudain M. varhandle: Functions for Robust Variable Handling. 2020.
98. Bache SM, Wickham H. magrittr: A Forward-Pipe Operator for R. 2014.
